# The epidemiology of pathogens with pandemic potential: A review of key parameters and clustering analysis

**DOI:** 10.1101/2025.03.13.25323659

**Authors:** Jack Ward, Oswaldo Gressani, Sol Kim, Niel Hens, W. John Edmunds

## Abstract

**Introduction:** In the light of the COVID-19 pandemic many countries are trying to widen their pandemic planning from its traditional focus on influenza. However, it is impossible to draw up detailed plans for every pathogen with epidemic potential. We set out to try to simplify this process by reviewing the epidemiology of a range of pathogens with pandemic potential and seeing whether they fall into groups with shared epidemiological traits.

**Methods:** We reviewed the epidemiological characteristics of 19 different pathogens with pandemic potential (those on the WHO priority list of pathogens, different strains of influenza and Mpox). We extracted data on key parameters (reproduction number serial interval, proportion of presymptomatic transmission, case fatality risk and transmission route) and applied an unsupervised learning algorithm. This combined Monte Carlo sampling with ensemble clustering to classify pathogens into distinct epidemiological archetypes based on their shared characteristics.

**Results:** From 154 articles we extracted 302 epidemiological parameter estimates. The clustering algorithms categorise these pathogens into six archetypes (1) highly transmissible Coronaviruses, (2) moderately transmissible Coronaviruses, (3) high-severity contact and zoonotic pathogens, (4) Influenza viruses (5) MERS-CoV-like and (6) MPV-like.

**Conclusion:** Unsupervised learning on epidemiological data can be used to define distinct pathogen archetypes. This method offers a valuable framework to allocate emerging and novel pathogens into defined groups to evaluate common approaches for their control.

## 1. Introduction

Recent global epidemics of COVID-19 and Mpox have illustrated that we remain vulnerable to global biological incidents. Historically, pandemic preparedness strategies have been limited in scope. For instance, prior to COVID-19, the UK government’s sole pandemic plan was the 2011 Influenza Pandemic Preparedness Strategy [1,2]. This narrow focus left critical gaps in threat readiness that have been exploited by non-influenza pathogens such as SARS-CoV-2 and Mpox virus (MPV).

Given the potential health and economic impacts of pandemics, the way in which we plan for such risks needs to be revised. There are 26 viral families known to infect humans [3], but only a fraction of these viruses will possess the ability for widespread transmission in the community [4]. Historically, this fraction has been listed, based on historical outbreaks and ranked to inform policy makers on which pathogens possess the highest pandemic potential. A list-based approach, while useful, is inflexible and is rooted in responding to yesterday’s pandemic rather than proactively planning.

Categorising pathogens based on shared epidemiological traits rather than using historical lists offers a more flexible and inclusive framework for pandemic planning [4]. A trait based approach would facilitate proactive planning for emerging threats by categorising pathogens by characteristics, allowing planners to assess a wider breadth of scenarios and control measures rather than specific historical examples [4].

To address these gaps, we propose classifying pathogens into archetypes based on epidemiological traits. Using data collected from previous systematic reviews where possible, and individual papers and parameter estimation where necessary, we implement an unsupervised machine learning algorithm combining Monte Carlo sampling with ensemble clustering to identify relevant pathogen archetypes. By categorising pathogens by their key epidemiological parameters, we show how pathogens can be grouped by shared characteristics which may point to common approaches for their control.

## 2. Methods

### 2.1 Review of epidemiological parameters

We selected 19 pathogens for review based on their epidemic or pandemic potential. This list was primarily guided by the World Health Organisation’s (WHO) R&D Blueprint (as of June 2024) [5] and included: SARS-CoV-2 (Wild-Type, Alpha, Delta, Omicron), SARS-CoV-1, MERS-CoV, Crimean-Congo hemorrhagic fever orthonairovirus (CCHFV), Ebola virus (EBOV), Marburg virus (MARV), Lassa virus (LASV), Nipah virus (NiV), Rift Valley fever virus (RVFV), Zika virus (ZIKV), Mpox virus (MPV), and several influenza A viruses (H1N1, H2N2, H3N2, H1N1pdm09, and A/H5N1).

We sought to identify quantitative estimates for parameters related to each pathogen’s transmission route(s), infection timeline, and severity. Key parameters included the reproduction number (R or R_0_), overdispersion parameter for the reproduction number (*k*), incubation period, latent period, infectious period, serial interval, case fatality risk (CFR), and infection fatality risk (IFR).

Our search strategy involved systematic queries of PubMed for peer-reviewed articles and preprints. Search terms were structured around three components: pathogen name, parameter type, and “systematic review” (**Supplementary Table S1**). Studies were required to report quantitative estimates derived from primary epidemiological data within systematic reviews or meta-analyses. For pathogens lacking comprehensive systematic reviews, we conducted targeted searches using pathogen-specific terminology, without a fixed strategy. We also included articles that provided datasets allowing for parameter estimation.

### 2.2 Parameter estimation

We estimated key parameters that were not available from the literature review. Where appropriate, we estimated values for the incubation period, serial interval, R_0_ and proportion of presymptomatic transmission for selected pathogens. For the incubation period, we used the {EpiLPS} package [6–8] where publicly available data permitted. We estimated serial intervals by fitting lognormal and gamma distributions to the number of onsets for a given day, accounting for double censoring using the R package {primarycensored} [9,10]. For R_0_, we used the package {epichains} [10,11], to provide an estimate for CCHFV on the basis of data collected by the European Centre for Disease Prevention and Control [11]. Full methodological details are provided in the Supplementary Information.

### 2.3 Clustering of epidemiological parameters

We compiled epidemiological estimates for each pathogen across a set of core parameters, including R, serial interval (SI), CFR, *k*, incubation period (IP), latent period, infectious period (**Table 1**), and transmission route. For each study, we reconstructed a full probability distribution from the reported summary statistics, applying Beta distributions for proportion outcomes (CFR) and Gamma distributions for non-negative continuous parameters (R and time to key events).

**Table 1.**
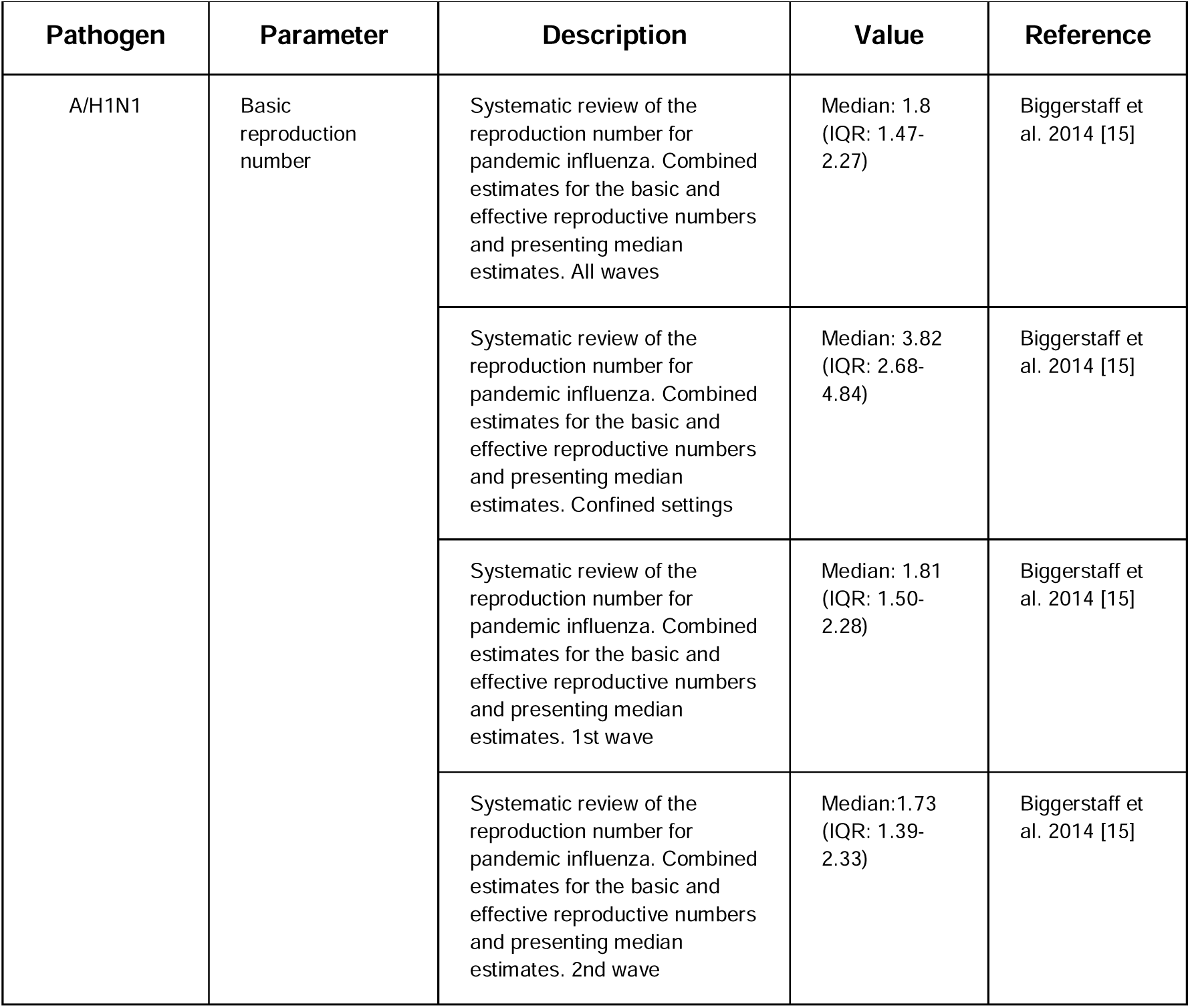
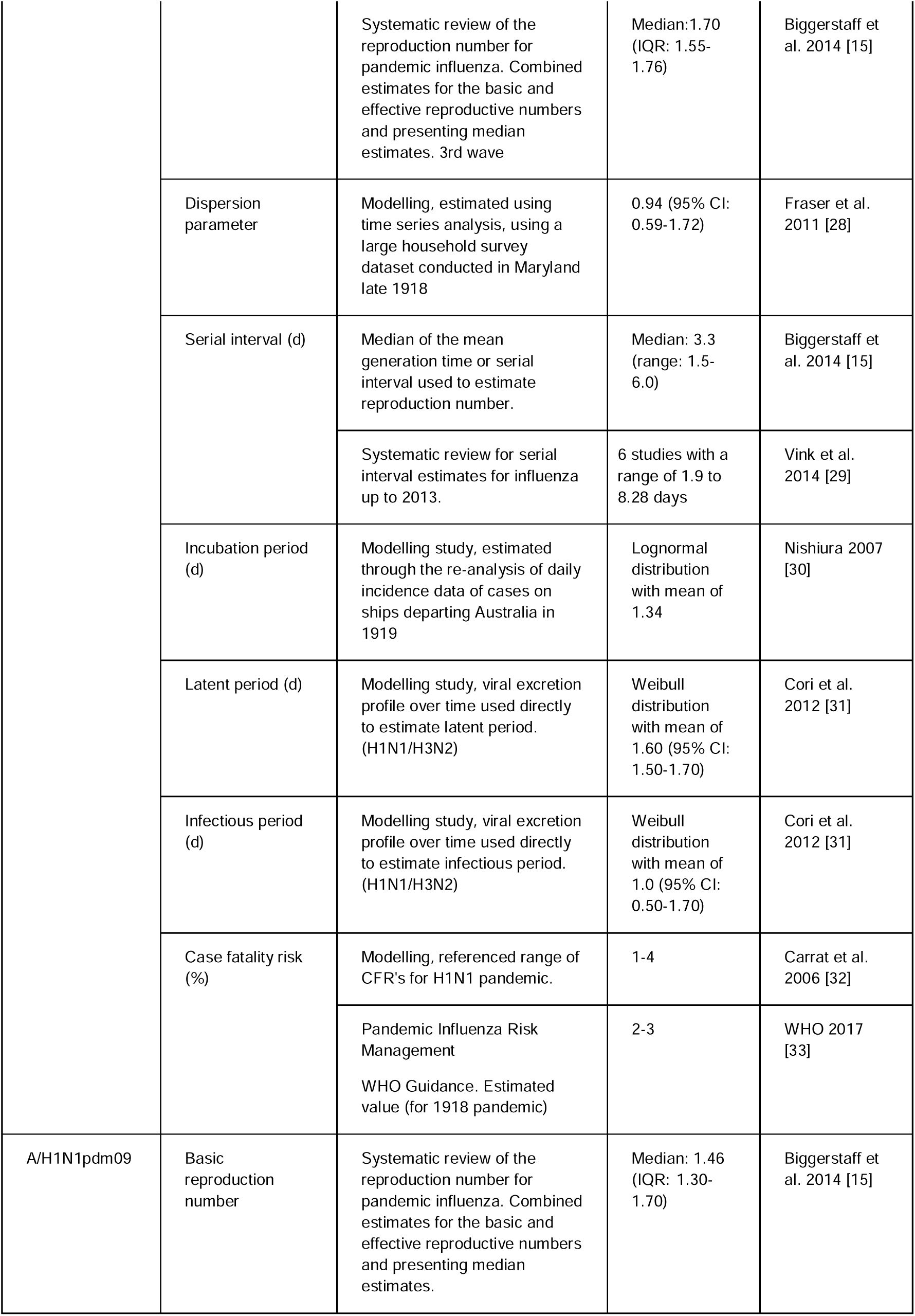
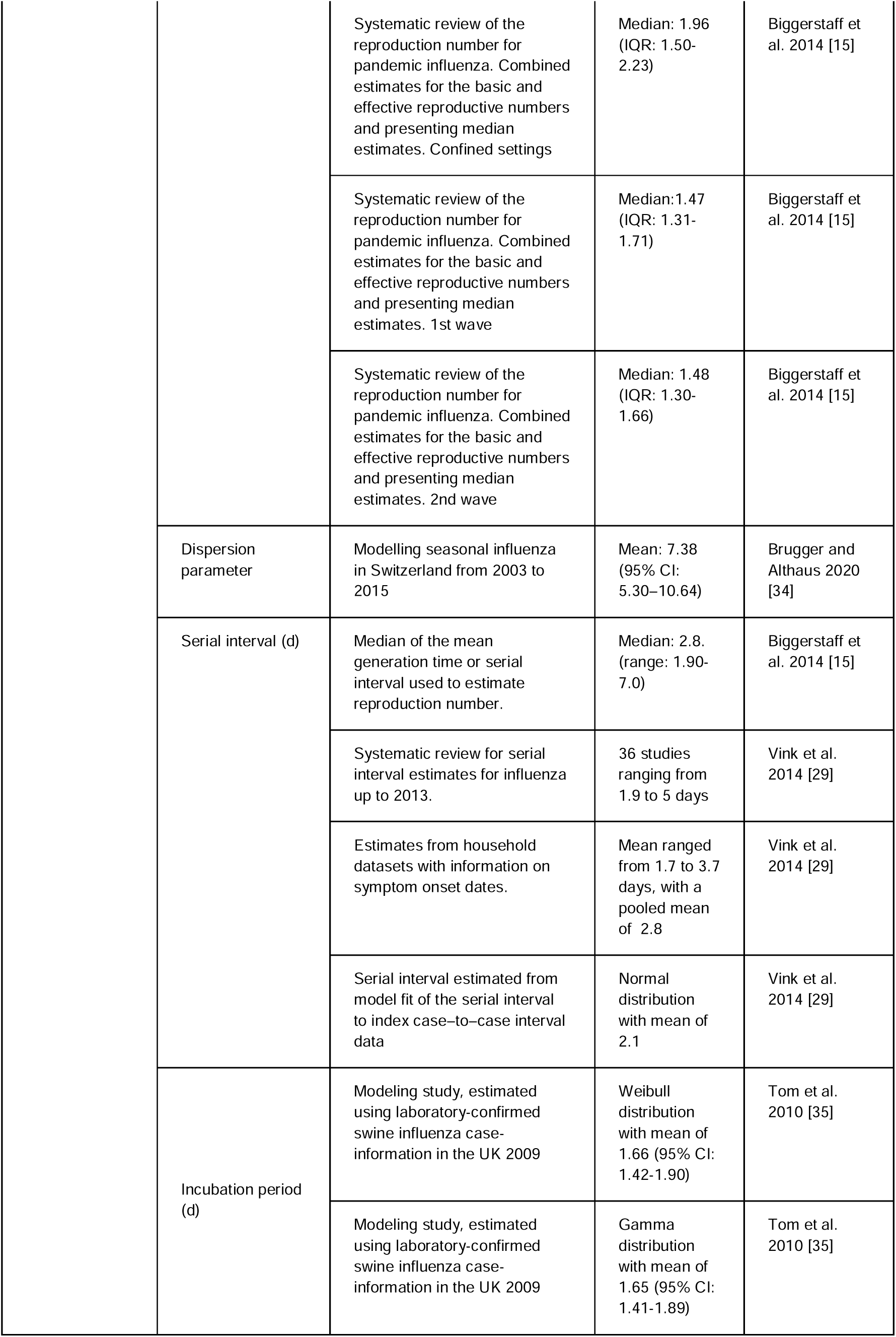
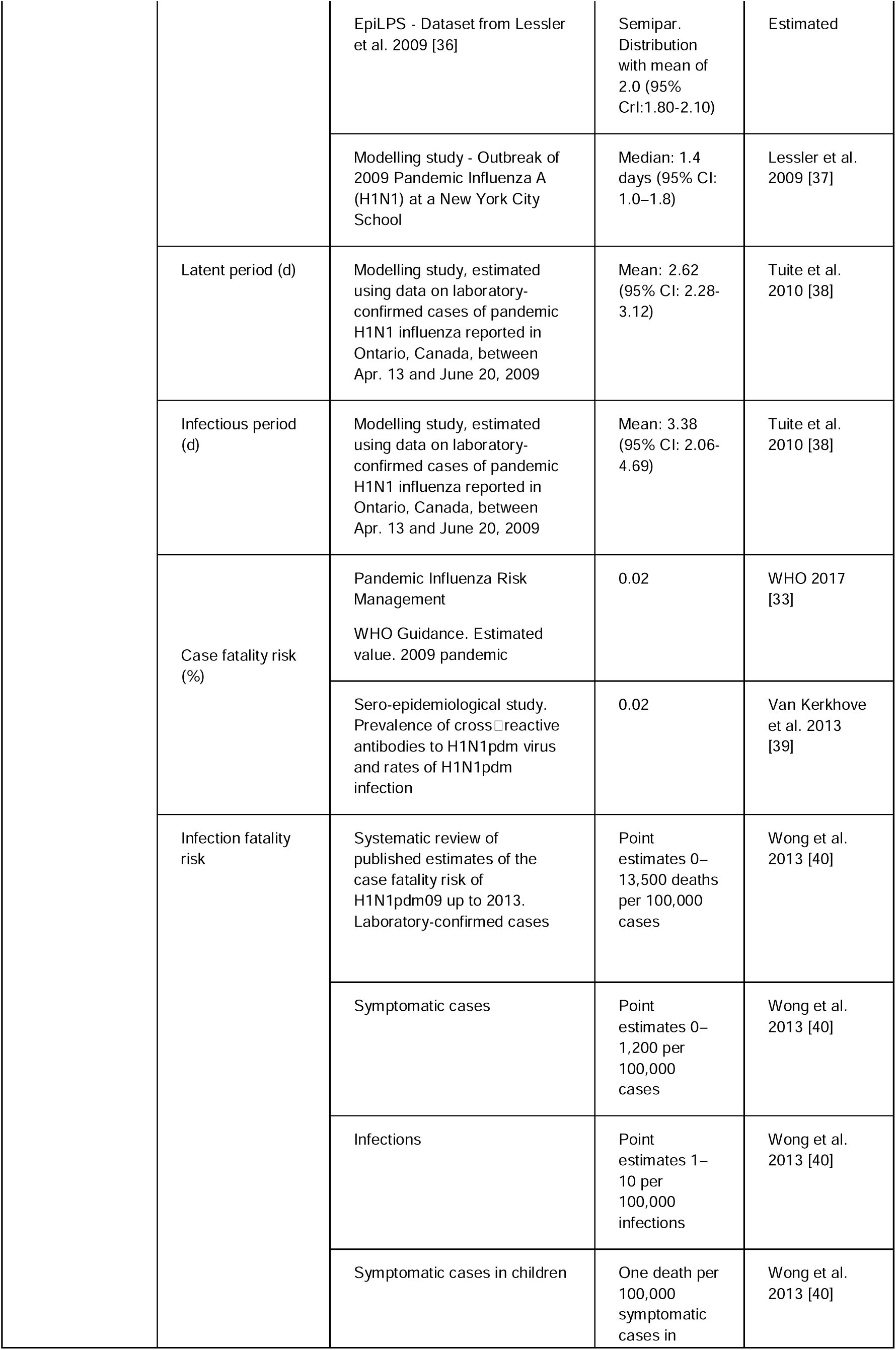
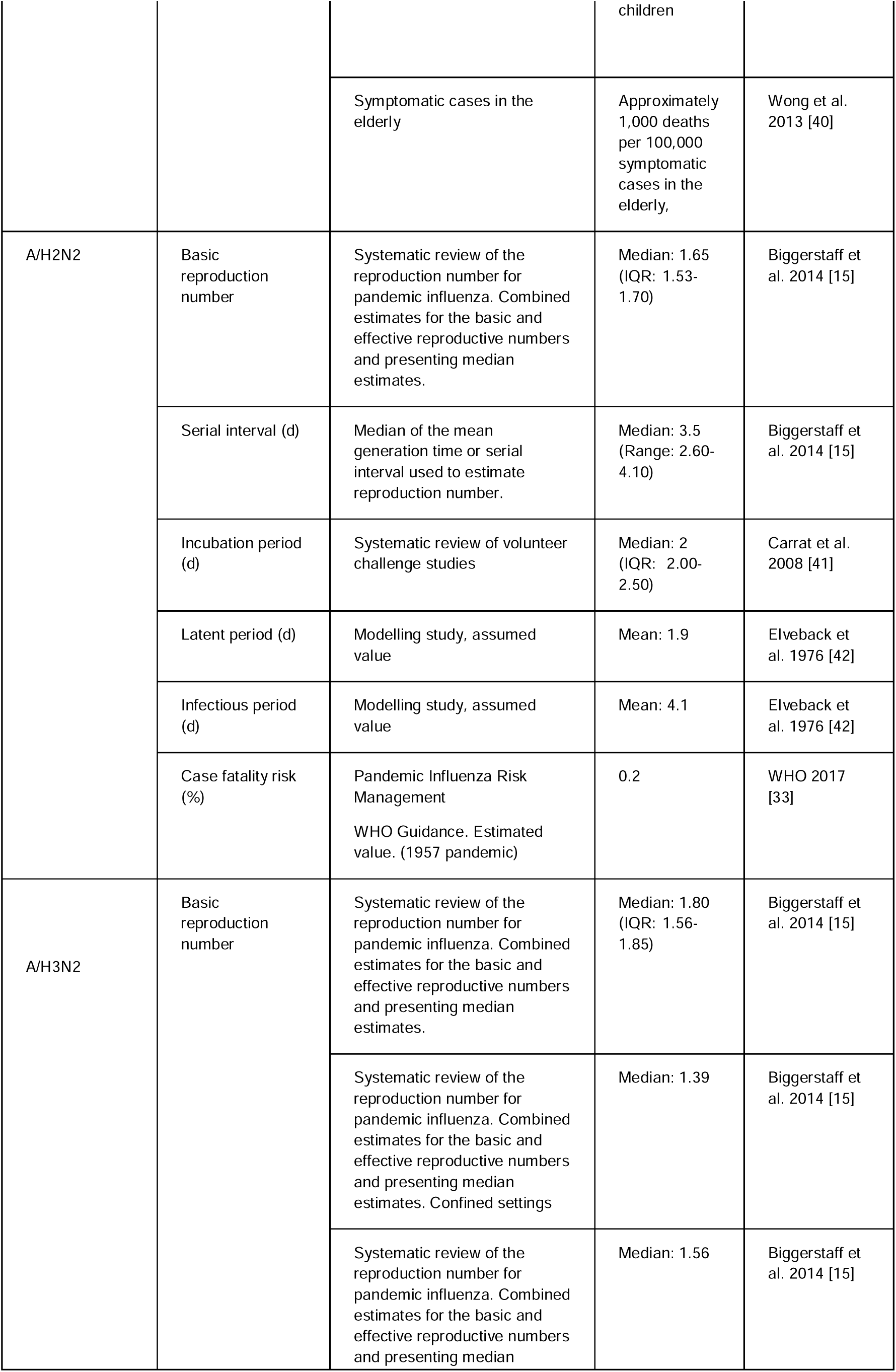
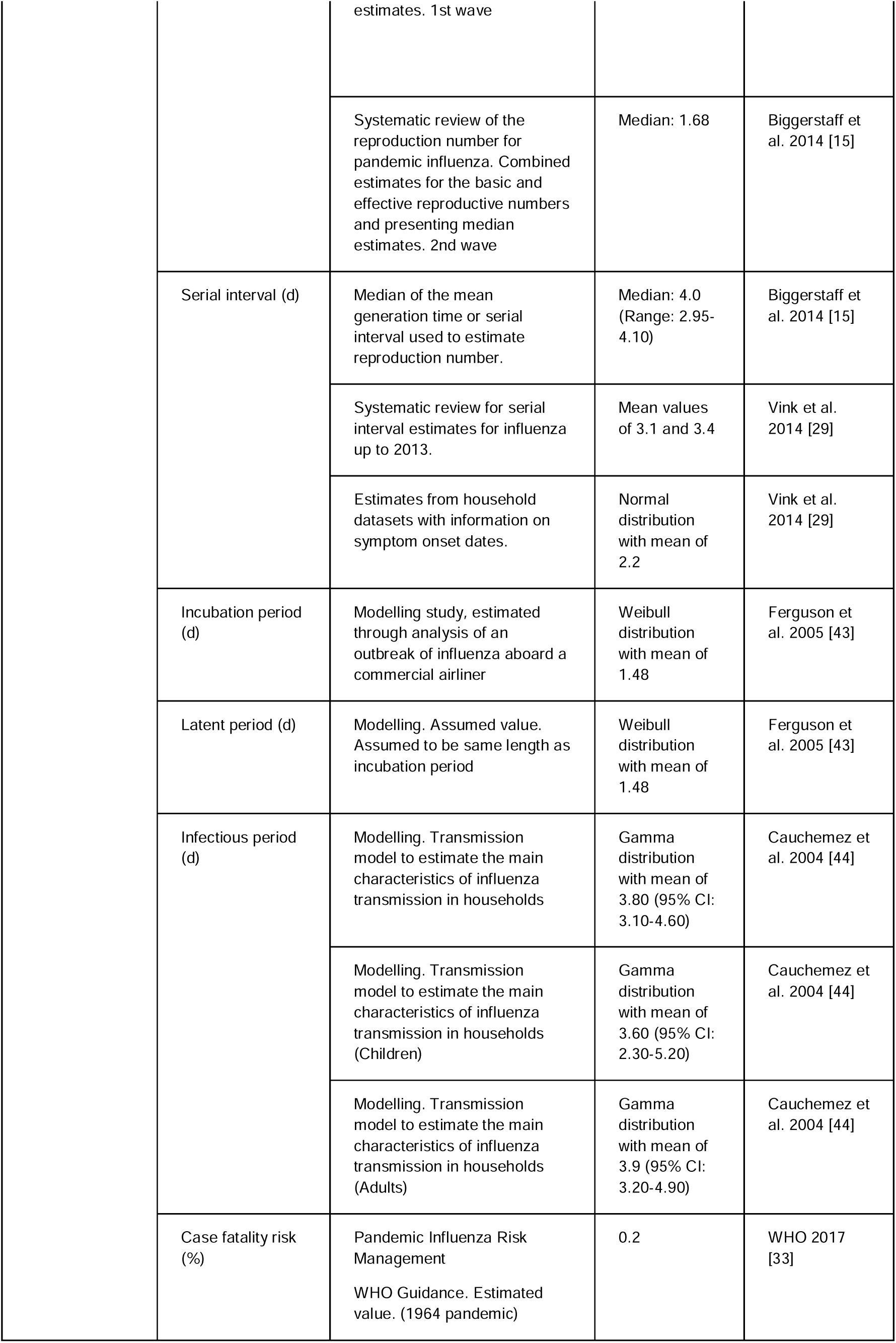
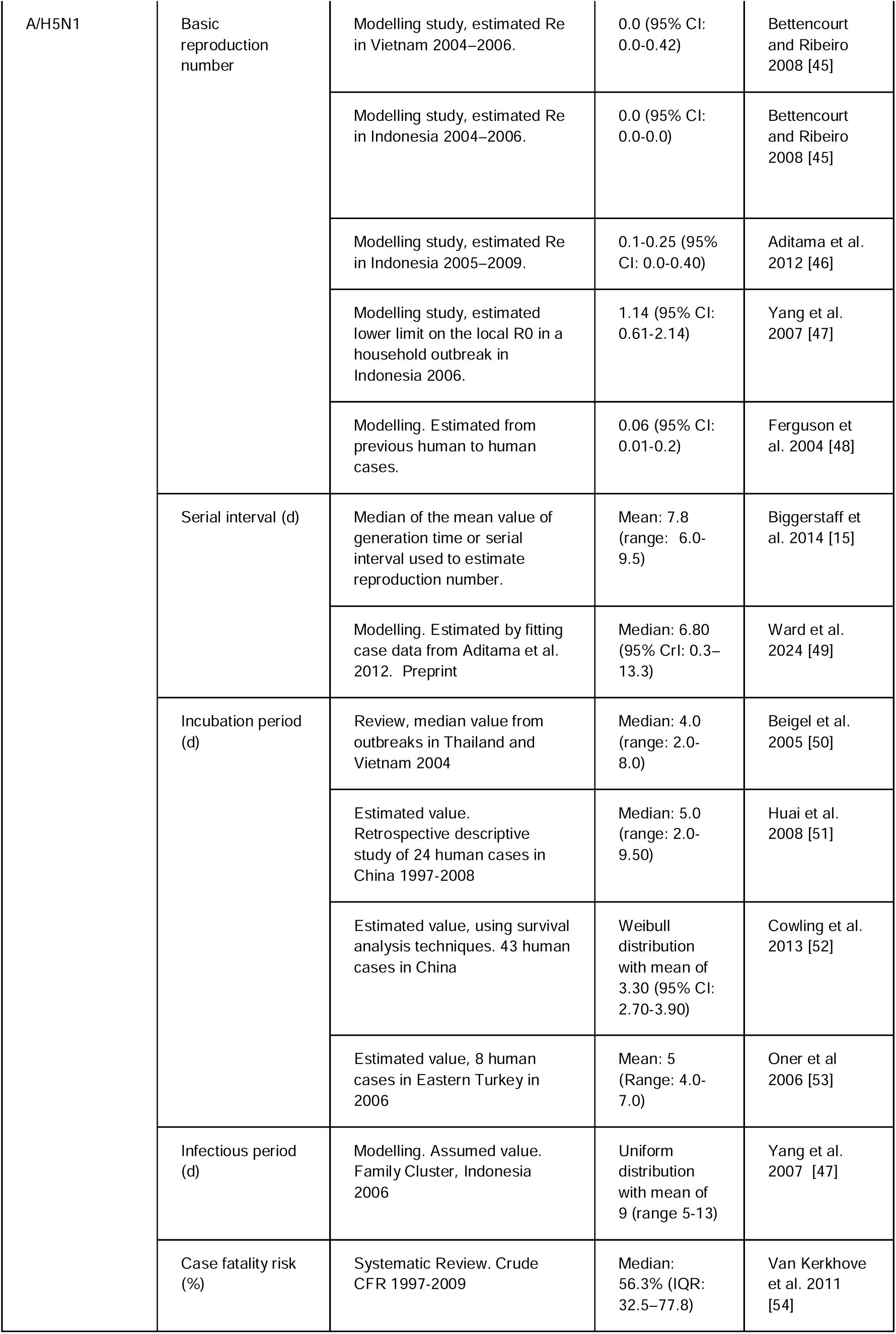
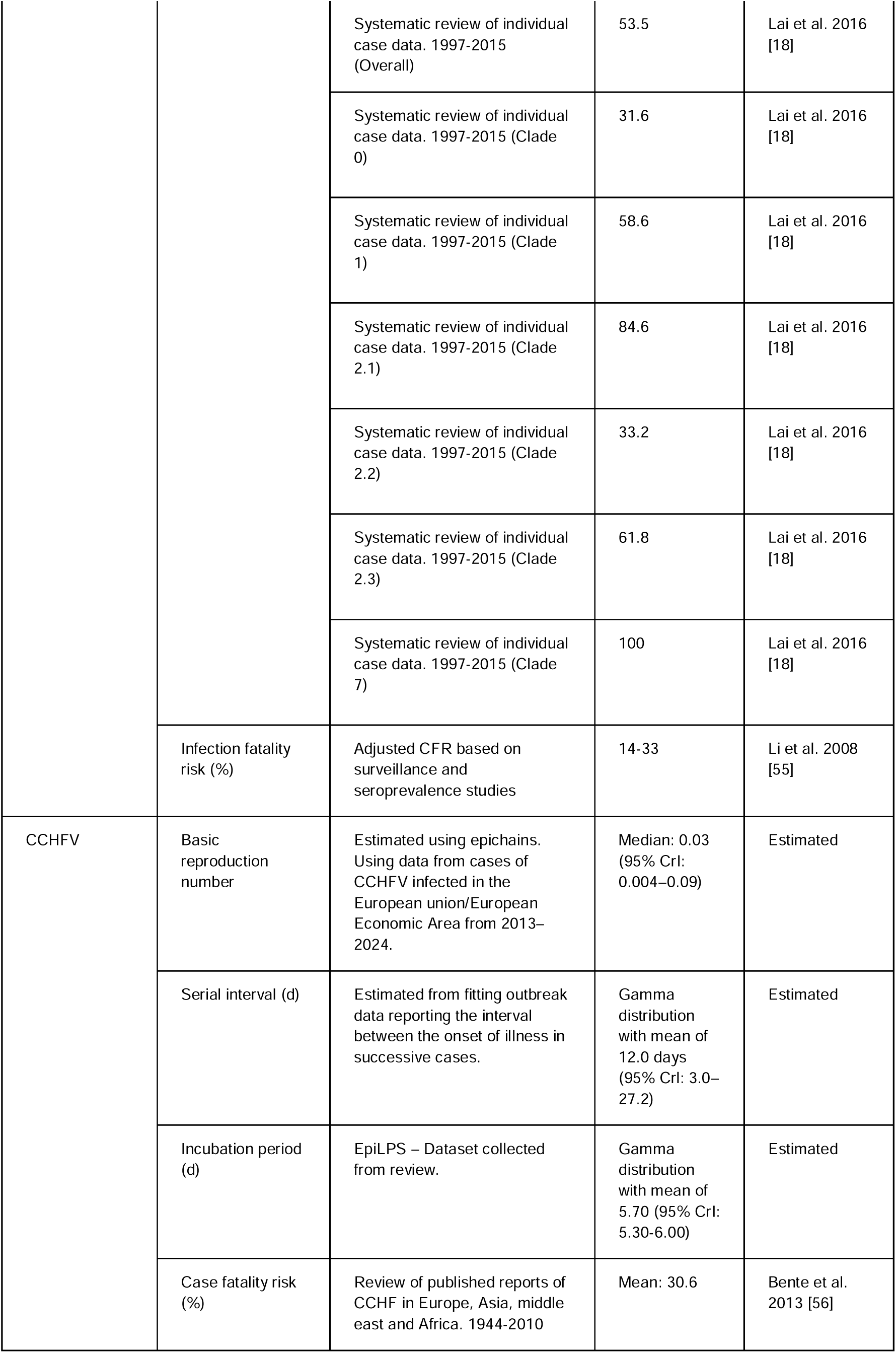
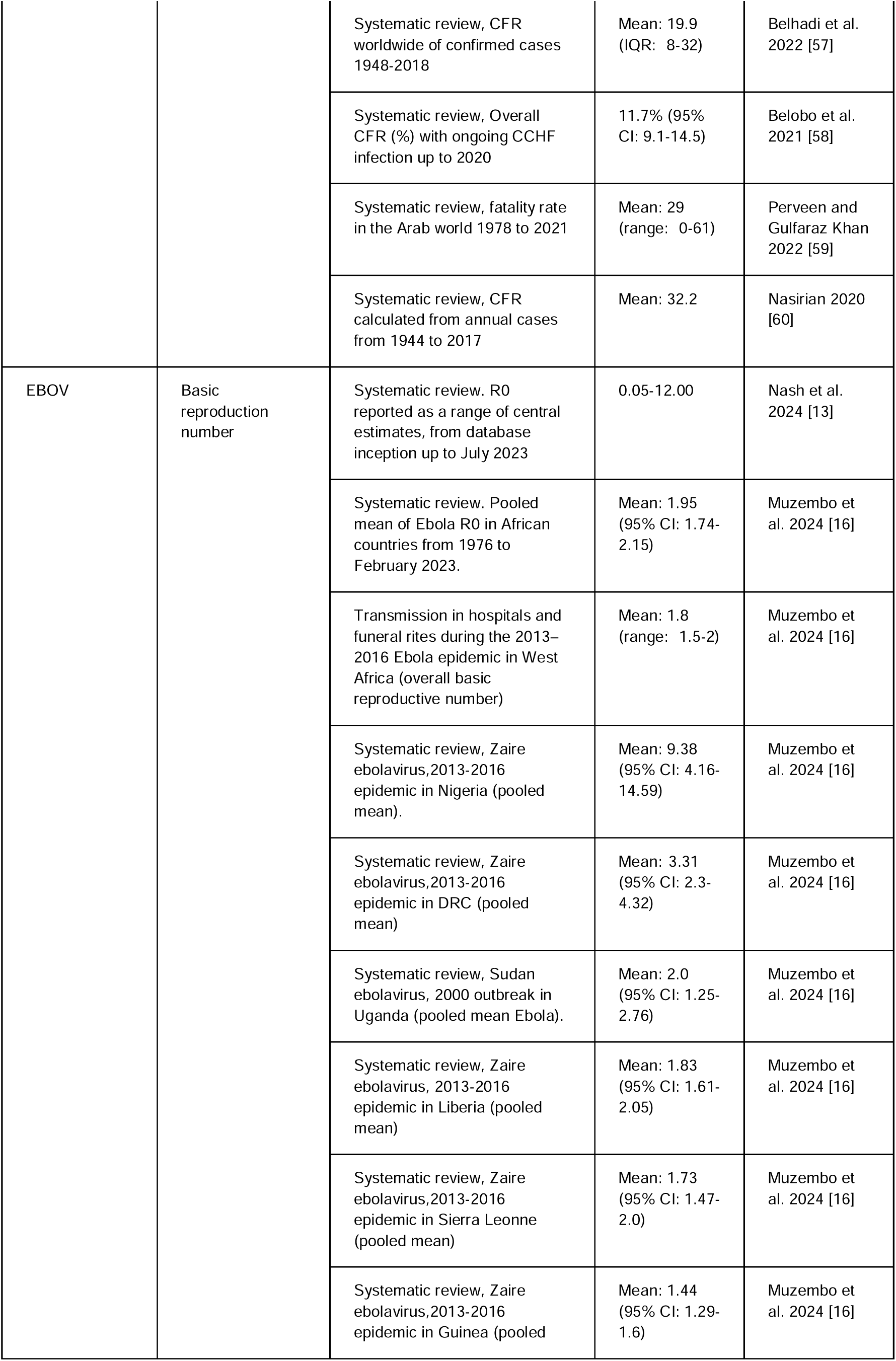
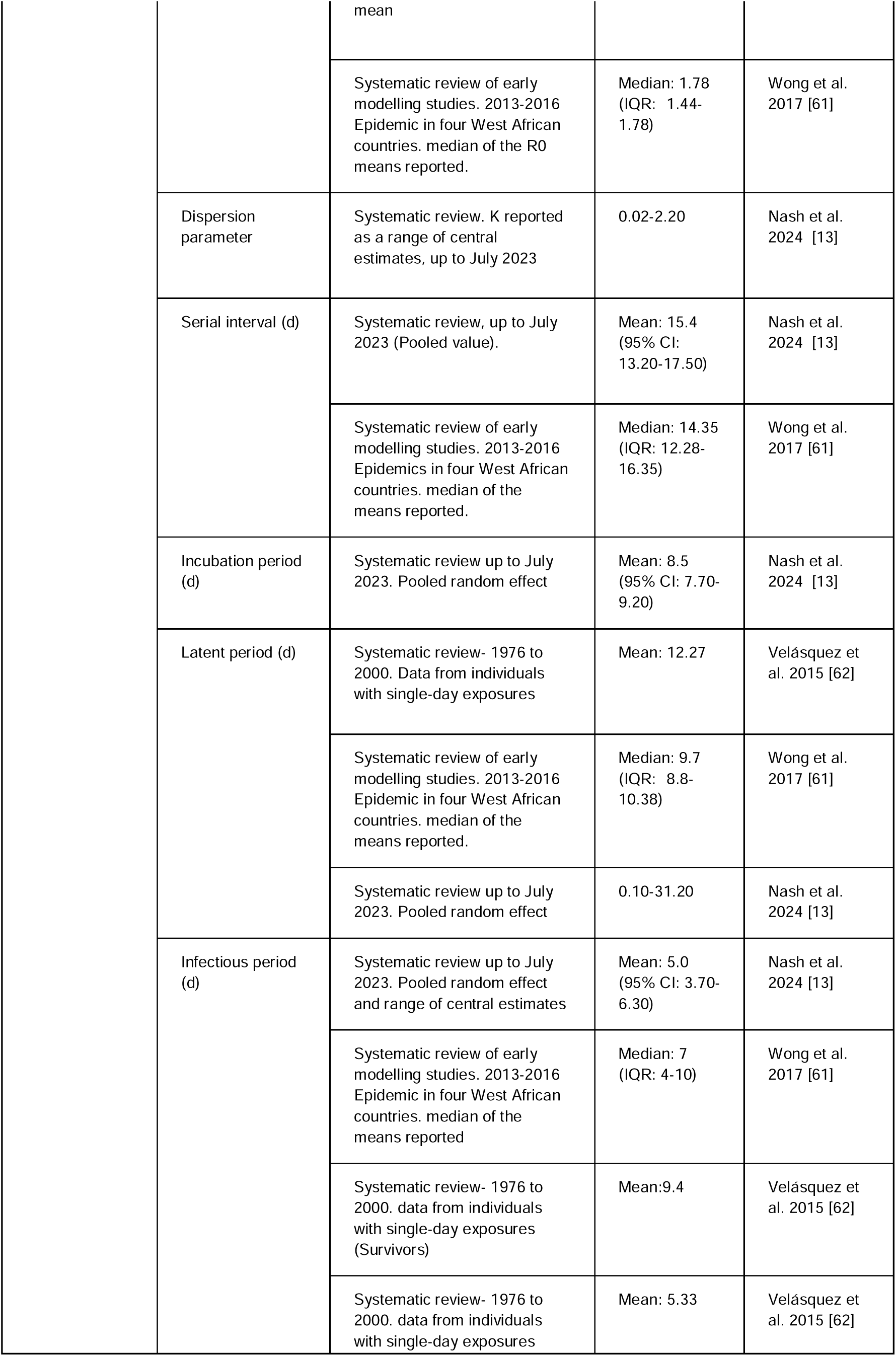
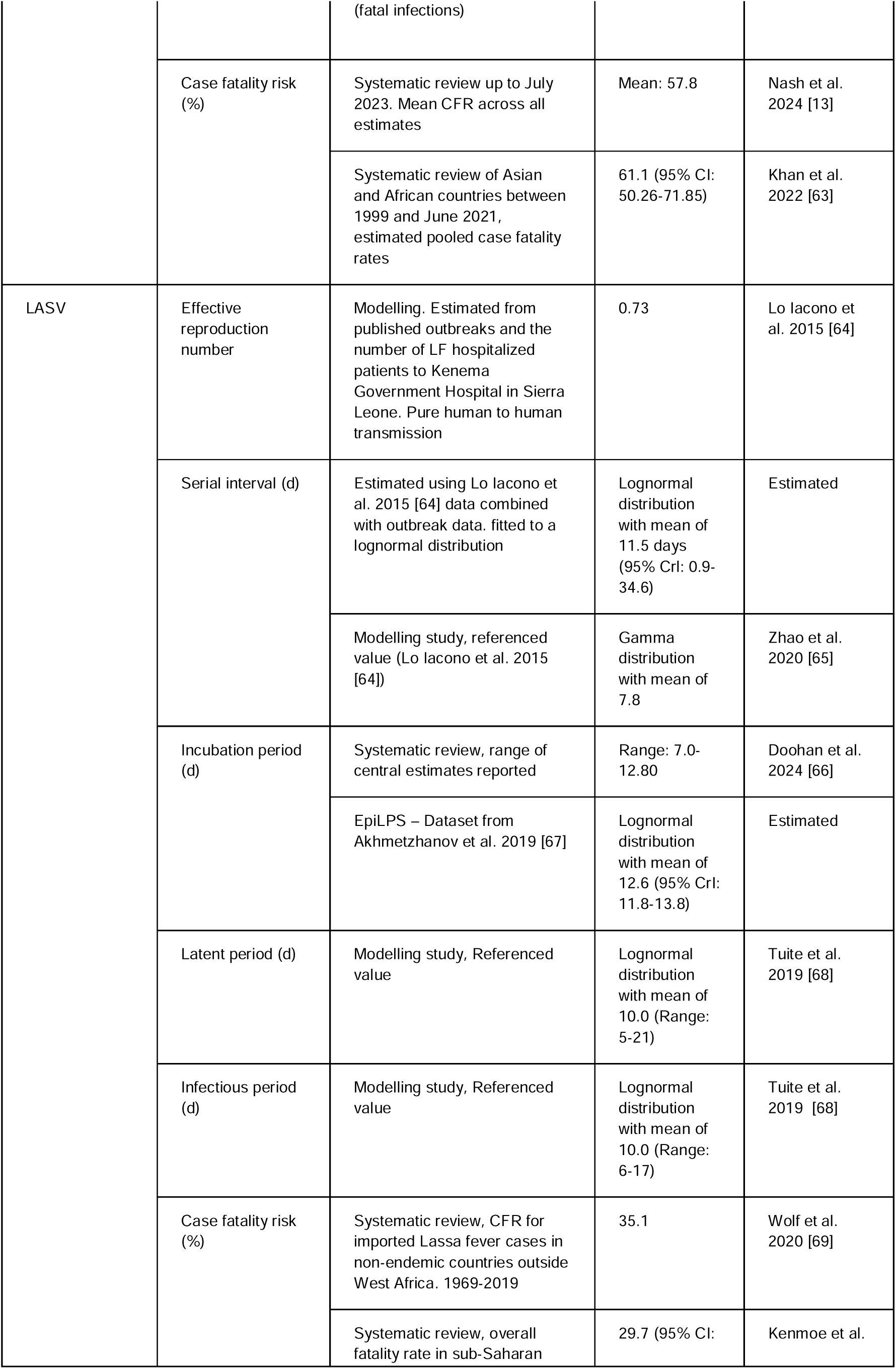
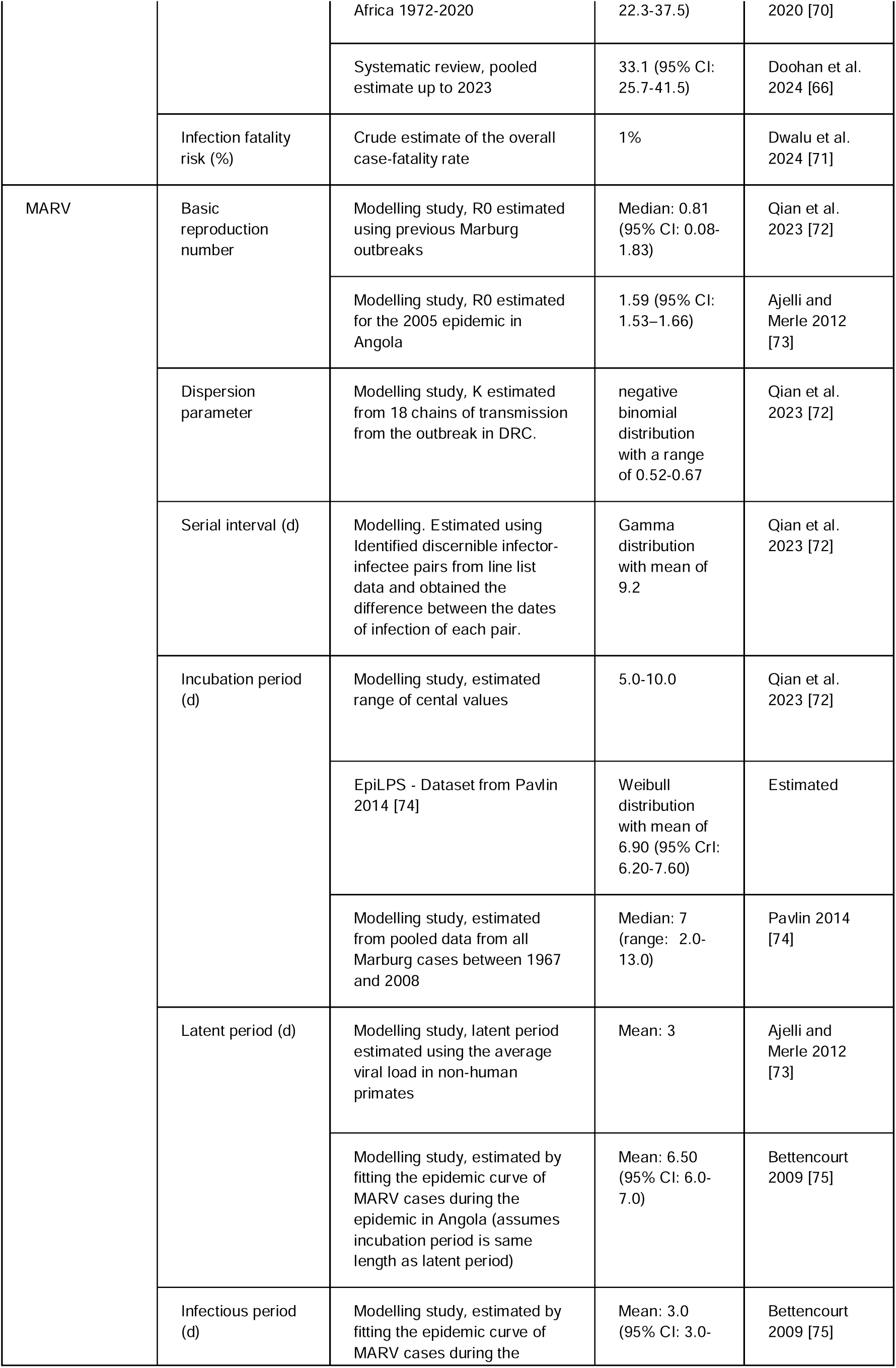
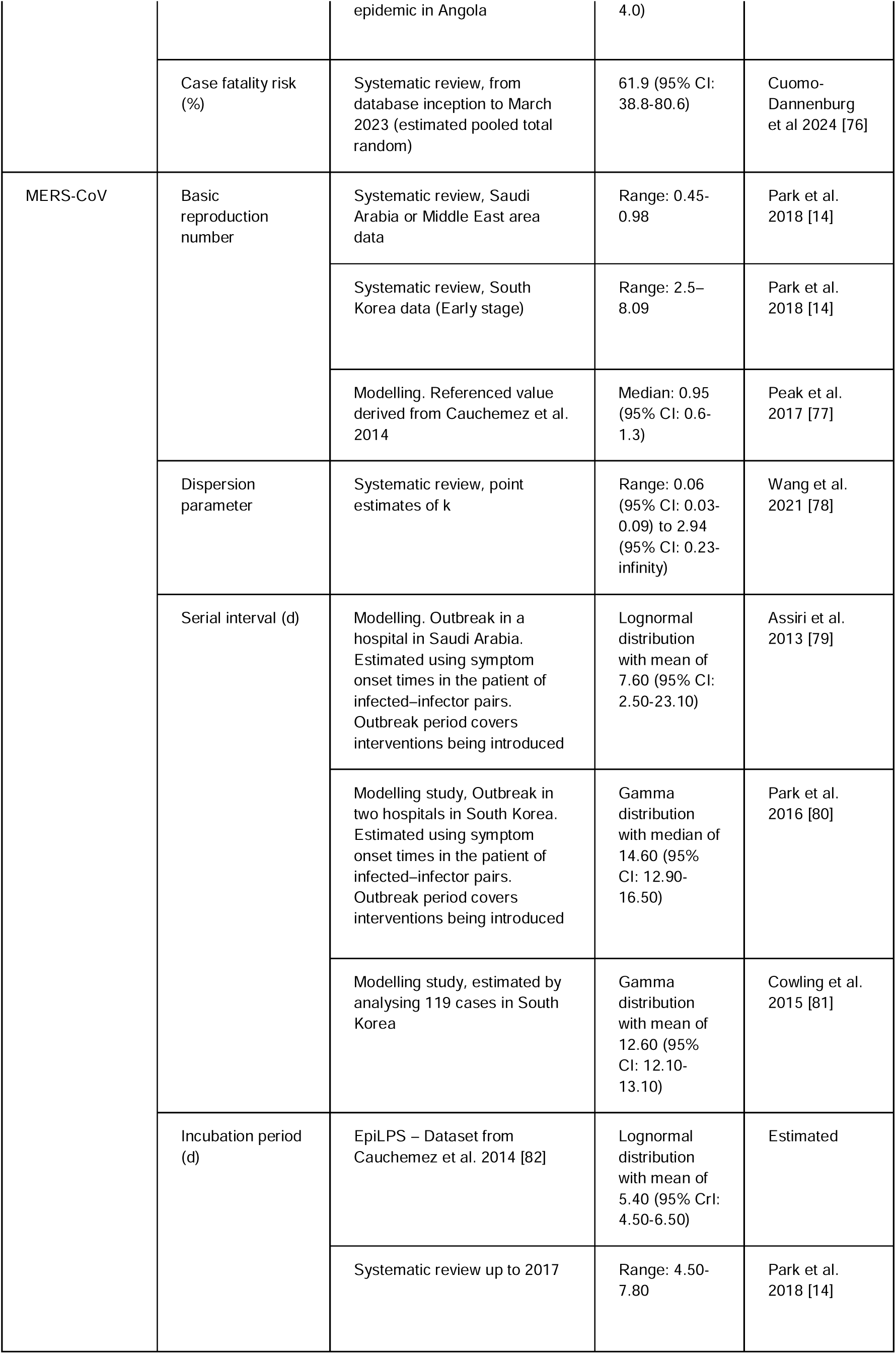
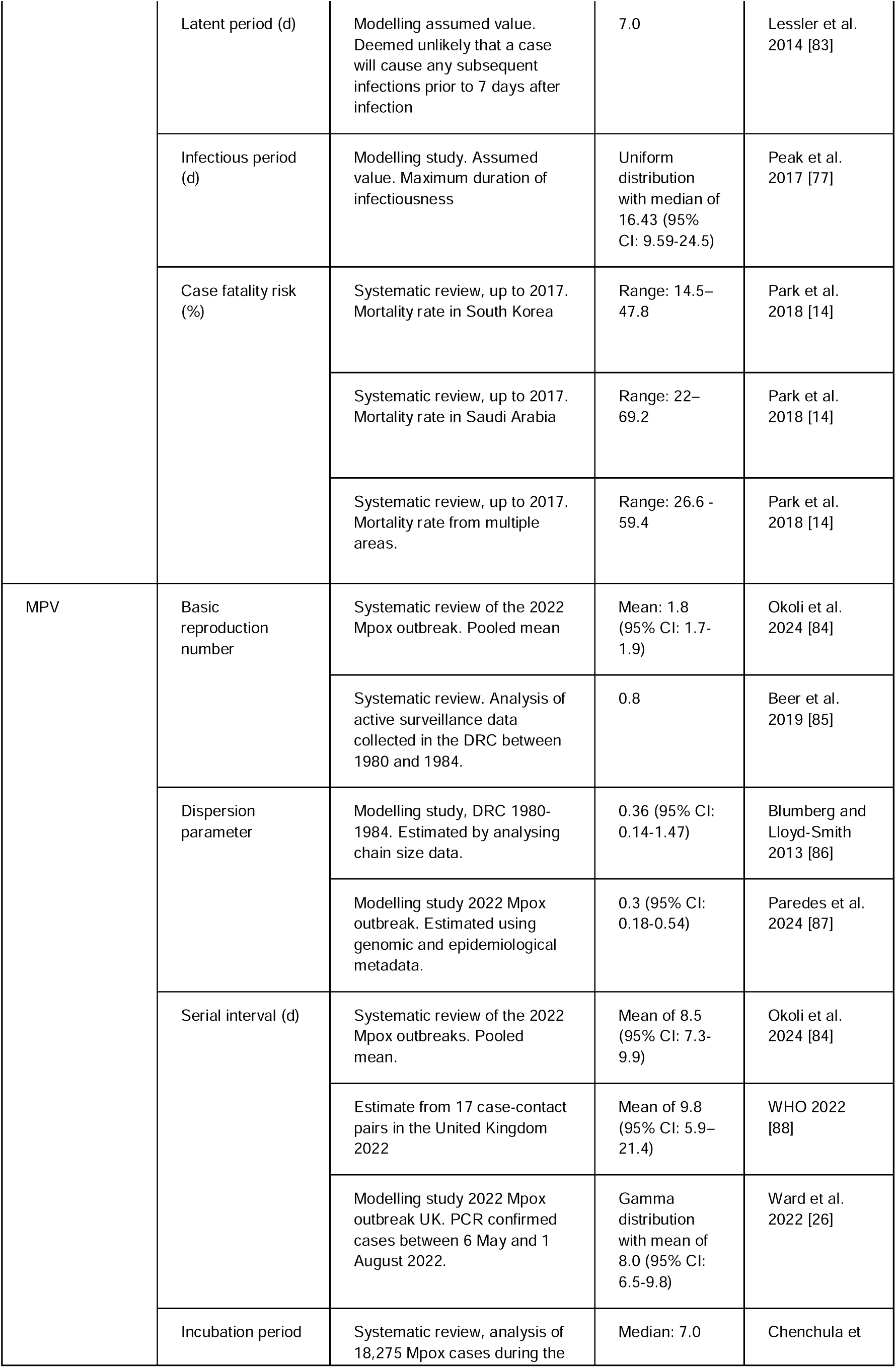
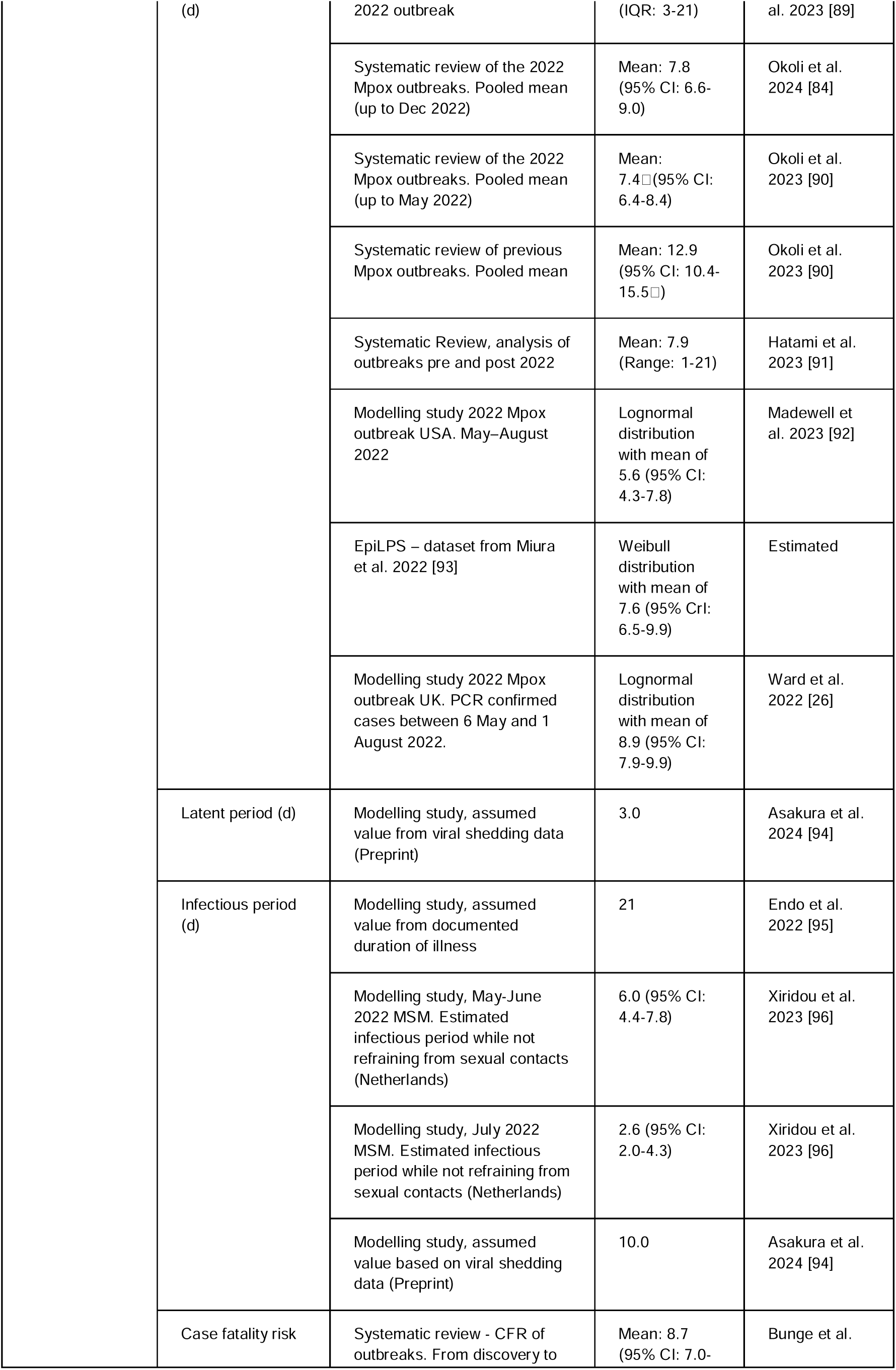
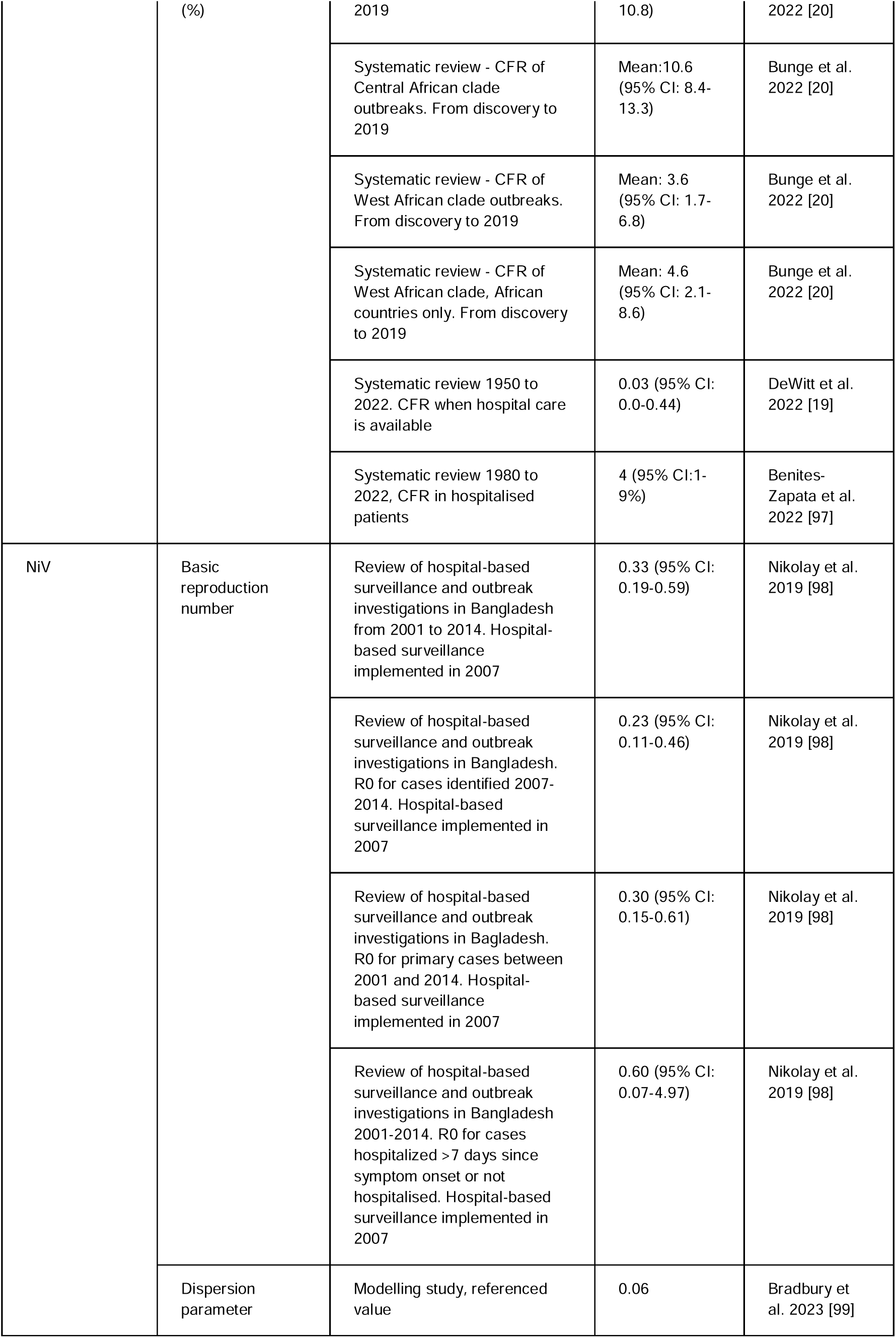
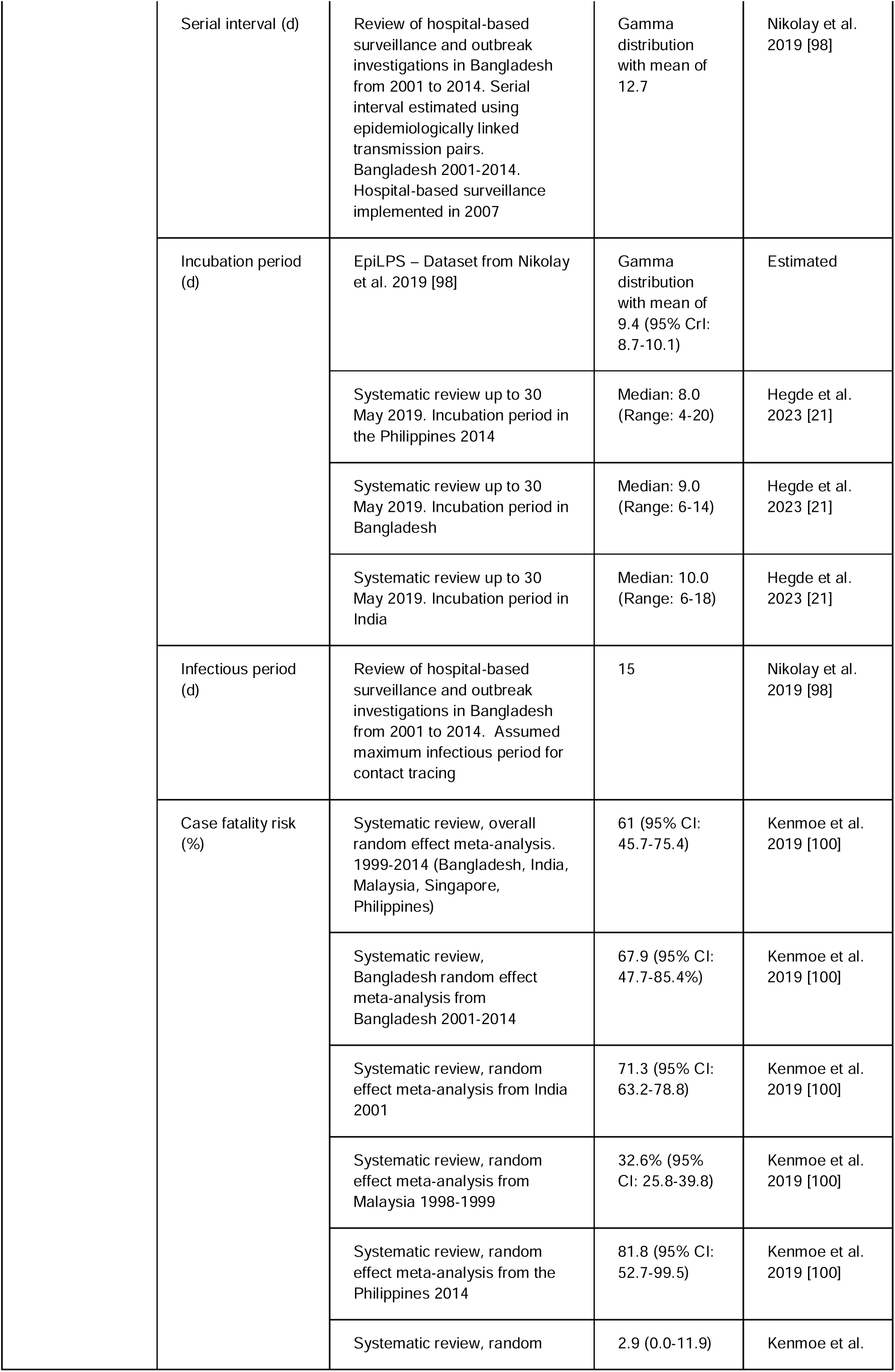
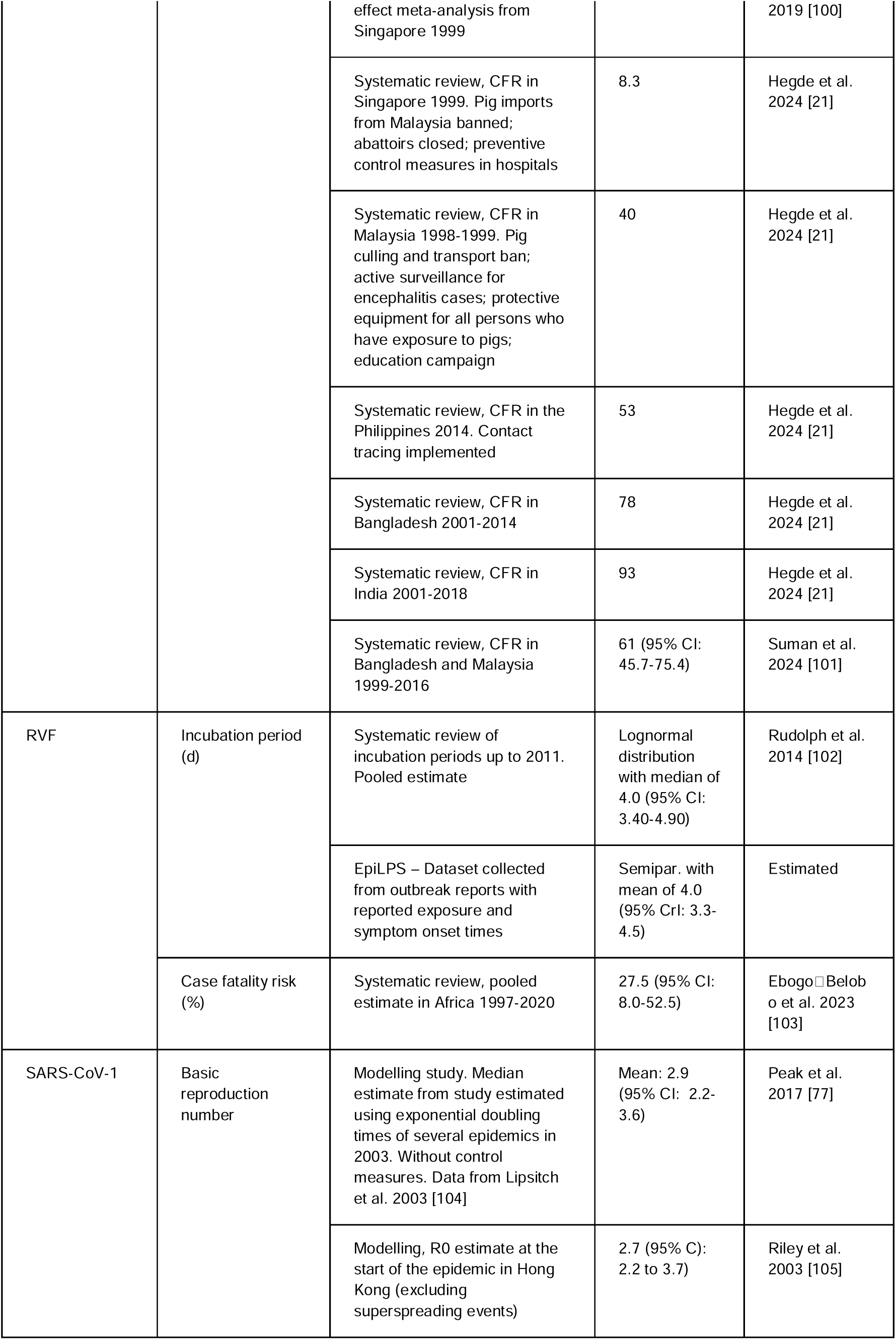
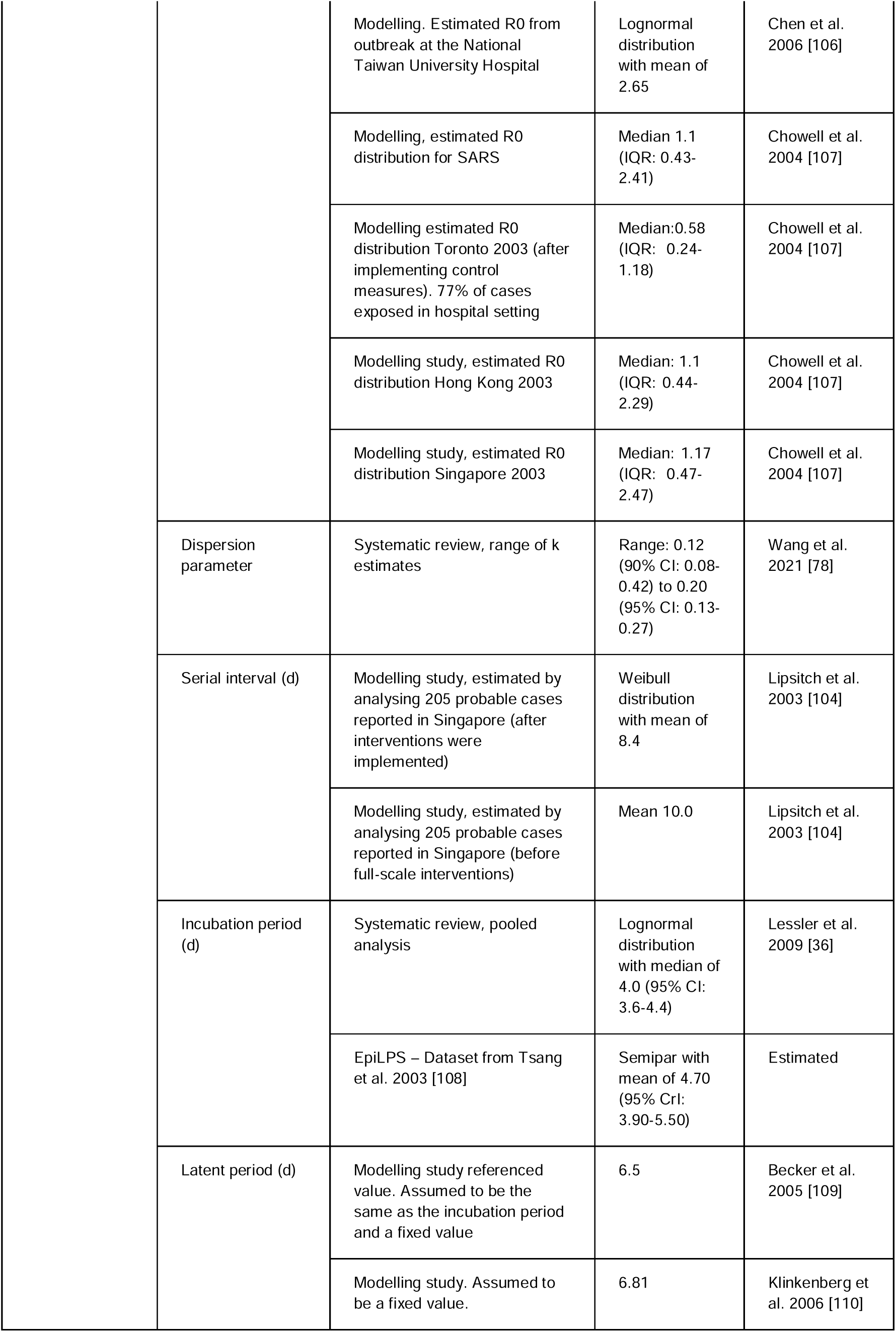
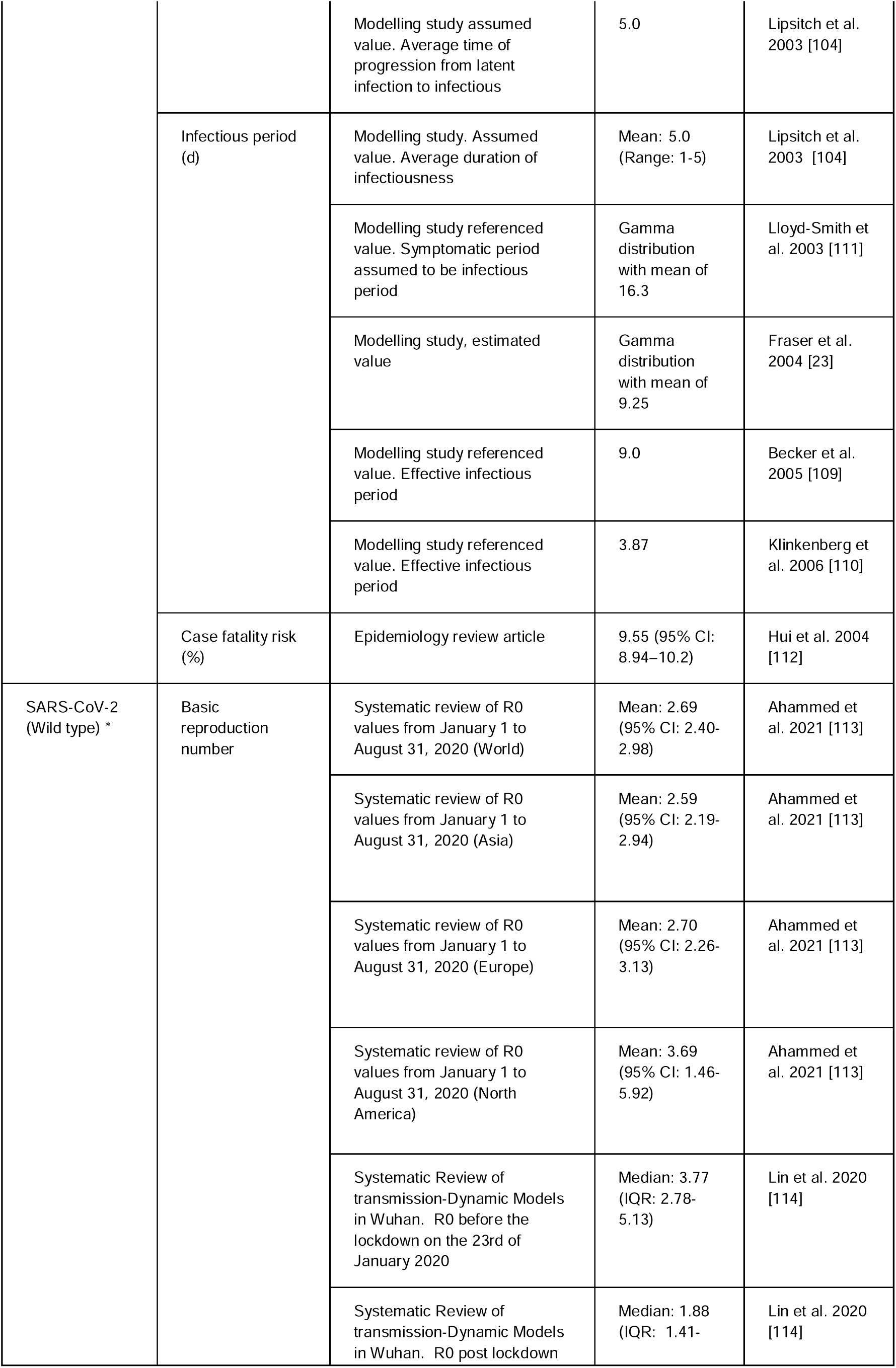
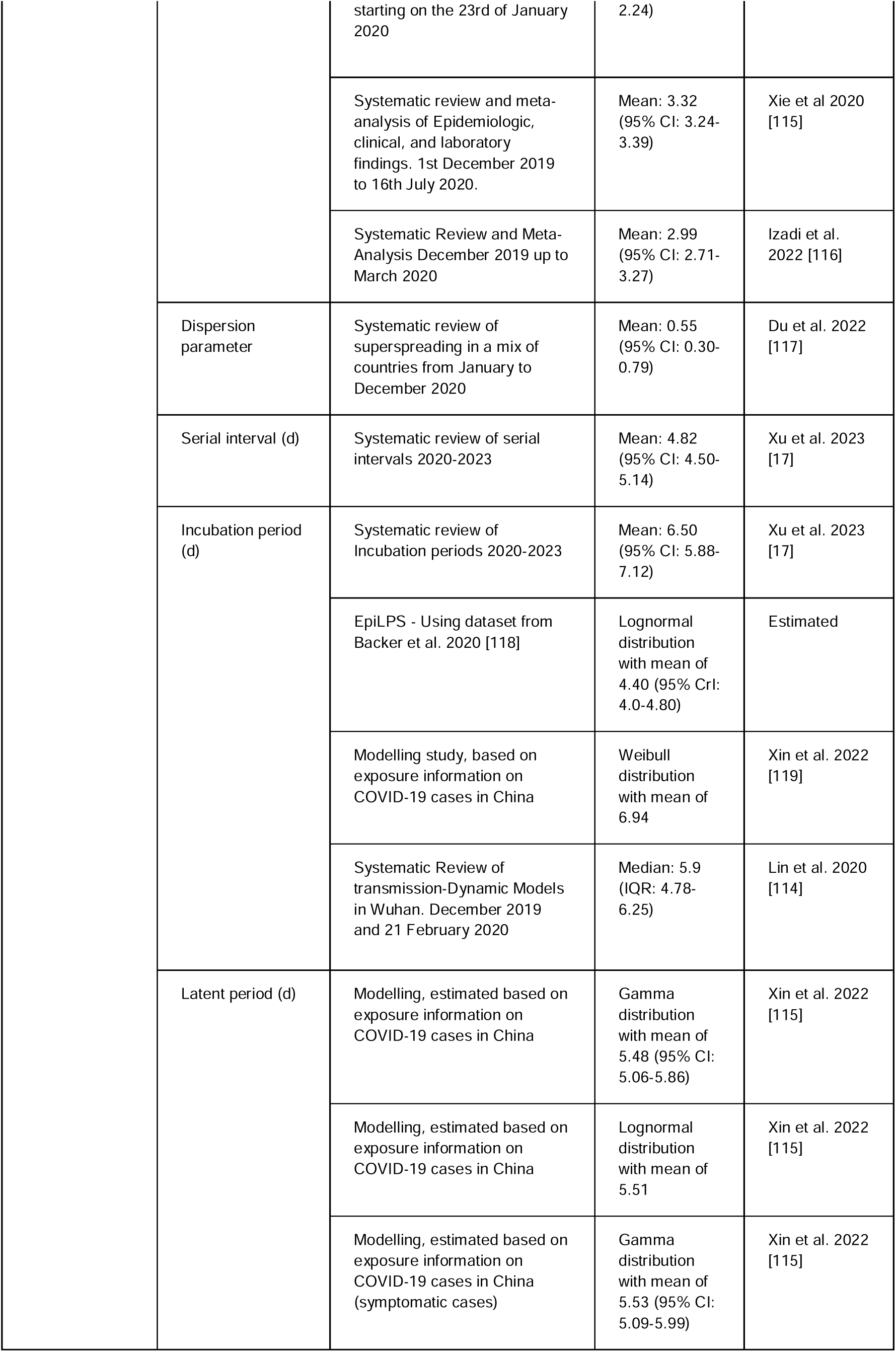
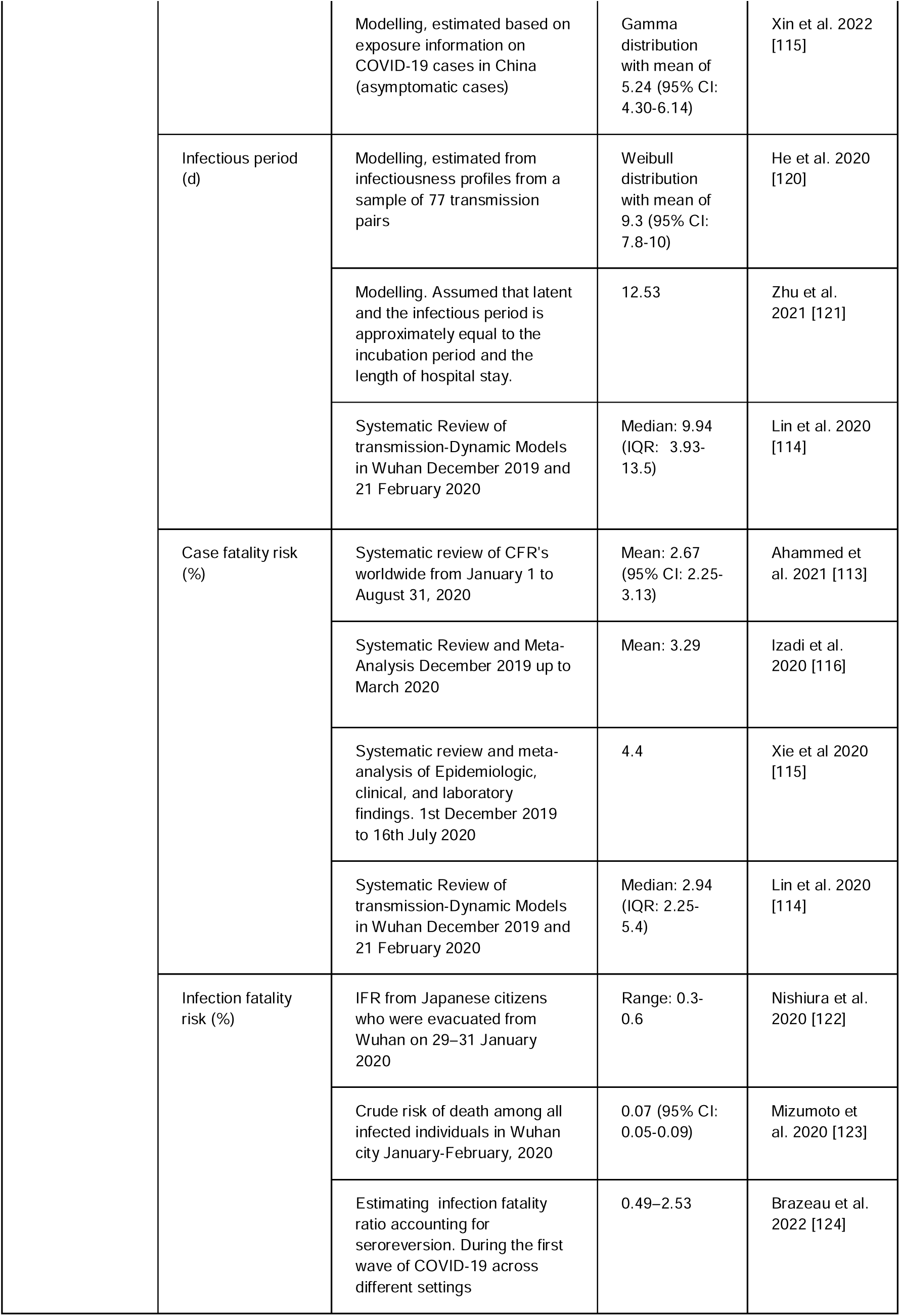
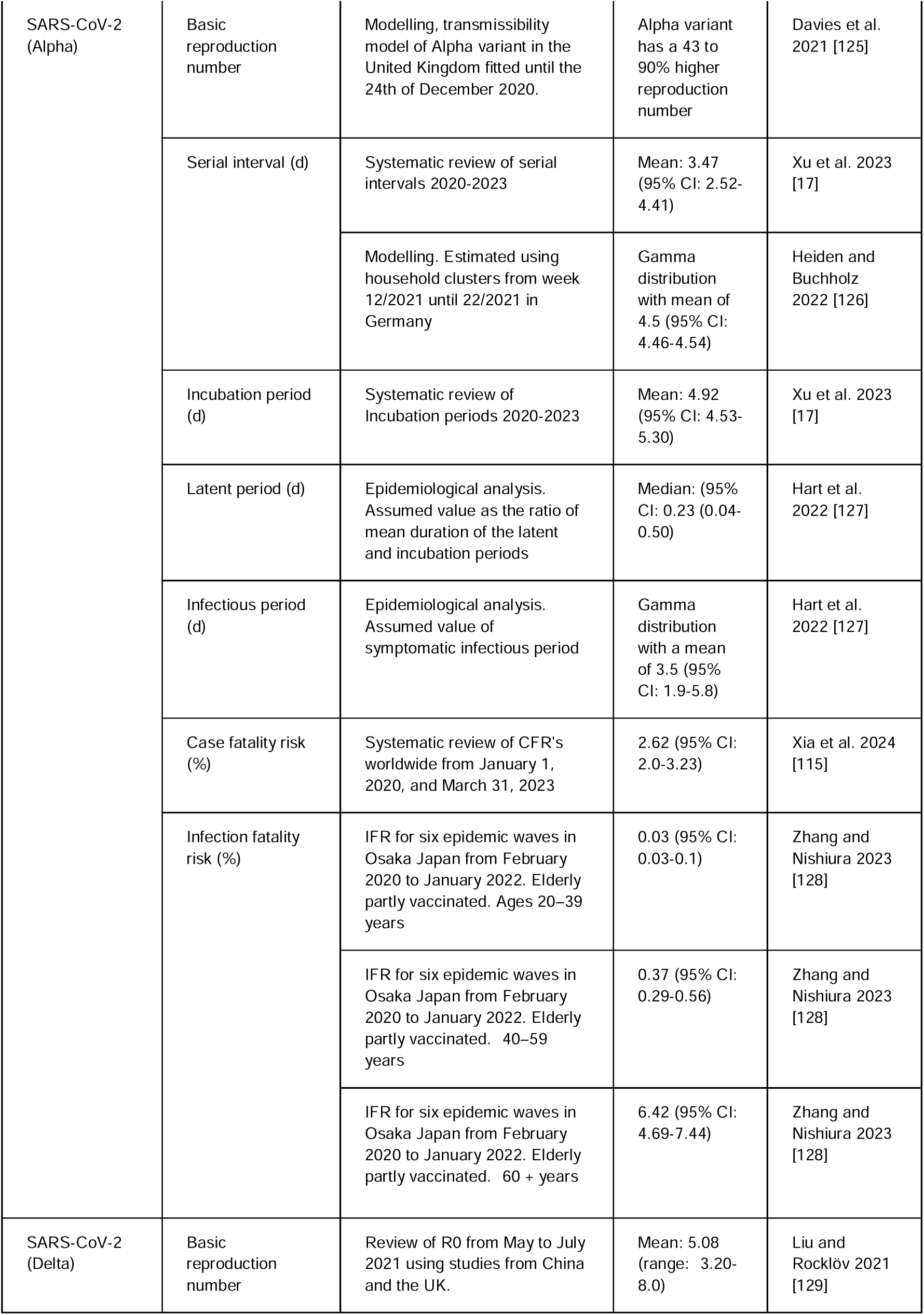
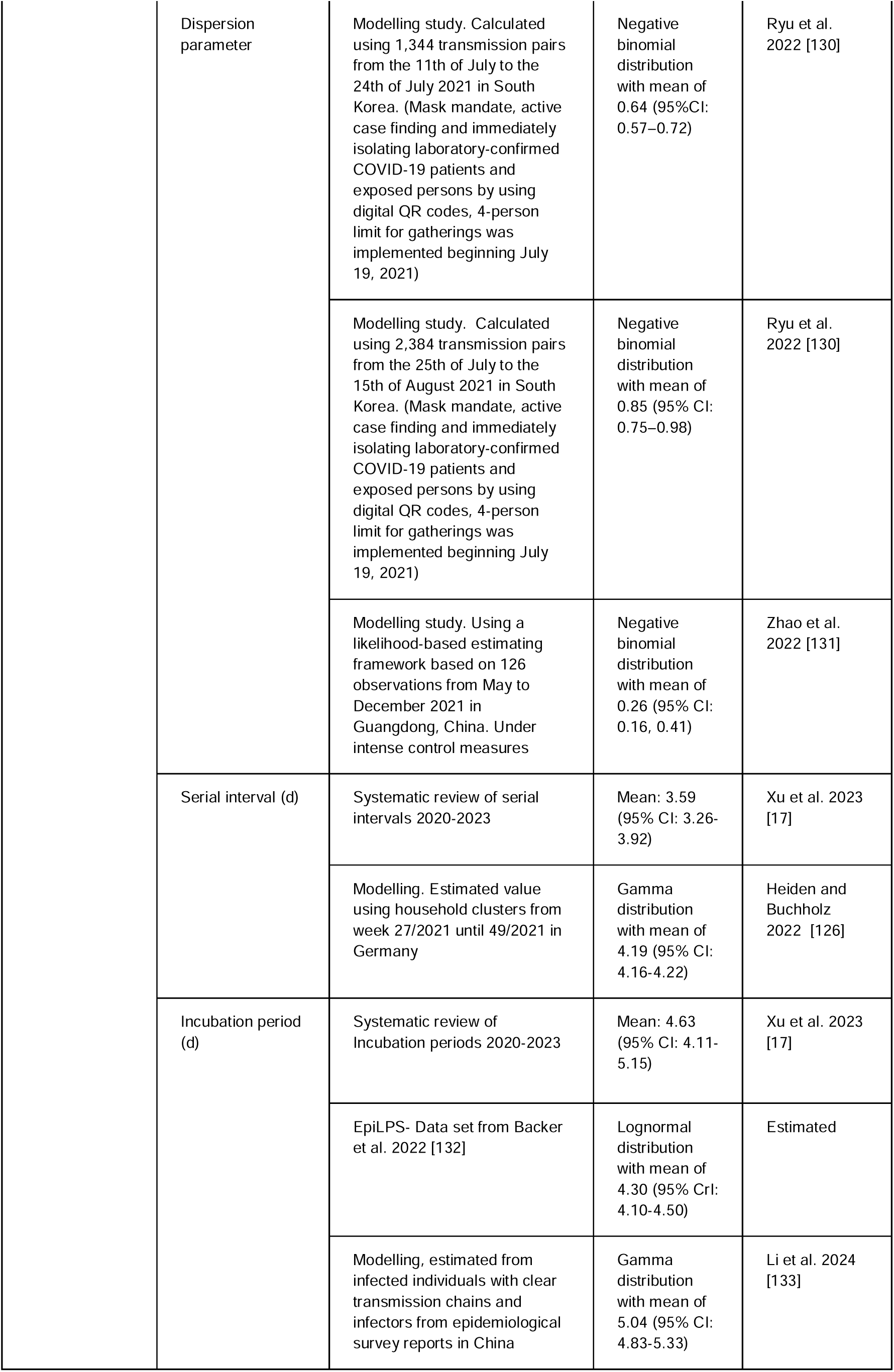
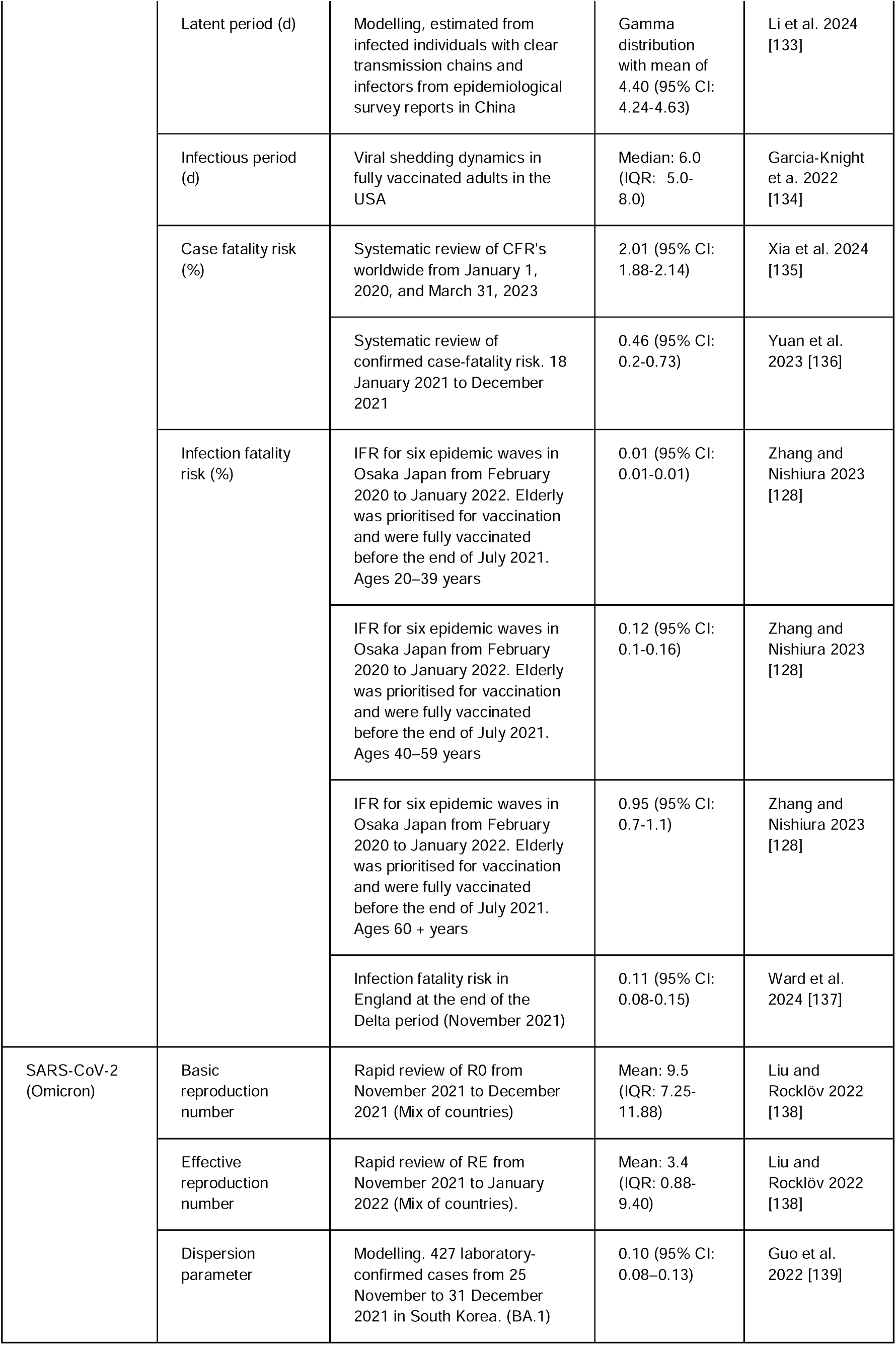
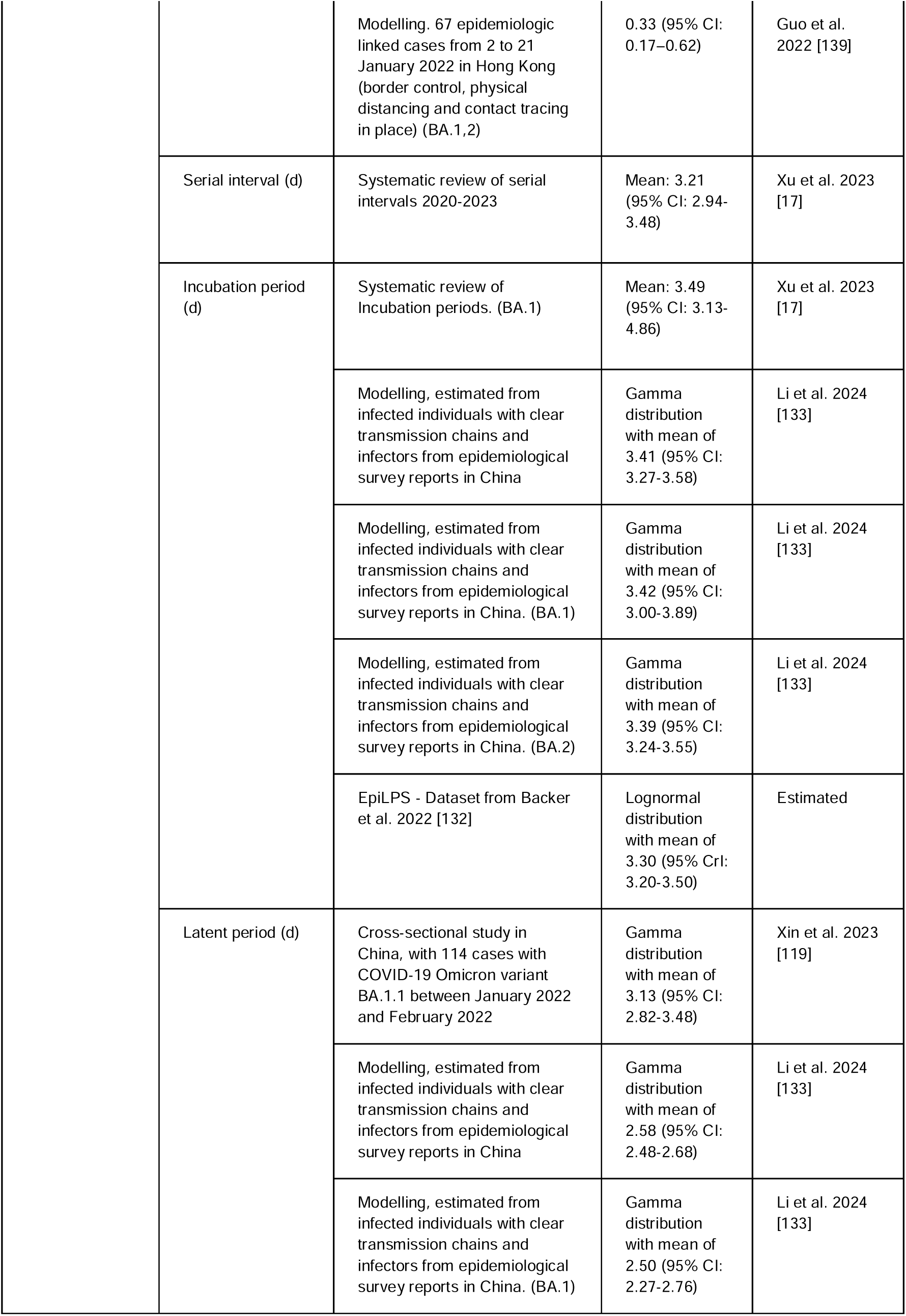
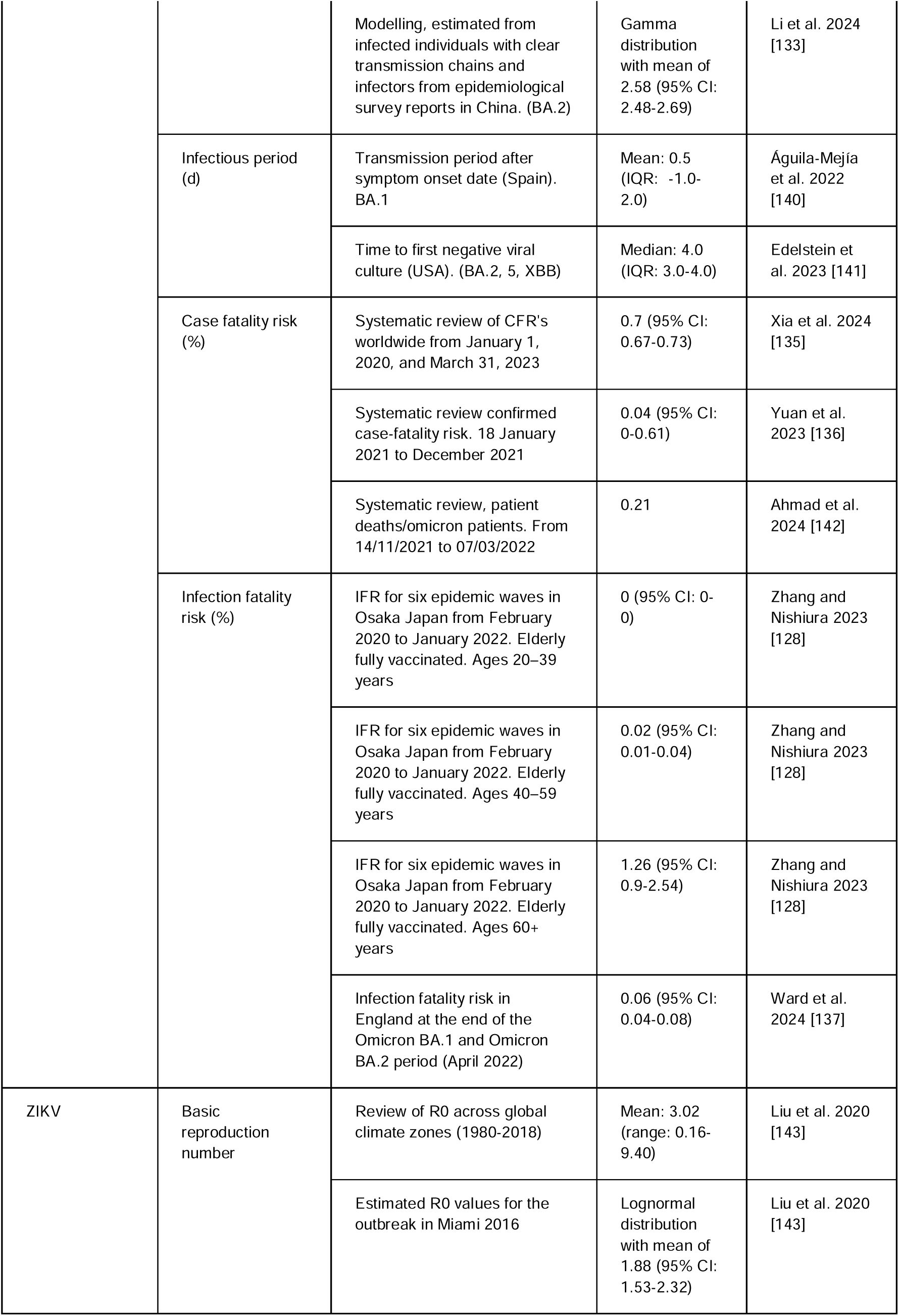
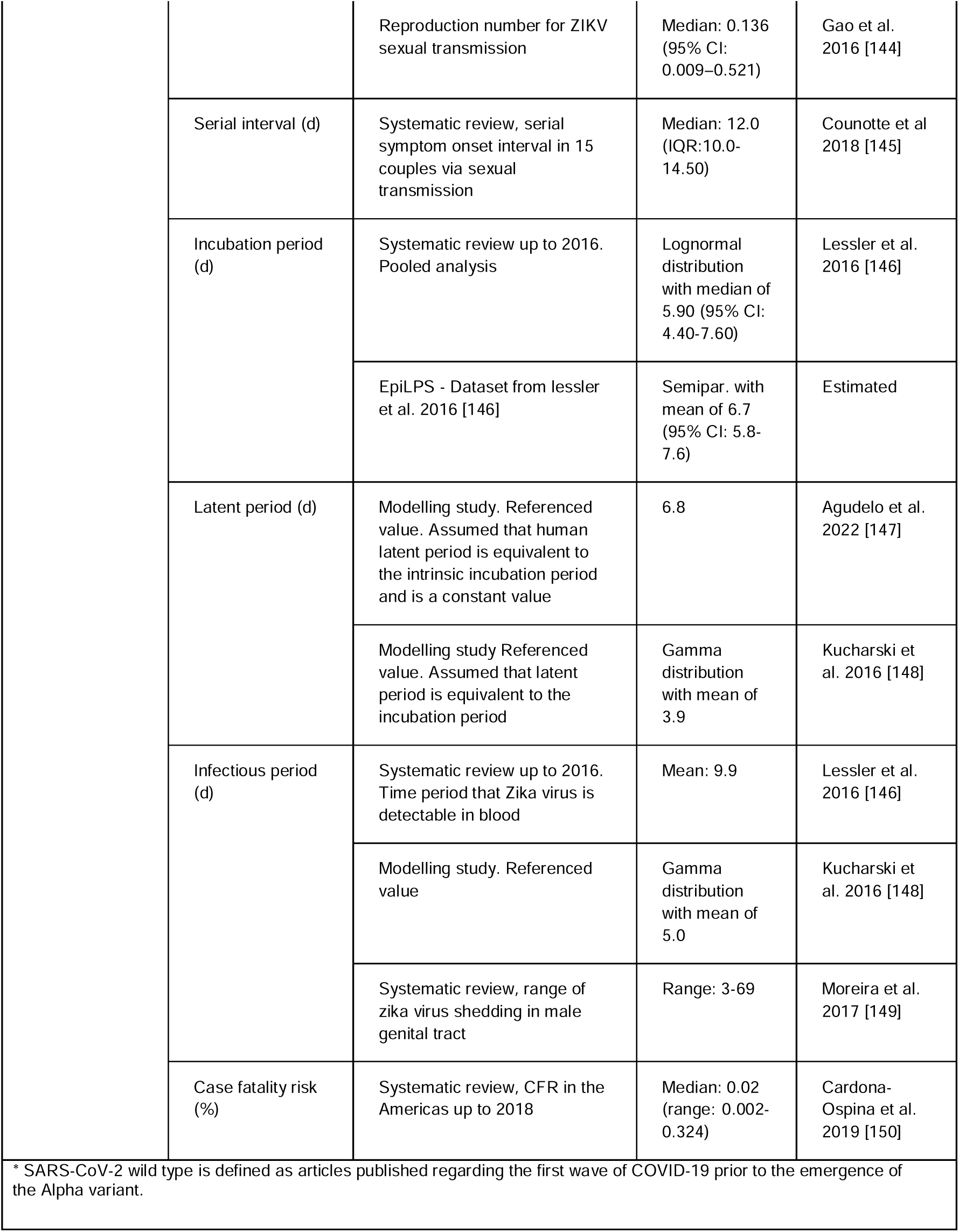
Epidemiological parameters of pandemic potential pathogens.

We performed a 5,000-iteration Monte Carlo simulation, using a non-parametric bootstrap-aggregation sampling method. In each of the Monte Carlo iterations, for a given pathogen and parameter, we resampled the available studies with replacement. A single random value was then drawn from each study’s distribution. The final parameter value for that iteration being the average of these draws. Transmission routes were encoded as fixed binary indicators (presence/absence) and were not subject to sampling.

Pathogens were clustered in two stages. First, for each Monte Carlo iteration, we performed an independent K-means clustering on the resulting pathogen parameter profiles. This yielded 5,000 plausible clustering solutions, each reflecting uncertainty in the underlying epidemiological estimates. Second, to obtain a single robust set of pathogen archetypes, we applied consensus clustering by constructing a co-assignment matrix representing the proportion of iterations in which each pair of pathogens clustered together. Hierarchical clustering of this matrix produced the final consensus dendrogram. The number of clusters was selected using silhouette width and epidemiological interpretability [12].

When presymptomatic transmission was included within the algorithm, we defined it as the probability that the serial interval is shorter than the incubation period, P (SI < IP). To estimate this for each pathogen, we compiled SI and IP distributions. Within each Monte Carlo iteration, one SI distribution and one IP distribution were selected from the study selection. Using these distributions, we simulated 5,000 paired SI–IP iterations. The final estimate for each iteration is the proportion of the 5,000 pairs in which the sampled SI was less than the sampled IP. For influenza A subtypes data on SI and IP distributions were pooled.

#### 2.3.4 Sensitivity analysis

We repeated the clustering using a restricted parameter set consisting R, SI, and CFR, with and without presymptomatic transmission. To determine how robust the pathogen classifications were to changes in the parameter space, and to evaluate the stability of pathogen groupings when the information content of the model was reduced.

To explore the flexibility and potential further application of our method, we performed an analysis on a widely available set of parameters: R, incubation period, and CFR. This enabled the incorporation of a more diverse range of pathogens, including those with different transmission dynamics, and to investigate how they fit within this classification. We extended this to include pathogens with had been excluded from the original analysis (RVFV), food and water borne pathogens (*Vibrio cholerae* and Norovirus), viruses common in paediatrics (Measles virus, Enterovirus A71 and Human metapneumovirus), bioterrorism related pathogens (*Yersinia pestis*, Variola virus and *Bacillus anthracis*), other vector borne pathogens (SFTS virus and Chikungunya), and a retrovirus (Human immunodeficiency virus) (**Supplementary Table S3**).

## 3. Results

### 3.1 Parameter review

A total of 154 articles were retrieved (**Supplementary Figure S1**). This included 43 articles obtained through the initial search of systematic reviews and 69 articles obtained from supplementary sources. Among these, one was a grey literature report published by the WHO. We extracted 302 parameter estimates from the articles identified (**Figure 1**). 28 articles were identified that provided sufficient data to estimate the incubation period with the {EpiLPS} package. Additionally, 14 articles contributed pre-existing datasets that were incorporated into the analysis.

**Figure 1.**
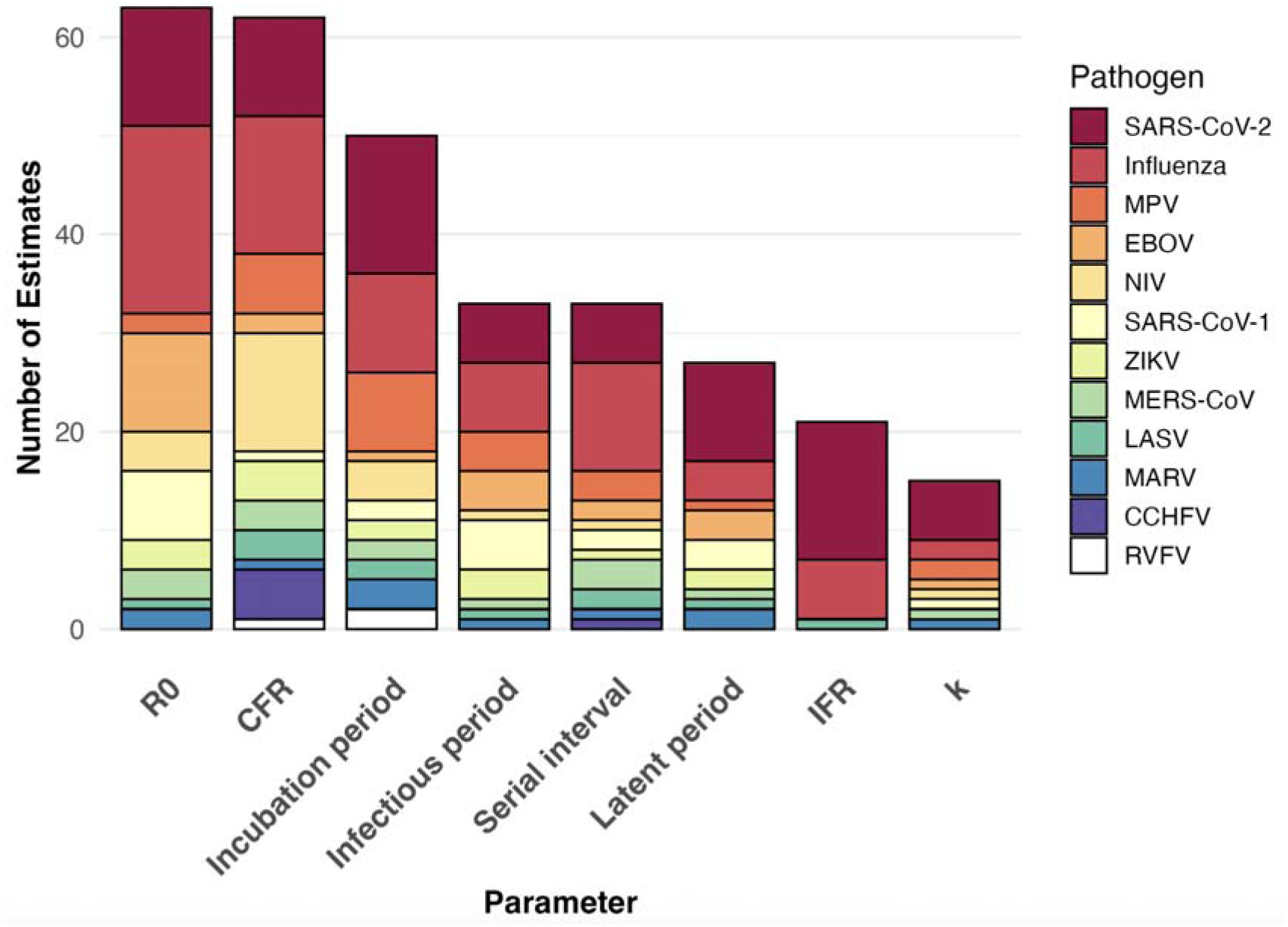
Distribution of parameter estimates from the literature search

From 154 articles we extracted 302 parameter estimates. 63 for the reproduction number, 15 for the dispersion parameter, 51 for the incubation period, 33 for the serial interval, 27 for the latent period, 33 for the infectious period, 59 for the CFR, and 21 for the IFR.

### 3.2 Epidemiological characteristics

#### 3.2.1 Transmission dynamics

We extracted 63 estimates for the reproduction number, which varied across pathogens and outbreaks due to differences in settings and the implementation of control measures (**Table 1**). One additional estimate was derived, with the median RL for CCHFV estimated at 0.03 (95% CrI: 0.004–0.09) based on cases reported in the European Union between 2013 and 2014 (**Supplementary figure S4**).

A total of 15 estimates for the dispersion parameter (k) were extracted, revealing varying degrees of heterogeneity in transmission. SARS-CoV-1, SARS-CoV-2, MERS-CoV and NiV exhibited particularly low k values, indicating a high potential for superspreading events, whereas influenza displayed greater uniformity in transmission. Pathogens such as EBOV were reported to have a wide range of estimates suggesting the degree of superspreading may be dependent on outbreak setting (**Table 1**).

#### 3.2.2 Time to key events

We extracted 51 estimates for the incubation period and provided 13 additional estimates based on publicly available data. The length of the incubation period varied across pathogens, with pandemic influenza exhibiting the shortest incubation period. In contrast, pathogens primarily transmitted through contact with infected body fluids, such as LASV, EBOV, and CCHF, had the longest incubation periods (**Table 1**).

For the serial interval, we extracted 33 estimates. As with the incubation period, serial intervals were shortest for pandemic influenza and SARS-CoV-2 and longest for contact-transmitted pathogens (**Table 1**). Additionally, we estimated serial intervals for LASV and CCHF, with mean values of 11.5 days (95% CrI: 0.9–34.6) (**Supplementary Figure S2**) and 12.0 days (95% CrI: 3.0–27.2) (**Supplementary Figure S3**), respectively.

For the latent and infectious periods, we extracted 27 and 33 estimates, respectively. These durations showed notable variation between pathogens, highlighting differences in disease progression (**Table 1**).

#### 3.2.3 Severity and Mortality Risk

We extracted 59 estimates for the case fatality rate (CFR) and 21 for the infection fatality rate (IFR). CFR estimates exhibited a broad range across pathogens, reflecting varying levels of disease severity (**Table 1**). Additionally, CFR varied within pathogens, with substantial heterogeneity between studies, outbreaks and age groups. For instance, NiV outbreaks with implemented control measures reported lower CFR estimates (**Table 1**).

Compared to CFR, fewer IFR estimates were reported. IFR values were consistently lower than CFR estimates for the same pathogen and demonstrated age-dependent variation (**Table 1**) Table 1. for Epidemiological parameters of pandemic potential pathogens. Located after conclusion

### 3.3 Ensemble clustering

We evaluated consensus clustering under the full parameter set, including and excluding the proportion of presymptomatic transmission (Figure 2 **& Supplementary Figure S9**). Although presymptomatic transmission estimates were generated for all pathogens, these values were highly uncertain, in some cases extending beyond realistic bounds (**Supplementary Table S2**). For this reason, and because their inclusion did not materially improve cluster stability, the clustering solution excluding presymptomatic transmission (Figure 2) was retained as the primary group.

**Figure 2.**
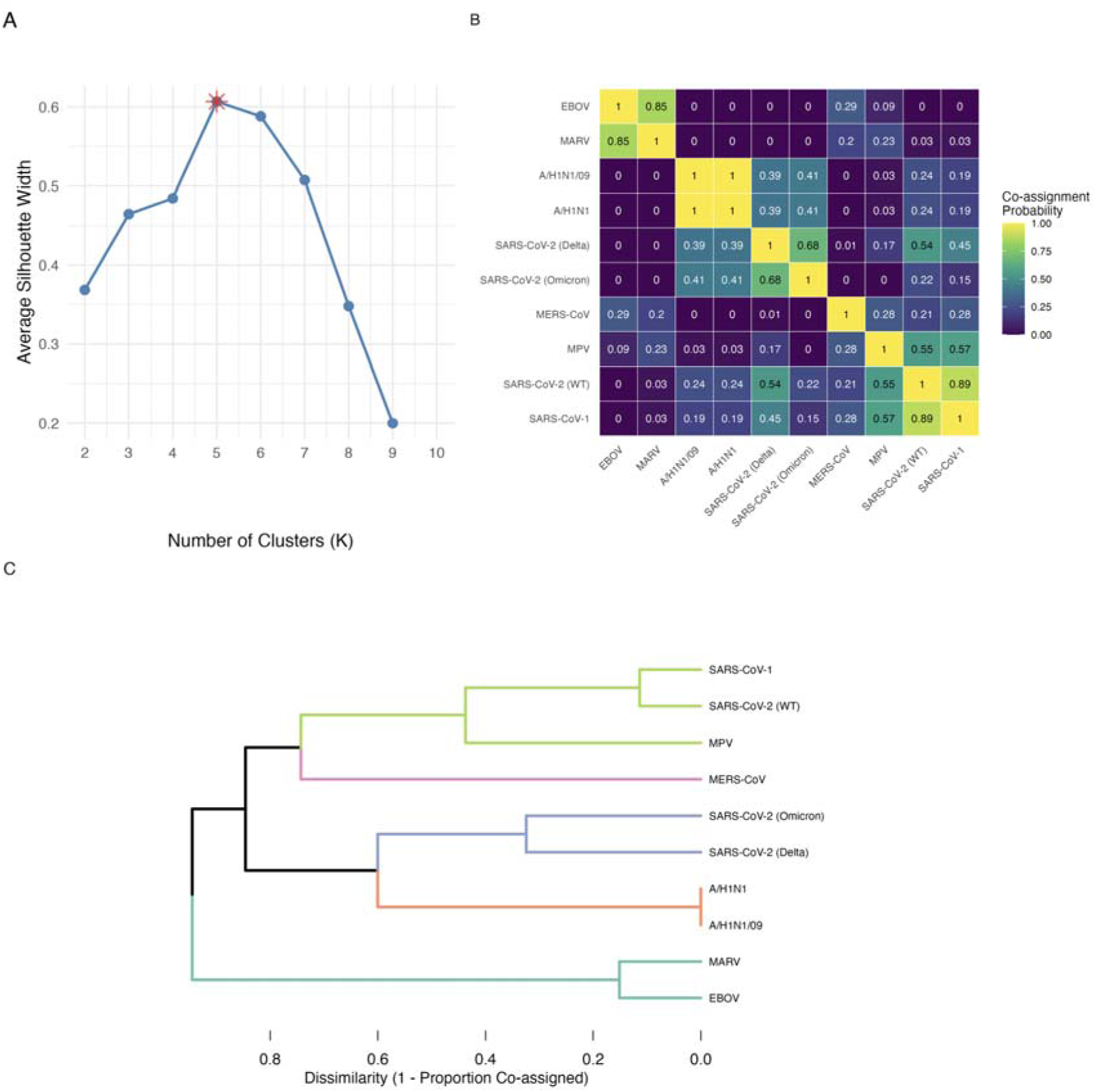
Hierarchical clustering with R_0_, serial interval, *k*, incubation period, latent and infectious period, case fatality risk and transmission route. A) The optimal number of consensus clusters determined by maximizing the average silhouette width. The optimal number of clusters is identified as K=5 indicated by the red asterisk. B) The co-assignment matrix, visualised as a heatmap, displays the proportion of the 5,000 Monte Carlo iterations in which each pair of pathogens was assigned to the same cluster.

The silhouette analysis (Figure 2A) suggested that five clusters maximised average silhouette width; however, one of the resulting groups combined SARS-CoV-1, SARS-CoV-2 (WT) and MPV, which is potentially epidemiologically conflicting. The co-assignment heatmap (Figure 2B) shows instability within this cluster: while SARS-CoV-1 and SARS-CoV-2 (WT) co-assign in 89% of Monte Carlo iterations, MPV co-assigns with both, slightly more than half of the time.

The final, stable classification of 18 pathogens based on their epidemiological and transmission characteristics. The horizontal branch lengths represent the dissimilarity between clusters, with shorter branches indicating a higher frequency of co-assignment in the underlying Monte Carlo simulations.

When the dendrogram was instead cut at K=6 (Figure 3), it resolved into more epidemiologically coherent clusters, with all major pathogen groups forming stable, internally consistent archetypes across the ensemble of Monte Carlo simulations. Given this, the six-cluster solution was selected as the final consensus classification. The characteristics of each archetype are detailed in **Table 2**.

**Figure 3.**
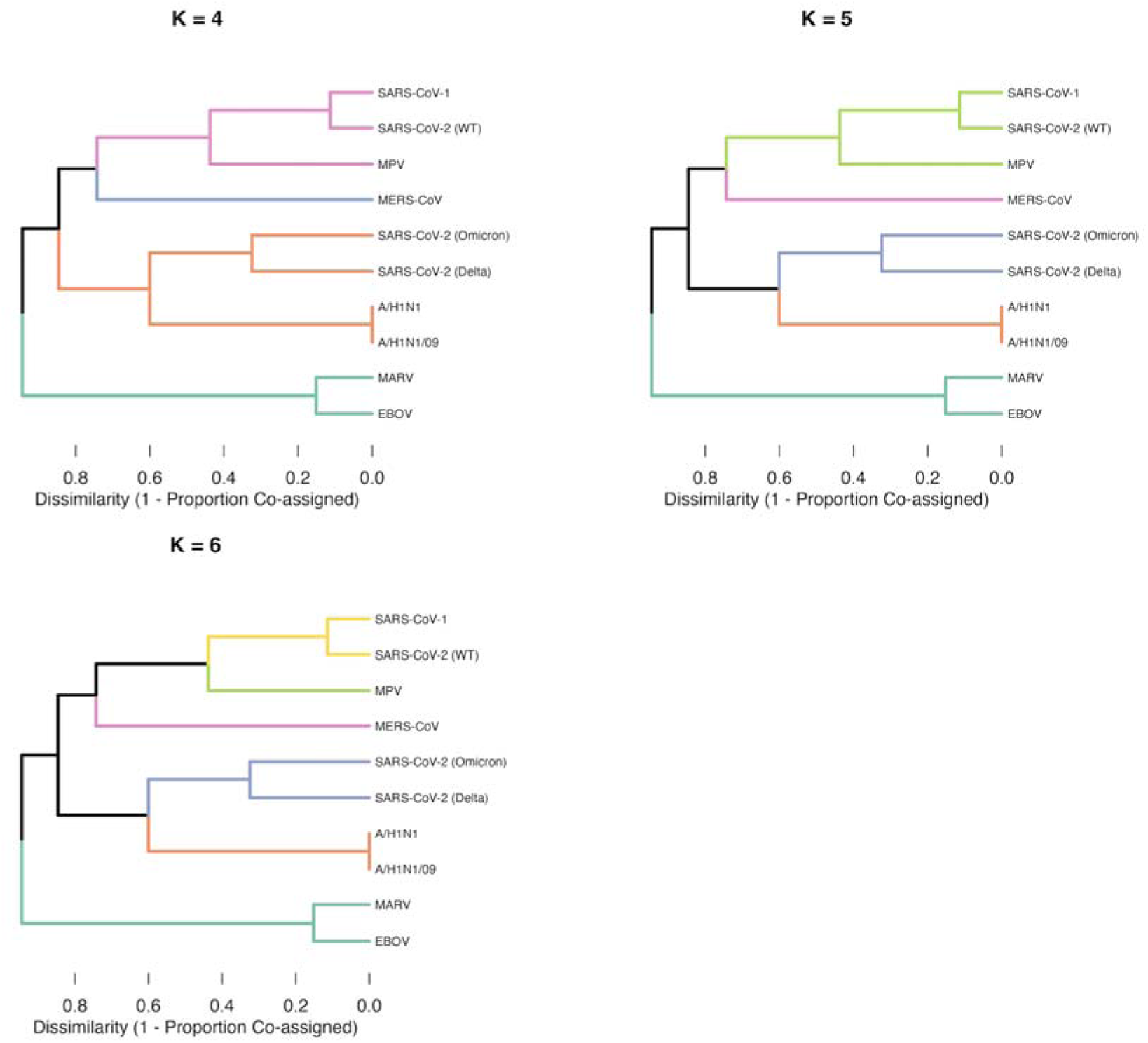
Hierarchical clustering with R_0_, serial interval, *k*, incubation period, latent and infectious period, case fatality risk and transmission route. K=4, 5, 6

**Table 2.** Archetype characterisation of Figure 3 when K=6.

| Table 2. Archetype characterisation of Figure 3 when K=6 |  |  |  |  |  |  |  |  |  |
| --- | --- | --- | --- | --- | --- | --- | --- | --- | --- |
| Cluster | Pathogens | Archetype parameters (mean (95% CI)) |  |  |  |  |  |  | Transmission route |
|  |  | R | k | Serial interval (d) | Incubation period (d) | Latent period (d) | Infectious period (d) | CFR (%) |  |
| 1 | COVID-19 (Delta and Omicron) | 7.38 (3.50–11.40) | 0.41 (0.10–0.82) | 3.55 (2.99–4.20) | 4.09 (3.33–5.03) | 3.57 (2.51–4.60) | 4.94 (3.06–7.74) | 0.1 (0.0–0.2) | Respiratory (n=2) |
| 2 | COVID-19 (WT), SARS-CoV-1 | 1.36 (1.22–3.37) | 0.36 (0.14–0.74) | 6.36 (2.17–16.38) | 5.08 (3.85–6.64) | 5.77 (5.01–6.71) | 8.33 (4.63–12.07) | 0.6 (0.2–10) | Respiratory (n=2) |
| 3 | EBOV, MARV | 1.88 (0.38–4.11) | 0.57 (0.18–1.06) | 11.92 (3.47–17.18) | 7.52 (4.37–9.34) | 8.71 (6.06–12.27) | 5.03 (3.07–8.38) | 60 (43–78) | Animal to human (n=2)<br>Direct contact (n=2) |
| 4 | A/H1N1, A/H1N1pdm09 | 1.88 (1.44–2.85) | 4.43 (0.65–10.15) | 2.73 (2.12–3.38) | 1.50 (1.34–1.85) | 2.14 (1.51–3.04) | 2.13 (0.56–4.42) | 0.1 (0.0–0.3) | Respiratory (n=2) |
| 5 | MERS-CoV | 1.40 (0.42–3.08) | 4.41 (0.0–49.01) | 12.41 (7.67–16.50) | 6.71 (5.73–8.03) | 7.00 (7.00–7.00) | 16.01 (9.45–24.27) | 39 (26–53) | Animal to human (n=1)<br>Respiratory (n=1) |
| 6 | MPV | 1.30 (0.80–1.85) | 0.48 (0.23–1.01) | 9.69 (7.38–13.97) | 8.58 (6.62–11.13) | 3.00 (3.00–3.00) | 6.36 (3.19–10.00) | 5.4 (2.4–8.3) | Animal to human (n=1)<br>Direct contact (n=1) |

The dendrogram from figure 2 is cut at K = 4, 5, and 6 clusters to show how pathogen groupings change as the tree is partitioned at different resolutions. Stable groupings appear where pathogens remain together across multiple values of K, whereas splits or reassignments indicate less well-defined relationships within the consensus structure.

### 3.4 Archetype characterisation

#### 3.4.1 Highly transmissible Coronaviruses

Archetype 1 contains the SARS-CoV-2 Delta and Omicron variants, which form a distinct high-transmission respiratory group. These pathogens have the highest reproductive number among all clusters (mean R of 7.38 (95% CI: 3.50–11.40) short serial intervals (3.55 (95% CI:2.99–4.20), and relatively short latent and infectious periods. The CFR is low (0.1 (95% CI: 0.0–0.2)).

#### 3.4.2 Moderately transmissible Coronaviruses

Archetype 2 comprises SARS-CoV-1 and the wild-type strain of SARS-CoV-2. These pathogens exhibit lower transmissivity (1.36 (95% CI: 1.22–3.37)), longer serial intervals (6.36 (95% CI: 2.17–16.38)), and longer infectious periods compared with Archetype 1. CFR displays a wider interval from 0.2 to 10%

#### 3.4.3 High-severity contact and zoonotic pathogens

Archetype 3 groups EBOV and MARV. These pathogens have very high CFRs (60 (95% CI: 43–78)), with the interval for human-to-human transmissibility ranging from 0.38 to 4.11. They display long serial, incubation and latent periods.

#### 3.4.4 Influenza viruses

Archetype 4 contains A/H1N1 and A/H1N1pdm09. These pathogens show moderately high transmissibility (1.88 (95% CI:1.44–2.85)), short incubation periods (1.50 (95% CI:1.34–1.85)) and short infectious periods (2.13 (95% CI: 0.56–4.42)). CFRs are low (0.1% (95% CI: 0.0–0.3)).

#### 3.4.5 MERS-CoV-like

Archetype 5 consists solely of MERS-CoV. The pathogen is characterised by a long serial interval (12.41 (95% CI:7.67–16.50)), long infectious period (16.01 (95% CI: 9.45–24.27)), and very high CFR (39% (CI:26–53)). Transmissibility is varied with the confidence interval ranging from 0.42 to 3.0.

#### 3.4.6 MPV-like

Archetype 6 consists of MPV. MPV exhibits moderate incubation (8.58 (95% CI:6.62–11.13)) and infectious periods (6.36 (95% CI: 3.19–10.00)). Moderate contact driven transmissibility (1.30 (95% CI: 0.80–1.85)), and a CFR of 5.4% (95% CI: 2.4–8.3).

#### 3.4.7 Sensitivity Analysis

To assess the robustness of the consensus clustering, we repeated the analysis using a reduced parameter set consisting of R, SI, and CFR, with (**Supplementary Figures S5 and S6**) and without presymptomatic transmission (**Supplementary Figures S7 and S8**). In both scenarios, pathogens grouped into larger and less clearly defined clusters, reflecting the limited discriminatory power of these parameters when used in isolation. When presymptomatic transmission was introduced SARS-CoV-2 viruses formed their own cluster separate to the Influenza A viruses (**Supplementary Figures S5 & S7).** Additionally, LASV also changed clusters when this parameter was added.

We assessed the flexibility of the framework by applying it to a more diverse range of pathogens using a core set of widely applicable parameters: R, IP, CFR, and transmission route. The framework incorporated and classified a wide variety of additional pathogens (**Supplementary Figure S11**).

## 4. Discussion

We reviewed key epidemiological parameters for 19 pathogens with pandemic potential and applied clustering algorithms to identify pathogen archetypes that share similar characteristics. Our findings suggest that grouping pathogens based on transmission traits could provide a pragmatic approach to pandemic preparedness.

The most frequently reported parameters were the incubation period, reproduction number, and CFR. However, data availability was uneven, with SARS-CoV-2 and influenza accounting for nearly half of all estimates. Parameter estimates varied both across pathogens and within studies of the same pathogen, aligning with previous reviews [13]. Notably, R, serial interval, and CFR estimates were highly context dependent. For example, MERS-CoV R estimates ranged from 5.4 (95% CI: 4.61-6.19) in an uncontrolled hospital outbreak to 0.14 (95% CI: 0.04-0.26) with control measures in place [14]. Similarly, influenza A/H1N1 RL estimates were higher in confined settings compared to overall estimates [15]. Likewise, EBOV estimates varied by country during the 2013-2016 epidemic [16]. Serial interval estimates also decreased when control measures were implemented, with the serial interval of SARS-CoV-2 decreasing post epidemic peak in China correlating with decreased time to isolation [17].

There were considerable differences in CFR estimates between outbreaks of the same pathogen. Influenza A/H5N1 varies by clade [18]. MPV varies when hospital care is available [19] or when comparing outcomes from outbreaks in Africa to outbreaks in the United States [20]. Varying estimates for NiV highlights how CFR can vary depending on country, strain and the control measures implemented, with the CFR being lower in Singapore (1999) compared to Malaysia (1998-1999) [21]. These examples illustrate that parameter estimates are generated across a wide range of contexts, and the importance of contextualising parameter estimates when applying them to modelling efforts.

Our clustering analysis identified six pathogen archetypes, each with shared characteristics that could inform the development of group-based, rather than pathogen-specific, control strategies (Figure 3). Given that key interventions like contact tracing and case isolation are directly influenced by parameters such as R, the serial interval, and the proportion of presymptomatic transmission [22,23]. Pathogen-specific planning however, is and will remain useful. Clustering pathogens with shared epidemiology would allow for specific plans to be adapted for pathogens that share similar characteristics. A dual approach would ideally allow for both in depth preparation for known risks and breadth in preparedness for a wider range of pathogens.

The archetypes proposed are a result of the quality, context, and variability of the underlying input parameters. Without classifying pathogens by setting and context, the archetypes must try and capture the range of plausible epidemiological behaviours that a pathogen can exhibit across different settings. However, this may bias the central tendency for parameters like R upwards, potentially misrepresenting a pathogen’s behaviour in more common scenarios. Therefore, the use of this framework is to outline a plausible parameter range for each archetype. Archetype 3 (EBOV and MARV) for example spans 0.38–4.11 with a mean of 1.88. When planning for an EBOV/MARV-like pathogen there may be settings where transmission is limited [13] or uncontrolled [16]. Likewise for a MERS-CoV-like pathogen in community settings transmission potential may be small, however in hospital settings there is a greater risk of secondary transmission [14]. Therefore, capturing the heterogeneity of these estimates is important to effectively plan for these pathogens.

Presymptomatic transmission was excluded from the main analysis as the estimates proved too uncertain to be meaningfully interpreted (**Supplementary Table S2)**. The values for SARS-CoV-2 variants and MPV were broadly consistent with published ranges [24–26]. Others showed biologically implausible estimates, such as MERS-CoV. This estimation is highly influenced by uncertainty in serial interval and incubation period estimates [27]. Because of the sensitivity to the underlying distributions, even small uncertainties or inconsistencies in the source data can produce large fluctuations in the presymptomatic estimates. Although including presymptomatic transmission in the clustering process (**Supplementary Figures S5 & S9**) produced some shifts in cluster membership, the consensus structure remained broadly robust. Given these issues, the presymptomatic transmission parameter was omitted from the primary clustering analysis to avoid overinterpreting estimates stemming from data uncertainty.

The estimates generated for the serial interval and R_0_ for LASV and CCHFV respectively are subject to considerable uncertainty. Human-to-human transmission for these pathogens is best studied in hospital settings, where nosocomial outbreaks have been documented. Identifying transmission for these pathogens in community or household settings is particularly difficult as it is hard to distinguish between vector/animal-to-human from human-to-human transmission. As a result, estimation of transmission parameters will largely rely on hospital-based outbreaks, as used in this study. Whilst these estimates are subject to significant uncertainty, they provide a feasible basis for inference. As such utilising these parameters within our framework (**Supplementary figure S5 & S7**) introduces additional uncertainty. Which is a limitation for this method.

These findings highlight the potential for this framework to serve as an adaptable tool for classifying and assessing pathogens beyond those included in this study. We extended the number of pathogens in the sensitivity analysis (**Supplementary Figure S11**) to include pathogens with different characteristics to the original selection. Highlighting that the framework could be used to classify water-borne pathogens, bioterrorism-related pathogens and pathogens with extended delay periods such as HIV. Additional work should address the impact of key interventions across each archetype. Moreover, the framework should be continuously updated as new epidemiological data emerge, refining the clustering methods and integrating additional pathogens, including both novel and emerging threats. To that end, the development of a global, standardised repository for epidemiological parameters would enable the rapid integration of data on novel and emerging threats.

Further research should be directed to explore the use of machine learning techniques to group pathogens. Our analysis reflects a set of decisions and assumptions that could reasonably be handled differently. Exploring how cluster membership changes under different approaches would help identify which aspects of the clustering are robust and which are most dependent on methodological choices. Such comparative work would support the development of more generalisable frameworks for future pandemic preparedness.

## 5. Conclusion

Documenting epidemiological parameters is crucial for effective outbreak risk analysis. We provide 302 parameter estimates for 19 pathogens, offering a valuable foundation for modelling their spread and containment. However, key transmission parameters such as the dispersion parameter and latent period remain underreported, highlighting the need for further research to strengthen outbreak preparedness. Our clustering approach demonstrates a practical framework for evaluating plausible parameter ranges across groups of similar pathogens. By maintaining a dynamic classification system, public health preparedness efforts can shift away from a reactive, pathogen-specific focus toward a more anticipatory, trait-based strategy for managing future infectious disease risks.

### Declaration of generative AI and AI-assisted technologies in the writing process

During the preparation of this work the author(s) used Gemini 2.5 pro, within the Cursor editor in order to generate code used in this analysis. After using this tool/service, the author(s) reviewed and edited the content as needed and take(s) full responsibility for the content of the publication.

## Supporting information

Supplementary infomation

## Data Availability

Code to reproduce this report is open on GitHub at https://github.com/oswaldogressani/Blueprint for incubation period estimates and https://github.com/Jward2847/archetypes# for serial interval, reproduction number and the clustering analysis.

## Acknowledgements

This work was supported by the ESCAPE project (101095619), co-funded by the European Union. Views and opinions expressed are however those of the author(s) only and do not necessarily reflect those of the European Union or European Health and Digital Executive Agency (HADEA). Neither the European Union nor the granting authority can be held responsible for them. This work was co-funded by UK Research and Innovation (UKRI) under the UK government’s Horizon Europe funding guarantee [grant number 10051037].

## Notes

### Competing Interest Statement

The authors have declared no competing interest.

### Summary of Updates

The replacement single-study use within the machine learning algorithm with a bootstrap aggregation method. The final clusters now incorporates evidence from all available studies listed in Table 1 for every parameter. In addition we have expanded the parameters included in the main analysis. This has changed the cluster membership.

## References

[1] Rietveld J, Hobson T, Mani L, Avin S, Sundaram L. The UK’s pandemic preparedness and early response to the COVID-19 pandemic. Glob Public Health 2024;19:2415499.

[2] Rt Hon the Baroness Hallett DBE. UK Covid-19 Inquiry. House of Commons; 2024.

[3] Looi M-K. What could the next pandemic be? BMJ 2023;381:909.

[4] Adalja AA, Watson M, Toner ES, Cicero A, Inglesby TV. Characteristics of microbes most likely to cause pandemics and global catastrophes. Curr Top Microbiol Immunol 2019;424:1–20.

5. [5] Prioritizing diseases for research and development in emergency contexts n.d. https://www.who.int/activities/prioritizing-diseases-for-research-and-development-in-emergency-contexts (accessed June 24, 2024).

[6] Gressani O. EpiLPS: A Fast and Flexible Bayesian Tool for Estimating Epidemiological Parameters. 2021.

[7] Gressani O, Torneri A, Hens N, Faes C. Flexible Bayesian estimation of incubation times. Am J Epidemiol 2025;194:490–501.

[8] Gressani O, Wallinga J, Althaus CL, Hens N, Faes C. EpiLPS: A fast and flexible Bayesian tool for estimation of the time-varying reproduction number. PLoS Comput Biol 2022;18:e1010618.

[9] Charniga K, Park SW, Akhmetzhanov AR, Cori A, Dushoff J, Funk S, et al. Best practices for estimating and reporting epidemiological delay distributions of infectious diseases. PLoS Comput Biol 2024;20:e1012520.

[10] Abbott S, Brand S, Pearson C, Funk S, Charniga K. primarycensored: Primary Event Censored Distributions 2025. 10.5281/zenodo.13632839.

[11] Cases of Crimean–Congo haemorrhagic fever infected in the EU/EEA, 2013–present. European Centre for Disease Prevention and Control 2021. https://www.ecdc.europa.eu/en/crimean-congo-haemorrhagic-fever/surveillance/cases-eu-since-2013 (accessed February 4, 2025).

[12] Hamerly G, Elkan C. Learning the k in k-means. Advances in Neural Information Processing Systems 2003;16.

[13] Nash RK, Bhatia S, Morgenstern C, Doohan P, Jorgensen D, McCain K, et al. Ebola virus disease mathematical models and epidemiological parameters: a systematic review. Lancet Infect Dis 2024;24:e762–73.

[14] Park J-E, Jung S, Kim A, Park J-E. MERS transmission and risk factors: a systematic review. BMC Public Health 2018;18:574.

[15] Biggerstaff M, Cauchemez S, Reed C, Gambhir M, Finelli L. Estimates of the reproduction number for seasonal, pandemic, and zoonotic influenza: a systematic review of the literature. BMC Infect Dis 2014;14:480.

[16] Muzembo BA, Kitahara K, Mitra D, Ntontolo NP, Ngatu NR, Ohno A, et al. The basic reproduction number (R0) of ebola virus disease: A systematic review and meta-analysis. Travel Med Infect Dis 2024;57:102685.

[17] Xu X, Wu Y, Kummer AG, Zhao Y, Hu Z, Wang Y, et al. Assessing changes in incubation period, serial interval, and generation time of SARS-CoV-2 variants of concern: a systematic review and meta-analysis. BMC Med 2023;21:374.

[18] Lai S, Qin Y, Cowling BJ, Ren X, Wardrop NA, Gilbert M, et al. Global epidemiology of avian influenza A H5N1 virus infection in humans, 1997-2015: a systematic review of individual case data. Lancet Infect Dis 2016;16:e108–18.

[19] DeWitt ME, Polk C, Williamson J, Shetty AK, Passaretti CL, McNeil CJ, et al. Global monkeypox case hospitalisation rates: A rapid systematic review and meta-analysis. EClinicalMedicine 2022;54:101710.

[20] Bunge EM, Hoet B, Chen L, Lienert F, Weidenthaler H, Baer LR, et al. The changing epidemiology of human monkeypox-A potential threat? A systematic review. PLoS Negl Trop Dis 2022;16:e0010141.

[21] Hegde ST, Lee KH, Styczynski A, Jones FK, Gomes I, Das P, et al. Potential for person-to-person transmission of henipaviruses: A systematic review of the literature. J Infect Dis 2024;229:733–42.

[22] Nishiura H, Linton NM, Akhmetzhanov AR. Serial interval of novel coronavirus (COVID-19) infections. Int J Infect Dis 2020;93:284–6.

[23] Fraser C, Riley S, Anderson RM, Ferguson NM. Factors that make an infectious disease outbreak controllable. Proc Natl Acad Sci U S A 2004;101:6146–51.

[24] Tindale LC, Stockdale JE, Coombe M, Garlock ES, Lau WYV, Saraswat M, et al. Evidence for transmission of COVID-19 prior to symptom onset. Elife 2020;9. 10.7554/eLife.57149.

[25] Casey-Bryars M, Griffin J, McAloon C, Byrne A, Madden J, Mc Evoy D, et al. Presymptomatic transmission of SARS-CoV-2 infection: a secondary analysis using published data. BMJ Open 2021;11:e041240.

[26] Ward T, Christie R, Paton RS, Cumming F, Overton CE. Transmission dynamics of monkeypox in the United Kingdom: contact tracing study. BMJ 2022;379:e073153.

[27] Slifka MK, Gao L. Is presymptomatic spread a major contributor to COVID-19 transmission? Nat Med 2020;26:1531–3.

[28] Fraser C, Cummings DAT, Klinkenberg D, Burke DS, Ferguson NM. Influenza transmission in households during the 1918 pandemic. Am J Epidemiol 2011;174:505–14.

[29] Vink MA, Bootsma MCJ, Wallinga J. Serial intervals of respiratory infectious diseases: a systematic review and analysis. Am J Epidemiol 2014;180:865–75.

[30] Nishiura H. Early efforts in modeling the incubation period of infectious diseases with an acute course of illness. Emerg Themes Epidemiol 2007;4:2.

[31] Cori A, Valleron AJ, Carrat F, Scalia Tomba G, Thomas G, Boëlle PY. Estimating influenza latency and infectious period durations using viral excretion data. Epidemics 2012;4:132–8.

[32] Carrat F, Luong J, Lao H, Sallé A-V, Lajaunie C, Wackernagel H. A “small-world-like” model for comparing interventions aimed at preventing and controlling influenza pandemics. BMC Med 2006;4:26.

[33] WHO. Pandemic Influenza Risk Management. 2017.

[34] Brugger J, Althaus CL. Transmission of and susceptibility to seasonal influenza in Switzerland from 2003 to 2015. Epidemics 2020;30:100373.

[35] Tom BDM, Van Hoek AJ, Pebody R, McMenamin J, Robertson C, Catchpole M, et al. Estimating time to onset of swine influenza symptoms after initial novel A(H1N1v) viral infection. Epidemiol Infect 2011;139:1418–24.

[36] Lessler J, Reich NG, Brookmeyer R, Perl TM, Nelson KE, Cummings DAT. Incubation periods of acute respiratory viral infections: a systematic review. Lancet Infect Dis 2009;9:291–300.

[37] Lessler J, Reich NG, Cummings DAT, New York City Department of Health and Mental Hygiene Swine Influenza Investigation Team, Nair HP, Jordan HT, et al. Outbreak of 2009 pandemic influenza A (H1N1) at a New York City school. N Engl J Med 2009;361:2628–36.

[38] Tuite AR, Greer AL, Whelan M, Winter A-L, Lee B, Yan P, et al. Estimated epidemiologic parameters and morbidity associated with pandemic H1N1 influenza. CMAJ 2010;182:131–6.

[39] Van Kerkhove MD, Hirve S, Koukounari A, Mounts AW, H1N1pdm serology working group. Estimating age-specific cumulative incidence for the 2009 influenza pandemic: a meta-analysis of A(H1N1)pdm09 serological studies from 19 countries. Influenza Other Respi Viruses 2013;7:872–86.

[40] Wong JY, Kelly H, Ip DKM, Wu JT, Leung GM, Cowling BJ. Case fatality risk of influenza A (H1N1pdm09): a systematic review: A systematic review. Epidemiology 2013;24:830–41.

[41] Carrat F, Vergu E, Ferguson NM, Lemaitre M, Cauchemez S, Leach S, et al. Time lines of infection and disease in human influenza: a review of volunteer challenge studies. Am J Epidemiol 2008;167:775–85.

[42] Elveback LR, Fox JP, Ackerman E, Langworthy A, Boyd M, Gatewood L. An influenza simulation model for immunization studies. Am J Epidemiol 1976;103:152–65.

[43] Ferguson NM, Cummings DAT, Cauchemez S, Fraser C, Riley S, Meeyai A, et al. Strategies for containing an emerging influenza pandemic in Southeast Asia. Nature 2005;437:209–14.

[44] Cauchemez S, Carrat F, Viboud C, Valleron AJ, Boëlle PY. A Bayesian MCMC approach to study transmission of influenza: application to household longitudinal data. Stat Med 2004;23:3469–87.

[45] Bettencourt LMA, Ribeiro RM. Real time bayesian estimation of the epidemic potential of emerging infectious diseases. PLoS One 2008;3:e2185.

[46] Aditama TY, Samaan G, Kusriastuti R, Sampurno OD, Purba W, Misriyah, et al. Avian influenza H5N1 transmission in households, Indonesia. PLoS One 2012;7:e29971.

[47] Yang Y, Halloran ME, Sugimoto JD, Longini IM Jr. Detecting human-to-human transmission of avian influenza A (H5N1). Emerg Infect Dis 2007;13:1348–53.

[48] Ferguson NM, Fraser C, Donnelly CA, Ghani AC, Anderson RM. Public health. Public health risk from the avian H5N1 influenza epidemic. Science 2004;304:968–9.

[49] Ward J, Lambert JW, Russell TW, Azam JM, Kucharski AJ, Funk S, et al. Estimates of epidemiological parameters for H5N1 influenza in humans: a rapid review. medRxiv 2024:2024.12.11.24318702. 10.1101/2024.12.11.24318702.

[50] Beigel JH, Farrar J, Han AM, Hayden FG, Hyer R, de Jong MD, et al. Avian Influenza A (H5N1) Infection in Humans. N Engl J Med 2005;353:1374–85.

[51] Huai Y, Xiang N, Zhou L, Feng L, Peng Z, Chapman RS, et al. Incubation period for human cases of avian influenza A (H5N1) infection, China. Emerg Infect Dis 2008;14:1819–21.

[52] Cowling BJ, Jin L, Lau EHY, Liao Q, Wu P, Jiang H, et al. Comparative epidemiology of human infections with avian influenza A H7N9 and H5N1 viruses in China: a population-based study of laboratory-confirmed cases. Lancet 2013;382:129–37.

[53] Oner AF, Bay A, Arslan S, Akdeniz H, Sahin HA, Cesur Y, et al. Avian influenza A (H5N1) infection in eastern Turkey in 2006. N Engl J Med 2006;355:2179–85.

[54] Van Kerkhove MD, Mumford E, Mounts AW, Bresee J, Ly S, Bridges CB, et al. Highly pathogenic avian influenza (H5N1): pathways of exposure at the animal-human interface, a systematic review. PLoS One 2011;6:e14582.

[55] Li FCK, Choi BCK, Sly T, Pak AWP. Evidence-based public health policy and practice: Finding the real case-fatality rate of H5N1 avian influenza. Journal of Epidemiology and Community Health (1979-) 2008;62:555–9.

[56] Bente DA, Forrester NL, Watts DM, McAuley AJ, Whitehouse CA, Bray M. Crimean-Congo hemorrhagic fever: history, epidemiology, pathogenesis, clinical syndrome and genetic diversity. Antiviral Res 2013;100:159–89.

[57] Belhadi D, El Baied M, Mulier G, Malvy D, Mentré F, Laouénan C. The number of cases, mortality and treatments of viral hemorrhagic fevers: A systematic review. PLoS Negl Trop Dis 2022;16:e0010889.

[58] Belobo JTE, Kenmoe S, Kengne-Nde C, Emoh CPD, Bowo-Ngandji A, Tchatchouang S, et al. Worldwide epidemiology of Crimean-Congo Hemorrhagic Fever Virus in humans, ticks and other animal species, a systematic review and meta-analysis. PLoS Negl Trop Dis 2021;15:e0009299.

[59] Perveen N, Khan G. Crimean-Congo hemorrhagic fever in the Arab world: A systematic review. Front Vet Sci 2022;9:938601.

[60] Nasirian H. New aspects about Crimean-Congo hemorrhagic fever (CCHF) cases and associated fatality trends: A global systematic review and meta-analysis. Comp Immunol Microbiol Infect Dis 2020;69:101429.

[61] Wong ZSY, Bui CM, Chughtai AA, Macintyre CR. A systematic review of early modelling studies of Ebola virus disease in West Africa. Epidemiol Infect 2017;145:1069–94.

[62] Velásquez GE, Aibana O, Ling EJ, Diakite I, Mooring EQ, Murray MB. Time From Infection to Disease and Infectiousness for Ebola Virus Disease, a Systematic Review. Clin Infect Dis 2015;61:1135–40.

[63] Khan SA, Imtiaz MA, Islam MM, Tanzin AZ, Islam A, Hassan MM. Major bat-borne zoonotic viral epidemics in Asia and Africa: A systematic review and meta-analysis. Vet Med Sci 2022;8:1787–801.

[64] Lo Iacono G, Cunningham AA, Fichet-Calvet E, Garry RF, Grant DS, Khan SH, et al. Using modelling to disentangle the relative contributions of zoonotic and anthroponotic transmission: the case of lassa fever. PLoS Negl Trop Dis 2015;9:e3398.

[65] Zhao S, Musa SS, Fu H, He D, Qin J. Large-scale Lassa fever outbreaks in Nigeria: quantifying the association between disease reproduction number and local rainfall. Epidemiol Infect 2020;148:e4.

[66] Doohan P, Jorgensen D, Naidoo TM, McCain K, Hicks JT, McCabe R, et al. Lassa fever outbreaks, mathematical models, and disease parameters: a systematic review and meta-analysis. Lancet Glob Health 2024;12:e1962–72.

[67] Akhmetzhanov AR, Asai Y, Nishiura H. Quantifying the seasonal drivers of transmission for Lassa fever in Nigeria. Philos Trans R Soc Lond B Biol Sci 2019;374:20180268.

[68] Tuite AR, Watts AG, Kraemer MUG, Khan K, Bogoch II. Potential for Seasonal Lassa Fever Case Exportation from Nigeria. Am J Trop Med Hyg 2019;100:647–51.

[69] Wolf T, Ellwanger R, Goetsch U, Wetzstein N, Gottschalk R. Fifty years of imported Lassa fever: a systematic review of primary and secondary cases. J Travel Med 2020;27. 10.1093/jtm/taaa035.

[70] Kenmoe S, Tchatchouang S, Ebogo-Belobo JT, Ka’e AC, Mahamat G, Guiamdjo Simo RE, et al. Systematic review and meta-analysis of the epidemiology of Lassa virus in humans, rodents and other mammals in sub-Saharan Africa. PLoS Negl Trop Dis 2020;14:e0008589.

[71] Dwalu E, Jetoh RW, Shobayo BI, Pewu I, Taweh F, Wilson-Sesay HW, et al. Trend of Lassa fever cases and factors associated with mortality in Liberia, 2016 - 2021: a secondary data analysis. Pan Afr Med J 2024;47:22.

[72] Qian GY, Edmunds WJ, Bausch DG, Jombart T. A mathematical model of Marburg virus disease outbreaks and the potential role of vaccination in control. BMC Med 2023;21:439.

[73] Ajelli M, Merler S. Transmission potential and design of adequate control measures for Marburg hemorrhagic fever. PLoS One 2012;7:e50948.

[74] Pavlin BI. Calculation of incubation period and serial interval from multiple outbreaks of Marburg virus disease. BMC Res Notes 2014;7:906.

[75] Bettencourt LMA. An Ensemble Trajectory Method for Real-Time Modeling and Prediction of Unfolding Epidemics: Analysis of the 2005 Marburg Fever Outbreak in Angola. Mathematical and Statistical Estimation Approaches in Epidemiology 2009:143.

[76] Cuomo-Dannenburg G, McCain K, McCabe R, Unwin HJT, Doohan P, Nash RK, et al. Marburg virus disease outbreaks, mathematical models, and disease parameters: a systematic review. Lancet Infect Dis 2024;24:e307–17.

[77] Peak CM, Childs LM, Grad YH, Buckee CO. Comparing nonpharmaceutical interventions for containing emerging epidemics. Proc Natl Acad Sci U S A 2017;114:4023–8.

[78] Wang J, Chen X, Guo Z, Zhao S, Huang Z, Zhuang Z, et al. Superspreading and heterogeneity in transmission of SARS, MERS, and COVID-19: A systematic review. Comput Struct Biotechnol J 2021;19:5039–46.

[79] Assiri A, McGeer A, Perl TM, Price CS, Al Rabeeah AA, Cummings DAT, et al. Hospital outbreak of Middle East respiratory syndrome coronavirus. N Engl J Med 2013;369:407–16.

[80] Park SH, Kim Y-S, Jung Y, Choi SY, Cho N-H, Jeong HW, et al. Outbreaks of Middle East Respiratory Syndrome in Two Hospitals Initiated by a Single Patient in Daejeon, South Korea. Infect Chemother 2016;48:99–107.

[81] Cowling BJ, Park M, Fang VJ, Wu P, Leung GM, Wu JT. Preliminary epidemiological assessment of MERS-CoV outbreak in South Korea, May to June 2015. Euro Surveill 2015;20:7–13.

[82] Cauchemez S, Fraser C, Van Kerkhove MD, Donnelly CA, Riley S, Rambaut A, et al. Middle East respiratory syndrome coronavirus: quantification of the extent of the epidemic, surveillance biases, and transmissibility. Lancet Infect Dis 2014;14:50–6.

[83] Lessler J, Rodriguez-Barraquer I, Cummings DAT, Garske T, Van Kerkhove M, Mills H, et al. Estimating Potential Incidence of MERS-CoV Associated with Hajj Pilgrims to Saudi Arabia, 2014. PLoS Curr 2014;6. 10.1371/currents.outbreaks.c5c9c9abd636164a9b6fd4dbda974369.

[84] Okoli GN, Van Caeseele P, Askin N, Abou-Setta AM. A global systematic evidence review with meta-analysis of the epidemiological characteristics of the 2022 Mpox outbreaks. Infection 2024;52:901–21.

[85] Beer EM, Rao VB. A systematic review of the epidemiology of human monkeypox outbreaks and implications for outbreak strategy. PLoS Negl Trop Dis 2019;13:e0007791.

[86] Blumberg S, Lloyd-Smith JO. Inference of R(0) and transmission heterogeneity from the size distribution of stuttering chains. PLoS Comput Biol 2013;9:e1002993.

[87] Paredes MI, Ahmed N, Figgins M, Colizza V, Lemey P, McCrone JT, et al. Underdetected dispersal and extensive local transmission drove the 2022 mpox epidemic. Cell 2024;187:1374–86.e13.

88. [88] Second meeting of the International Health Regulations (2005) (IHR) Emergency Committee regarding the multi-country outbreak of monkeypox n.d. https://www.who.int/news/item/23-07-2022-second-meeting-of-the-international-health-regulations-(2005)-(ihr)-emergency-committee-regarding-the-multi-country-outbreak-of-monkeypox (accessed November 10, 2025).

[89] Chenchula S, Ghanta MK, Amerneni KC, Rajakarunakaran P, Chandra MB, Chavan M, et al. A systematic review to identify novel clinical characteristics of monkeypox virus infection and therapeutic and preventive strategies to combat the virus. Arch Virol 2023;168:195.

[90] Okoli GN, Van Caeseele P, Askin N, Abou-Setta AM. Comparative evaluation of the clinical presentation and epidemiology of the 2022 and previous Mpox outbreaks: a rapid review and meta-analysis. Infect Dis 2023;55:490–508.

[91] Hatami H, Jamshidi P, Arbabi M, Safavi-Naini SAA, Farokh P, Izadi-Jorshari G, et al. Demographic, Epidemiologic, and Clinical Characteristics of Human Monkeypox Disease Pre- and Post-2022 Outbreaks: A Systematic Review and Meta-Analysis. Biomedicines 2023;11. 10.3390/biomedicines11030957.

[92] Madewell ZJ, Charniga K, Masters NB, Asher J, Fahrenwald L, Still W, et al. Serial Interval and Incubation Period Estimates of Monkeypox Virus Infection in 12 Jurisdictions, United States, May-August 2022. Emerg Infect Dis 2023;29:818–21.

[93] Miura F, van Ewijk CE, Backer JA, Xiridou M, Franz E, Op de Coul E, et al. Estimated incubation period for monkeypox cases confirmed in the Netherlands, May 2022. Euro Surveill 2022;27:2200448.

[94] Asakura TR, Jung S-M, Murayama H, Ghaznavi C, Sakamoto H, Teshima A, et al. Projecting international mpox spread in Asia: ongoing global health risk. medRxiv 2024. 10.1101/2024.04.17.24305832.

[95] Endo A, Murayama H, Abbott S, Ratnayake R, Pearson CAB, Edmunds WJ, et al. Heavy-tailed sexual contact networks and monkeypox epidemiology in the global outbreak, 2022. Science 2022;378:90–4.

[96] Xiridou M, Miura F, Adam P, Op de Coul E, de Wit J, Wallinga J. The Fading of the Mpox Outbreak Among Men Who Have Sex With Men: A Mathematical Modelling Study. The Journal of Infectious Diseases 2024;230:e121–30.

[97] Benites-Zapata VA, Ulloque-Badaracco JR, Alarcon-Braga EA, Hernandez-Bustamante EA, Mosquera-Rojas MD, Bonilla-Aldana DK, et al. Clinical features, hospitalisation and deaths associated with monkeypox: a systematic review and meta-analysis. Ann Clin Microbiol Antimicrob 2022;21:36.

[98] Nikolay B, Salje H, Hossain MJ, Khan AKMD, Sazzad HMS, Rahman M, et al. Transmission of Nipah Virus - 14 Years of Investigations in Bangladesh. N Engl J Med 2019;380:1804–14.

[99] Bradbury NV, Hart WS, Lovell-Read FA, Polonsky JA, Thompson RN. Exact calculation of end-of-outbreak probabilities using contact tracing data. J R Soc Interface 2023;20:20230374.

[100] Kenmoe S, Demanou M, Bigna JJ, Nde Kengne C, Fatawou Modiyinji A, Simo FBN, et al. Case fatality rate and risk factors for Nipah virus encephalitis: A systematic review and meta-analysis. J Clin Virol 2019;117:19–26.

[101] Suman N, Khandelwal E, Chiluvuri P, Rami DS, Chansoria S, Jerry A, et al. NIPAH virus encephalitis: Unveiling the epidemiology, risk factors, and clinical outcomes - A systematic review and meta-analysis. J Pharm Bioallied Sci 2024;16:S102–5.

[102] Rudolph KE, Lessler J, Moloney RM, Kmush B, Cummings DAT. Incubation periods of mosquito-borne viral infections: a systematic review. Am J Trop Med Hyg 2014;90:882–91.

[103] Ebogo-Belobo JT, Kenmoe S, Abanda NN, Bowo-Ngandji A, Mbaga DS, Magoudjou-Pekam JN, et al. Contemporary epidemiological data of Rift Valley fever virus in humans, mosquitoes and other animal species in Africa: A systematic review and meta-analysis. Vet Med Sci 2023;9:2309–28.

[104] Lipsitch M, Cohen T, Cooper B, Robins JM, Ma S, James L, et al. Transmission dynamics and control of severe acute respiratory syndrome. Science 2003;300:1966–70.

[105] Riley S, Fraser C, Donnelly CA, Ghani AC, Abu-Raddad LJ, Hedley AJ, et al. Transmission dynamics of the etiological agent of SARS in Hong Kong: impact of public health interventions. Science 2003;300:1961–6.

[106] Chen S-C, Chang C-F, Liao C-M. Predictive models of control strategies involved in containing indoor airborne infections. Indoor Air 2006;16:469–81.

[107] Chowell G, Castillo-Chavez C, Fenimore PW, Kribs-Zaleta CM, Arriola L, Hyman JM. Model parameters and outbreak control for SARS. Emerg Infect Dis 2004;10:1258–63.

[108] Tsang KW, Ho PL, Ooi GC, Yee WK, Wang T, Chan-Yeung M, et al. A cluster of cases of severe acute respiratory syndrome in Hong Kong. N Engl J Med 2003;348:1977–85.

[109] Becker NG, Glass K, Li Z, Aldis GK. Controlling emerging infectious diseases like SARS. Math Biosci 2005;193:205–21.

[110] Klinkenberg D, Fraser C, Heesterbeek H. The effectiveness of contact tracing in emerging epidemics. PLoS One 2006;1:e12.

[111] Lloyd-Smith JO, Galvani AP, Getz WM. Curtailing transmission of severe acute respiratory syndrome within a community and its hospital. Proc Biol Sci 2003;270:1979–89.

[112] Hui DSC, Chan MCH, Wu AK, Ng PC. Severe acute respiratory syndrome (SARS): epidemiology and clinical features. Postgrad Med J 2004;80:373–81.

[113] Ahammed T, Anjum A, Rahman MM, Haider N, Kock R, Uddin MJ. Estimation of novel coronavirus (COVID-19) reproduction number and case fatality rate: A systematic review and meta-analysis. Health Sci Rep 2021;4:e274.

[114] Lin Y-F, Duan Q, Zhou Y, Yuan T, Li P, Fitzpatrick T, et al. Spread and Impact of COVID-19 in China: A Systematic Review and Synthesis of Predictions From Transmission-Dynamic Models. Front Med 2020;7:321.

[115] Xie Y, Wang Z, Liao H, Marley G, Wu D, Tang W. Epidemiologic, clinical, and laboratory findings of the COVID-19 in the current pandemic: systematic review and meta-analysis. BMC Infect Dis 2020;20:640.

[116] Izadi N, Taherpour N, Mokhayeri Y, Sotoodeh Ghorbani S, Rahmani K, Hashemi Nazari SS. Epidemiologic Parameters for COVID-19: A Systematic Review and Meta-Analysis. Med J Islam Repub Iran 2022;36:155.

[117] Du Z, Wang C, Liu C, Bai Y, Pei S, Adam DC, et al. Systematic review and meta-analyses of superspreading of SARS-CoV-2 infections. Transbound Emerg Dis 2022;69:e3007–14.

[118] Backer JA, Klinkenberg D, Wallinga J. Incubation period of 2019 novel coronavirus (2019-nCoV) infections among travellers from Wuhan, China, 20-28 January 2020. Euro Surveill 2020;25. 10.2807/1560-7917.ES.2020.25.5.2000062.

[119] Xin H, Li Y, Wu P, Li Z, Lau EHY, Qin Y, et al. Estimating the Latent Period of Coronavirus Disease 2019 (COVID-19). Clin Infect Dis 2022;74:1678–81.

[120] He X, Lau EHY, Wu P, Deng X, Wang J, Hao X, et al. Temporal dynamics in viral shedding and transmissibility of COVID-19. Nat Med 2020;26:672–5.

[121] Zhu H, Li Y, Jin X, Huang J, Liu X, Qian Y, et al. Transmission dynamics and control methodology of COVID-19: A modeling study. Appl Math Model 2021;89:1983–98.

[122] Nishiura H, Kobayashi T, Yang Y, Hayashi K, Miyama T, Kinoshita R, et al. The rate of underascertainment of novel Coronavirus (2019-nCoV) infection: Estimation using Japanese passengers data on evacuation flights. J Clin Med 2020;9:419.

[123] Mizumoto K, Kagaya K, Chowell G. Early epidemiological assessment of the transmission potential and virulence of coronavirus disease 2019 (COVID-19) in Wuhan City, China, January-February, 2020. BMC Med 2020;18:217.

[124] Brazeau NF, Verity R, Jenks S, Fu H, Whittaker C, Winskill P, et al. Estimating the COVID-19 infection fatality ratio accounting for seroreversion using statistical modelling. Commun Med (Lond) 2022;2:54.

[125] Davies NG, Abbott S, Barnard RC, Jarvis CI, Kucharski AJ, Munday JD, et al. Estimated transmissibility and impact of SARS-CoV-2 lineage B.1.1.7 in England. Science 2021;372. 10.1126/science.abg3055.

[126] An der Heiden M, Buchholz U. Serial interval in households infected with SARS-CoV-2 variant B.1.1.529 (Omicron) is even shorter compared to Delta. Epidemiol Infect 2022;150:e165.

[127] Hart WS, Miller E, Andrews NJ, Waight P, Maini PK, Funk S, et al. Generation time of the alpha and delta SARS-CoV-2 variants: an epidemiological analysis. Lancet Infect Dis 2022;22:603–10.

[128] Zhang T, Nishiura H. Estimating infection fatality risk and ascertainment bias of COVID-19 in Osaka, Japan from February 2020 to January 2022. Sci Rep 2023;13:5540.

[129] Liu Y, Rocklöv J. The reproductive number of the Delta variant of SARS-CoV-2 is far higher compared to the ancestral SARS-CoV-2 virus. J Travel Med 2021;28. 10.1093/jtm/taab124.

[130] Ryu S, Kim D, Lim J-S, Ali ST, Cowling BJ. Serial Interval and Transmission Dynamics during SARS-CoV-2 Delta Variant Predominance, South Korea. Emerg Infect Dis 2022;28:407–10.

[131] Zhao S, Guo Z, Chong MKC, He D, Wang MH. Superspreading potential of SARS-CoV-2 Delta variants under intensive disease control measures in China. J Travel Med 2022;29. 10.1093/jtm/taac025.

[132] Backer JA, Eggink D, Andeweg SP, Veldhuijzen IK, van Maarseveen N, Vermaas K, et al. Shorter serial intervals in SARS-CoV-2 cases with Omicron BA.1 variant compared with Delta variant, the Netherlands, 13 to 26 December 2021. Euro Surveill 2022;27:2200042.

[133] Li Y, Jiang X, Qiu Y, Gao F, Xin H, Li D, et al. Latent and incubation periods of Delta, BA.1, and BA.2 variant cases and associated factors: a cross-sectional study in China. BMC Infect Dis 2024;24:294.

[134] Garcia-Knight M, Anglin K, Tassetto M, Lu S, Zhang A, Goldberg SA, et al. Infectious viral shedding of SARS-CoV-2 Delta following vaccination: A longitudinal cohort study. PLoS Pathog 2022;18:e1010802.

[135] Xia Q, Yang Y, Wang F, Huang Z, Qiu W, Mao A. Case fatality rates of COVIDL19 during epidemic periods of variants of concern: A meta-analysis by continents. Int J Infect Dis 2024;141:106950.

[136] Yuan Z, Shao Z, Ma L, Guo R. Clinical Severity of SARS-CoV-2 Variants during COVID-19 Vaccination: A Systematic Review and Meta-Analysis. Viruses 2023;15. 10.3390/v15101994.

[137] Ward T, Fyles M, Glaser A, Paton RS, Ferguson W, Overton CE. The real-time infection hospitalisation and fatality risk across the COVID-19 pandemic in England. Nat Commun 2024;15:4633.

[138] Liu Y, Rocklöv J. The effective reproductive number of the Omicron variant of SARS-CoV-2 is several times relative to Delta. J Travel Med 2022;29. 10.1093/jtm/taac037.

[139] Guo Z, Zhao S, Ryu S, Mok CKP, Hung CT, Chong KC, et al. Superspreading potential of infection seeded by the SARS-CoV-2 Omicron BA.1 variant in South Korea. J Infect 2022;85:e77–9.

[140] Del Águila-Mejía J, Wallmann R, Calvo-Montes J, Rodríguez-Lozano J, Valle-Madrazo T, Aginagalde-Llorente A. Secondary Attack Rate, Transmission and Incubation Periods, and Serial Interval of SARS-CoV-2 Omicron Variant, Spain. Emerg Infect Dis 2022;28:1224–8.

[141] Edelstein GE, Boucau J, Uddin R, Marino C, Liew MY, Barry M, et al. SARS-CoV-2 virologic rebound with nirmatrelvir-ritonavir therapy: An observational study: An observational study. Ann Intern Med 2023;176:1577–85.

[142] Ahmad SJ, Degiannis JR, Borucki J, Pouwels S, Rawaf DL, Lala A, et al. Fatality Rates After Infection With the Omicron Variant (B.1.1.529): How Deadly has it been? A Systematic Review and Meta-Analysis. J Acute Med 2024;14:51–60.

[143] Liu Y, Lillepold K, Semenza JC, Tozan Y, Quam MBM, Rocklöv J. Reviewing estimates of the basic reproduction number for dengue, Zika and chikungunya across global climate zones. Environ Res 2020;182:109114.

[144] Gao D, Lou Y, He D, Porco TC, Kuang Y, Chowell G, et al. Prevention and Control of Zika as a Mosquito-Borne and Sexually Transmitted Disease: A Mathematical Modeling Analysis. Sci Rep 2016;6:28070.

[145] Counotte MJ, Kim CR, Wang J, Bernstein K, Deal CD, Broutet NJN, et al. Sexual transmission of Zika virus and other flaviviruses: A living systematic review. PLoS Med 2018;15:e1002611.

[146] Lessler J, Ott CT, Carcelen AC, Konikoff JM, Williamson J, Bi Q, et al. Times to key events in Zika virus infection and implications for blood donation: a systematic review. Bull World Health Organ 2016;94:841–9.

[147] Agudelo S, Ventresca M. Modeling the spread of the Zika virus by sexual and mosquito transmission. PLoS One 2022;17:e0270127.

[148] Kucharski AJ, Funk S, Eggo RM, Mallet H-P, Edmunds WJ, Nilles EJ. Transmission Dynamics of Zika Virus in Island Populations: A Modelling Analysis of the 2013-14 French Polynesia Outbreak. PLoS Negl Trop Dis 2016;10:e0004726.

[149] Moreira J, Peixoto TM, Siqueira AM, Lamas CC. Sexually acquired Zika virus: a systematic review. Clin Microbiol Infect 2017;23:296–305.

[150] Cardona-Ospina JA, Henao-SanMartin V, Acevedo-Mendoza WF, Nasner-Posso KM, Martínez-Pulgarín DF, Restrepo-López A, et al. Fatal Zika virus infection in the Americas: A systematic review. Int J Infect Dis 2019;88:49–59.

