## Supplementary infomation for "The epidemiology of pathogens with pandemic potential: A review of key parameters and clustering analysis"

#### Supplementary information

1. Review 2
    1.1 Search strategy 2
    1.2 Literature search results 4
2. Parameter estimation 4
    2.1 Incubation period estimation 4
    2.2 Serial interval estimation 5
        2.2.1 LASV 5
        2.2.2 CCHFV 6
    2.3 Reproduction number estimation 7
        2.3.1 CCHFV 7
3. Clustering analysis 8
    3.1 Data Synthesis and Parameter Sampling 8
    3.2 Defining Pathogen Archetypes 9
        3.2.1 Per-Iteration K-Means Clustering 9
        3.2.2 Consensus Clustering 9
    3.3 Presymptomatic transmission 9
4. Sensitivity analysis 10
    4.1 Robustness to Parameter Selection 10
    4.2 Generalisability 17
5. Computational details 21
6. References 21

### 1. Review

#### 1.1 Search strategy

The primary search was conducted in PubMed to identify systematic reviews for the 19 pathogens selected. The search strategy, outlined in **Supplementary Table S1**, combined pathogen-specific terms with the keywords related to the epidemiological parameters of interest and "systematic review". Literature published up to July 5, 2024, was included, except for Nipah virus, for which the search extended to September 24, 2024. For SARS and MERS, the terms "NOT SARS-CoV-2 OR COVID-19" were added to exclude irrelevant studies focused on the COVID-19 pandemic. This stage aimed to explore the breadth of existing systematic reviews and identify parameters requiring further targeted searches.

**Supplementary Table S1. Search strategies**

| Pathogen | Parameter | Review |
|---|---|---|
| Crimean–Congo hemorrhagic fever OR CCHF OR Crimean Hemorrhagic Fever OR Congo Hemorrhagic Fever | basic reproductive number OR R0 OR basic reproduction number OR basic reproduction ratio OR basic reproductive rate | Systematic review |
| Ebola OR Ebola virus OR Ebola virus disease OR EBOV OR EVD | Incubation period OR incubation | |
| Marburg OR Marburg virus OR MARV OR Marburg virus disease OR MVD | Overdispersion OR dispersion parameter | |
| Lassa OR Lassa fever OR Lassa hemorrhagic fever OR Lassa virus | Latent period OR latency period OR pre-infectious period | |
| Middle East respiratory syndrome–related coronavirus OR MERS-CoV OR Middle East respiratory syndrome OR MERS | Infectious period | |
| Severe acute respiratory syndrome OR SARS OR SARS-CoV OR severe acute respiratory syndrome coronavirus | Serial interval | |
| Rift Valley fever OR RVF | Case fatality ratio OR Case fatality rate OR case-fatality risk OR CFR | |
| (Zika fever OR Zika virus disease OR Zika OR Zika virus OR ZIKV) | infection fatality ratio OR infection fatality rate OR IFR | |

| |
|---|
| (Nipah virus OR Nipah OR NiV) |
| H1N1 influenza A virus OR A/H1N1 OR H1N1 OR Spanish flu OR 1918 influenza pandemic |
| Influenza A virus subtype H2N2 OR A/H2N2 OR H2N2 OR 1957–1958 influenza pandemic OR Asian Flu |
| Influenza A virus subtype H3N2 OR A/H3N2 OR H3N2 OR Hong Kong Flu OR 1968 flu pandemic |
| 2009 swine flu OR Pandemic H1N1/09 virus OR 2009 swine flu pandemic |
| H5N1 OR Influenza A virus subtype H5N1 OR A/H5N1 |
| (COVID-19 OR SARS-CoV-2 OR 2019-nCoV OR "coronavirus disease 2019" OR "severe acute respiratory syndrome coronavirus 2") AND (wild type OR ancestral variant OR Wuhan variant) |
| (COVID-19 OR SARS-CoV-2 OR 2019-nCoV OR "coronavirus disease 2019" OR "severe acute respiratory syndrome coronavirus 2") AND (Alpha OR Alpha variant OR B.1.1.7) |
| (COVID-19 OR SARS-CoV-2 OR 2019-nCoV OR "coronavirus disease 2019" OR "severe acute respiratory syndrome coronavirus 2") AND (Delta OR Delta variant OR B.1.617.2 ) |
| (COVID-19 OR SARS-CoV-2 OR 2019-nCoV OR "coronavirus disease 2019" OR "severe acute respiratory syndrome coronavirus 2") AND (Omicron OR B.1.1.529 OR Omicron variant) |
| Mpox OR monkeypox OR Monkeypox virus OR MPV |

Following the database search, titles and abstracts were screened to identify relevant systematic reviews. Studies were included if they provided data on at least one of the specified parameters. Reviews that were not systematic or lacked relevant data were excluded. Full texts of the selected articles were then reviewed to extract comprehensive data, including summary statistics, contextual information, and probability distributions. All extracted data were documented in a centralised data extraction sheet.

To address gaps in the systematic review data, we conducted supplementary searches for additional sources. These included narrative reviews, rapid reviews, modeling studies, epidemiological investigations, and human infection studies. Due to the extensive volume of potential sources, no formal search strategy was applied at this stage. Data from these supplementary sources were extracted using the same standardised format and integrated into the central data sheet. This search was conducted up to January 2025.

The extracted parameters were categorised based on their source and nature. Parameters from modeling studies were classified into three categories where applicable: estimated values (directly estimated from empirical data), referenced values (taken from other articles), and assumed values (assumed for modeling purposes based on opinion or other data sources).

##### 1.2 Literature search results

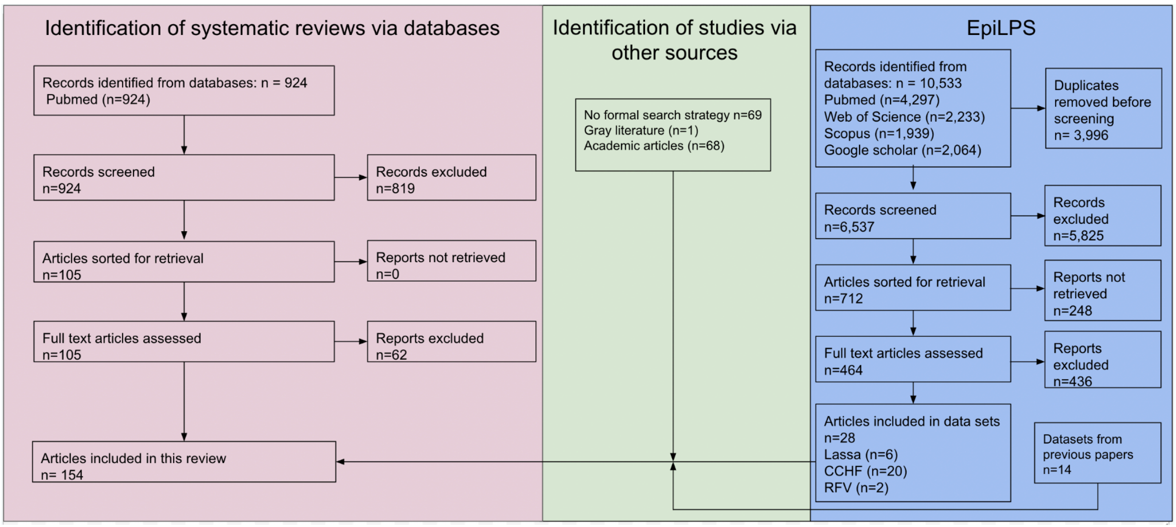

**Supplementary Figure S1. Flowchart illustrating the review process.**

#### 2. Parameter estimation

##### 2.1 Incubation period estimation

To complement the analyses derived from existing literature, the EpiLPS package [1–3] was used to estimate the incubation period of different pathogens when publicly available data permitted. EpiLPS relies on the methodology of Gressani et al. [2] to estimate the incubation distribution by using a flexible semi-parametric Bayesian approach based on Laplacian-P-splines.

Where data was not publicly available, it was collected on previous outbreaks using outbreak reports. For pathogens such as Lassa fever, Crimean-Congo haemorrhagic fever (CCHF), and Rift Valley fever (RVF), searches were conducted in May 2023 using the databases PubMed, Web of Science, Scopus, and Google Scholar. The search strategy combined pathogen-specific terms with keywords such as “community transmission,” “nosocomial transmission,” “reservoir transmission,” and “outbreak” or “cluster.” Relevant reports were identified based on their inclusion of exposure timelines and corresponding symptom onset data, which were subsequently used to estimate incubation periods.

##### 2.2 Serial interval estimation

To estimate serial intervals where appropriate, we used the model presented in Ward et al. 2024 [4] to fit both lognormal and gamma distributions to the number of onsets for a given day from outbreak data reporting the interval between the onset of illness in successive cases. To account for the double censoring we used the R package {primarycensored} [5,6]. Leave-one-out (LOO) analysis was used to select the final parametric model .

###### 2.2.1 LASV

Only one serial interval estimate for LASV was identified during the review stage. With a mean of 7.8 days (SD of 10.7) [7], this was deemed to be biased towards a shorter end of potential serial intervals for Lassa fever, considering the accepted incubation period of 7-21 days [8], which would in turn produce high presymptomatic transmission percentage estimates. We combined the dataset used for the Zhao et al. 2020 estimate (Lo Iacono et al. 2015 [9]) with data collected on Lassa outbreaks as described in **Supplementary Figure S1.** We estimated the serial interval distribution to have a mean of 11.8 days (95% CrI: 1.4–43) when fitted with a lognormal distribution, and a mean of 11.5 days (95% CrI: 0.9–34.6) when fitted with a gamma distribution (**Supplementary Figure S2**).

The leave-one-out information criterion (LOOIC) is computed and used to assess the model goodness-of-fit (with smaller values indicating a better fit). In this analysis, the lognormal distribution is the preferred choice as it demonstrates superior predictive accuracy with a smaller LOOIC as compared to the gamma distribution (534.9 vs 557.6).

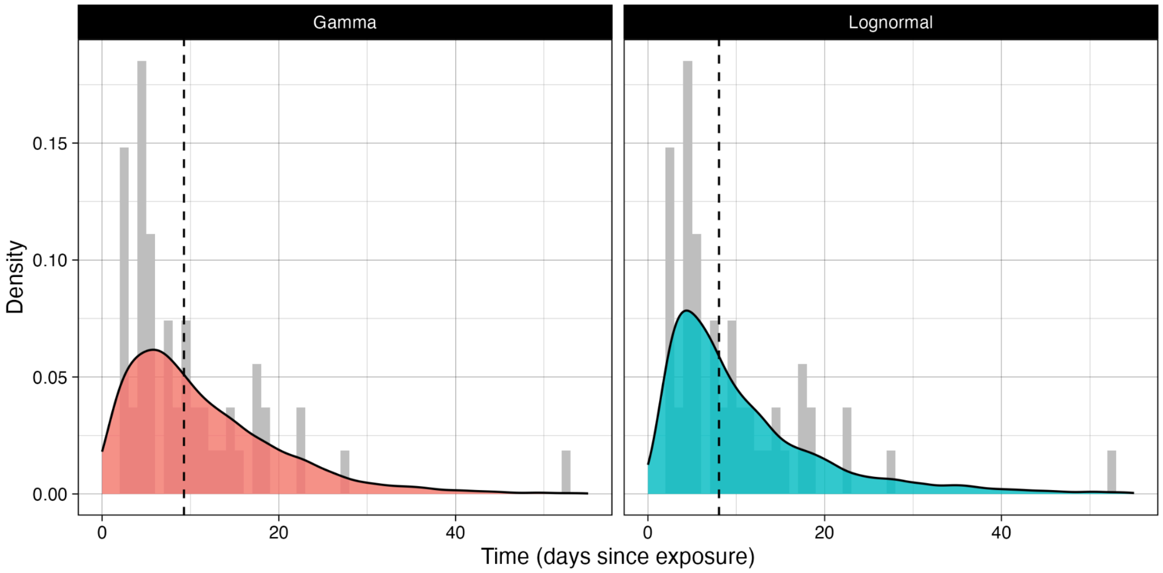

**Supplementary Figure S2. LASV serial interval distribution.**
We estimate the serial interval distribution for LASV to have a mean (vertical dashed line) of 11.8 days (95% CrI: 1.4–43) when fitted with a lognormal distribution, and a mean of 11.5 days (95% CrI: 0.9–34.6) when fitted with a gamma distribution.

##### 2.2.2 CCHFV

We did not identify any published estimates for CCHFV during the literature search. We collected data on CCHFV outbreaks as described in **Supplementary Figure S1** and estimated the serial interval distribution to have a mean of 12.0 days (95% CrI: 6.5–20.5) when fitted with a lognormal distribution, and a mean of 12.0 days (95% CrI: 3.0–27.2) when fitted with a gamma distribution (**Supplementary Figure S3**). In this analysis, the gamma distribution is the preferred choice as it demonstrates superior predictive accuracy with a smaller LOOIC as compared to the lognormal distribution (135.4 vs 304.1).

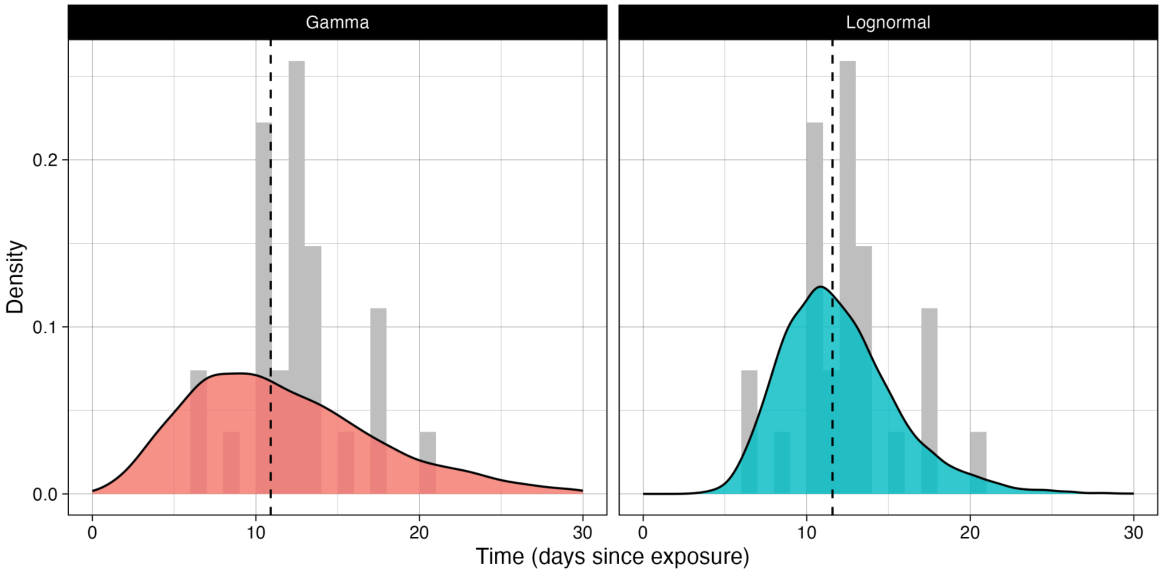

**Supplementary Figure S3. CCHFV serial interval distribution.**
We estimate the serial interval distribution to have a mean (vertical dashed line) of 12.0 days (95% CrI: 6.5–20.5) when fitted with a lognormal distribution, and a mean of 12.0 days (95% CrI: 3.0–27.2) when fitted with a gamma distribution.

#### 2.3 Reproduction number estimation

##### 2.3.1 CCHFV

No basic reproduction number estimates for CCHFV were identified in the literature search. To provide an estimate for the clustering analysis we estimated the basic reproduction number using the package {epichains} [10,11]. The parameters were estimated using Markov chain Monte Carlo (MCMC), implemented in the {MCMCpack package} [12]. We used this to estimate the reproduction number under the assumption of a negative binomial offspring distribution [10]. We used data on CCHFV outbreak clusters to estimate $R_0$. This data was collected from cases of CCHFV infected in the European Union/European Economic Area from 2013–2024 as reported by European Centre for Disease Prevention and Control (ECDC) [13]**.** We assumed cases with unknown transmission were tick-borne cases. We estimated the $R_0$ for CCHFV with a median of 0.03 (95% CrI: 0.004–0.09) (**Supplementary figure S4)**

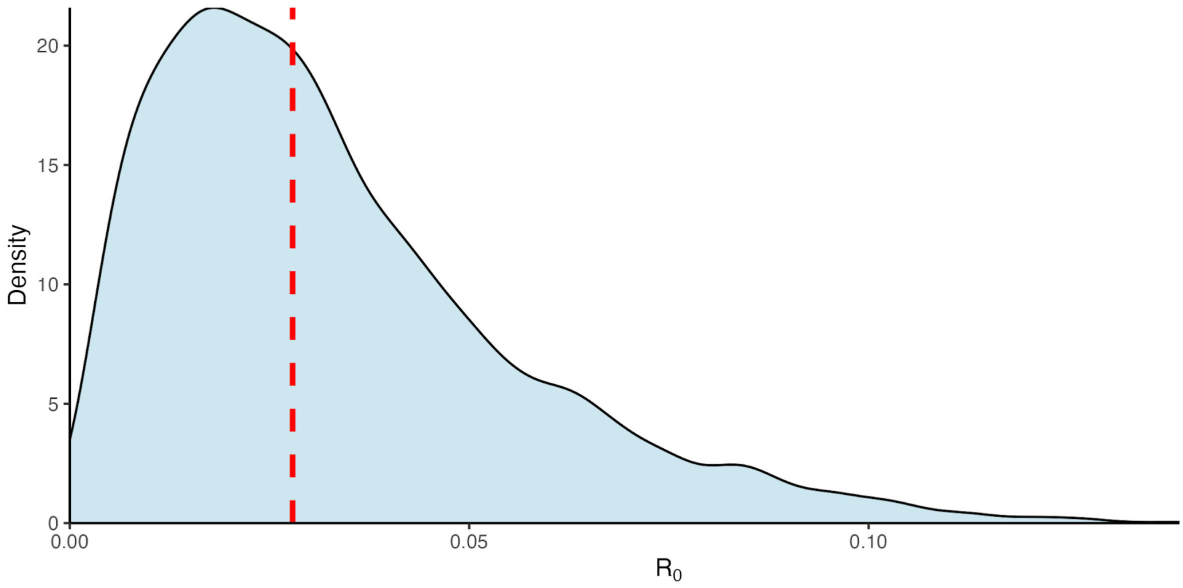

**Supplementary figure S4. Estimated CCHFV $R_0$ distribution in the EU from 2013-2024.**

Estimated $R_0$ distribution for CCHFV, red dotted line indicates the median value of 0.03 (95% CrI: 0.004–0.09).

### 3. Clustering analysis

##### 3.1 Data Synthesis and Parameter Sampling

For each pathogen, we assembled available estimates for a set of core parameters: the reproduction number (R), serial interval (SI), CFR, *k*, incubation period (IP), latent period (LP), infectious period (IP) (**Table 1**) and transmission route. For each continuous parameter from each study, we reconstructed a full probability distribution from its reported summary statistics. To ensure comparability across pathogens, we used a standardised approach for this reconstruction. The Beta distribution was used for parameters representing proportions bounded between 0 and 1 (CFR), while the Gamma distribution was used for all non-negative continuous values (R and time to key events).

To account for between-study heterogeneity, we used a non-parametric bootstrap-aggregation approach. In each of the 5,000 Monte Carlo iterations, for a given pathogen and parameter, we resampled the available studies with replacement. A single random value was then drawn from each selected study's reconstructed distribution. The final parameter value for that iteration was the average of these draws. Transmission routes were encoded as fixed binary indicators (presence/absence) and were not subject to sampling.

##### 3.2 Defining Pathogen Archetypes

To include the uncertainty induced from the parameter synthesis into the final groupings, we employed a two-stage clustering process. We performed an independent clustering analysis for each of the 5,000 Monte Carlo iterations, followed by a consensus step to synthesise these results into a single set of archetypes.

###### 3.2.1 Per-Iteration K-Means Clustering

Each of the 5,000 Monte Carlo iterations yielded a consistent parameter vector for every pathogen. Within each of these iterations, we performed an independent K-means clustering analysis. Continuous parameters were standardised (to a mean of 0 and a standard deviation of 1) across all pathogens for that specific iteration, while binary transmission route indicators were left on their original 0–1 scale. We then applied K-means clustering to these standardised features, using 25 random starts to ensure a stable solution. This resulted in 5,000 independent sets of cluster assignments, with each set representing a plausible grouping of pathogens

###### 3.2.2 Consensus Clustering

To derive a single, stable set of archetypes from these individual clustering results, we performed consensus clustering. We constructed a co-assignment matrix whose $i, j$ entry counted the number of iterations in which pathogens $i$ and $j$ were placed in the same K-means cluster. This matrix represents the probability that two pathogens are functionally similar across the full range of parameter uncertainty. This was then converted into a dissimilarity matrix ($Dij = 1 - P(i, j)$). We applied agglomerative hierarchical clustering with average linkage to this dissimilarity matrix to produce the final consensus grouping. The optimal number of consensus clusters (K) was guided by the average silhouette width (K=2–10) and finalised based on epidemiological interpretability [14].

The final hierarchical clustering was visualised as a dendrogram. For each resulting consensus cluster, we characterised its archetype by summarising the distribution of each parameter (mean and 95% confidence interval [CI]) across all Monte Carlo samples belonging to the pathogens within that cluster.

##### 3.3 Presymptomatic transmission

When presymptomatic transmission was included within the algorithm, we defined this as the probability that the serial interval was shorter than the incubation period (P(SI < IP)). To estimate this, we compiled SI and IP distributions for each pathogen. Within each iteration, one SI and one IP distribution were randomly selected from the available studies for a given pathogen, from which the distribution family and corresponding parameters were extracted.

Using these distributions, we calculated the presymptomatic proportion via a Monte Carlo simulation of 5,000 samples. We induced a positive rank correlation between the sampled pairs by generating a single vector of 5,000 random uniform quantiles and applying it to the inverse cumulative distribution functions of both the SI and IP. Any negative values were truncated to zero. The final estimate for each iteration was the proportion of the 5,000 pairs in which the sampled SI was less than the sampled IP. For influenza A subtypes, data on SI and IP distributions were pooled.

#### 4. Sensitivity analysis

##### 4.1 Robustness to Parameter Selection

To assess the stability of the identified pathogen archetypes, we repeated the clustering using alternative combinations of epidemiological parameters. We examined whether the consensus clusters were preserved when restricting the analysis to a reduced parameter set consisting of the reproduction number, serial interval, case fatality risk, presymptomatic transmission, and transmission route (**Supplementary Figure S5 & S6**). This was repeated to exclude presymptomatic transmission (**Supplementary Figure S7 & S8**). In addition the parameters used for **figure 2** were repeated to include presymptomatic transmission (**Supplementary Figure S9 & S10**). **Table S2** shows the archetype characterisation of **Supplementary figure S10** when K=6.

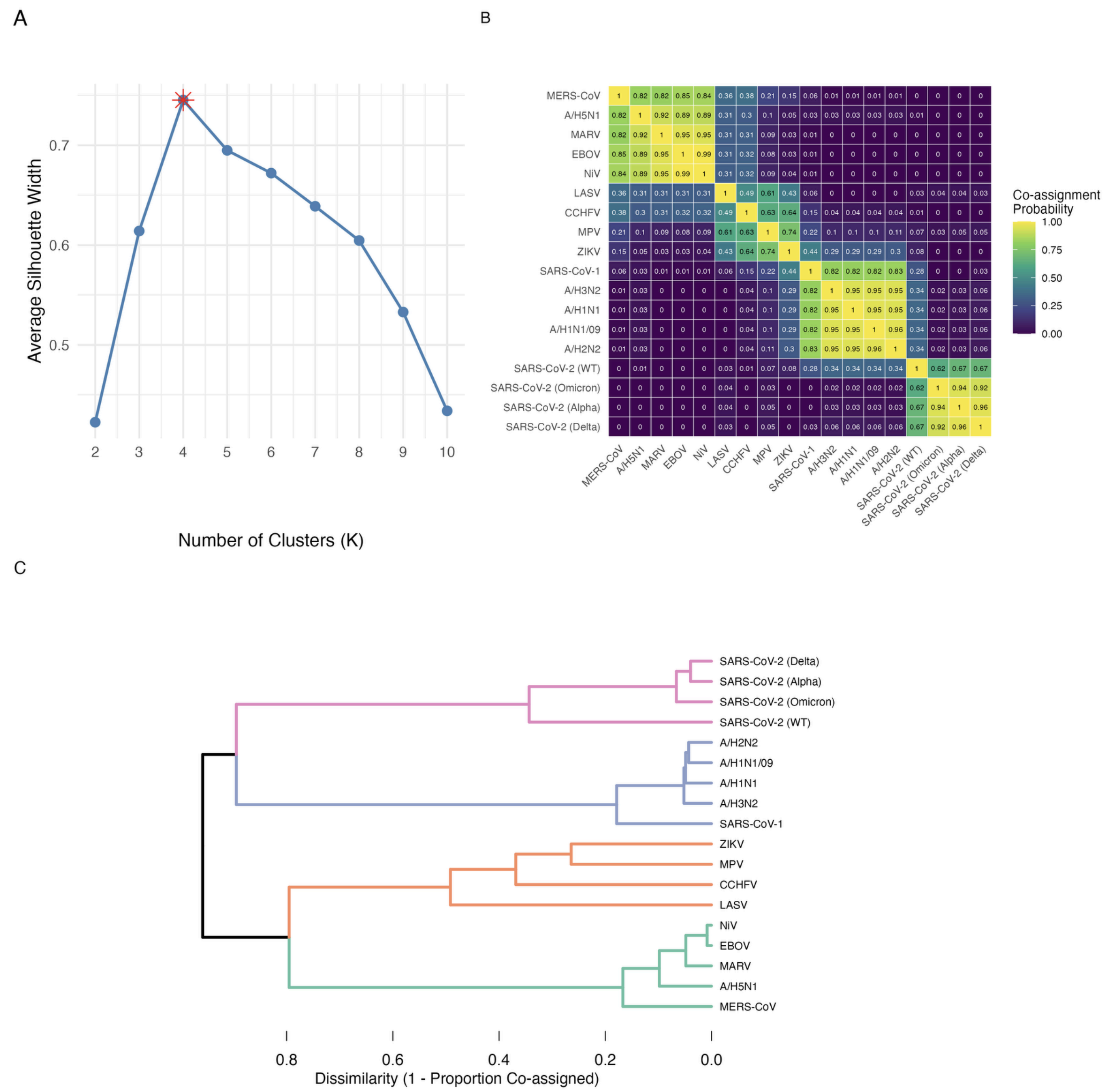

**Supplementary figure S5. Hierarchical clustering with $R_0$, serial interval, proportion of presymptomatic transmission, case fatality risk and transmission route.**

A) The optimal number of consensus clusters determined by maximizing the average silhouette width. The optimal number of clusters is identified as K=4 indicated by the red asterisk.

B) The co-assignment matrix, visualised as a heatmap, displays the proportion of the 5,000 Monte Carlo iterations in which each pair of pathogens was assigned to the same cluster.

C) The final, stable classification of 18 pathogens based on their epidemiological and transmission characteristics. The horizontal branch lengths represent the dissimilarity between clusters, with shorter branches indicating a higher frequency of co-assignment in the underlying Monte Carlo simulations.

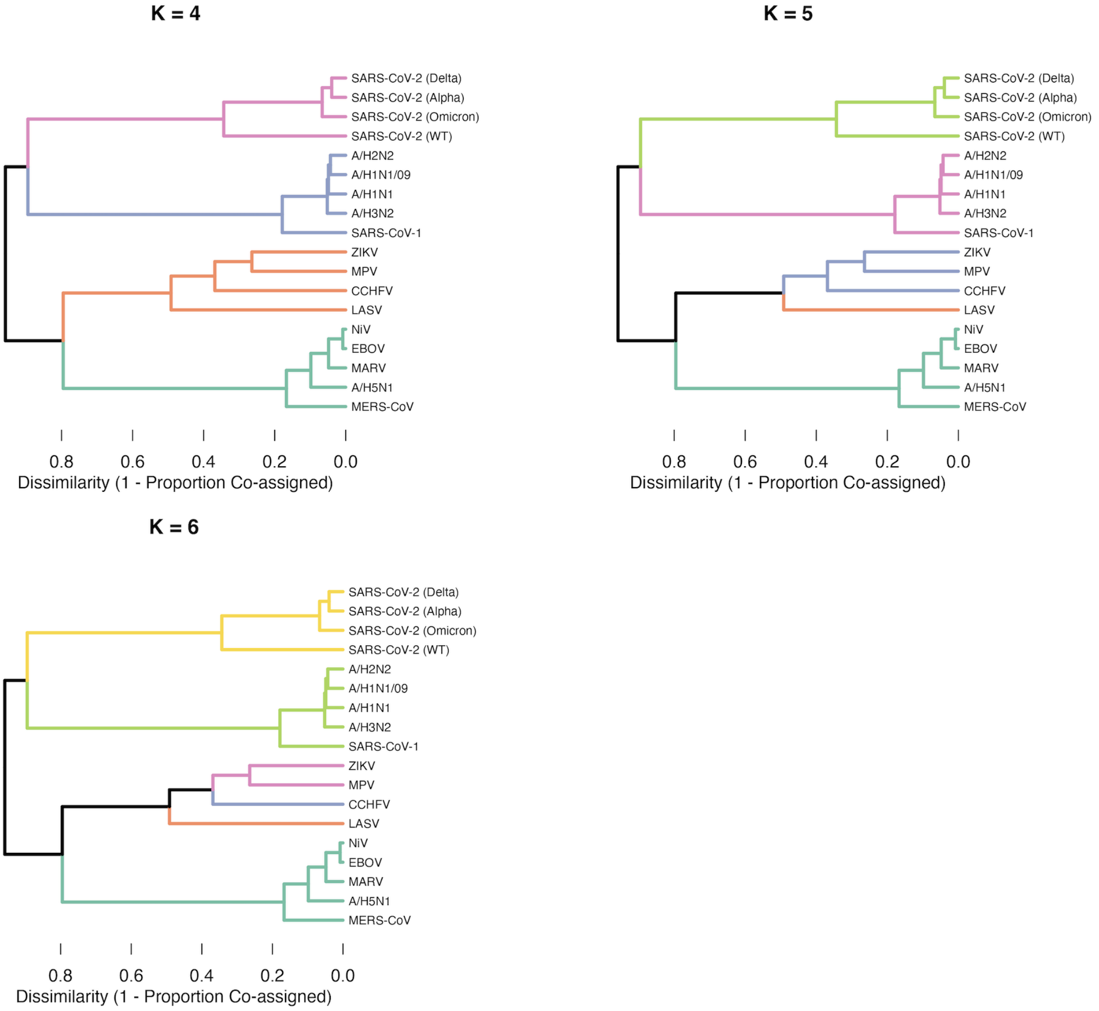

**Supplementary figure S6. Hierarchical clustering with $R_0$, serial interval, proportion of presymptomatic transmission, case fatality risk and transmission route. K=4, 5, 6**
The dendrogram from **Supplementary figure S5** is cut at K = 4, 5, and 6 clusters to show how pathogen groupings change as the tree is partitioned at different resolutions. Stable groupings appear where pathogens remain together across multiple values of K, whereas splits or reassignments indicate less well-defined relationships within the consensus structure.

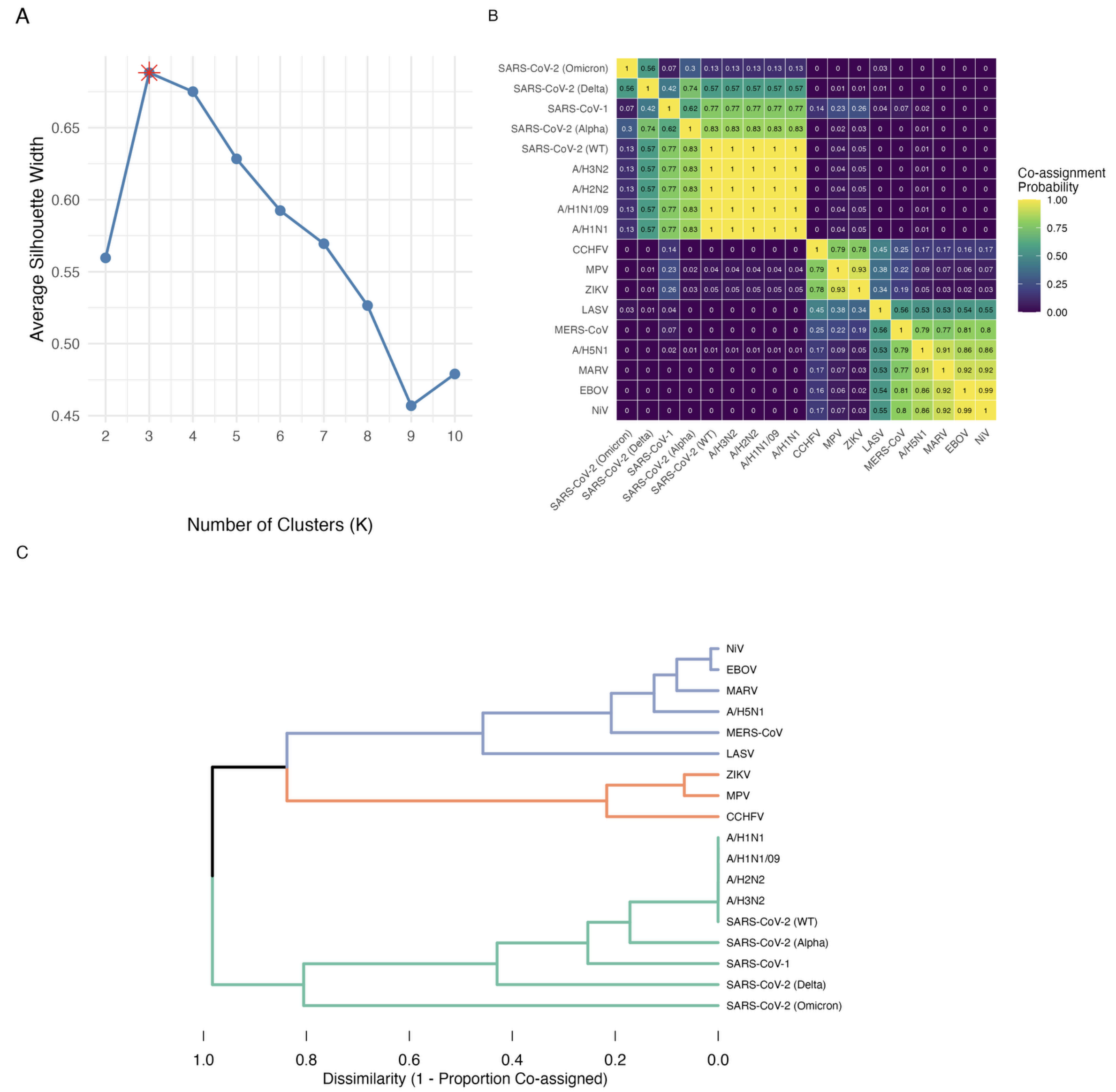

**Supplementary figure S7. Hierarchical clustering with $R_0$, serial interval, case fatality risk and transmission route.**

A) The optimal number of consensus clusters determined by maximizing the average silhouette width. The optimal number of clusters is identified as K=3 indicated by the red asterisk.

B) The co-assignment matrix, visualised as a heatmap, displays the proportion of the 5,000 Monte Carlo iterations in which each pair of pathogens was assigned to the same cluster.

C) The final, stable classification of 18 pathogens based on their epidemiological and transmission characteristics. The horizontal branch lengths represent the dissimilarity between clusters, with shorter branches indicating a higher frequency of co-assignment in the underlying Monte Carlo simulations.

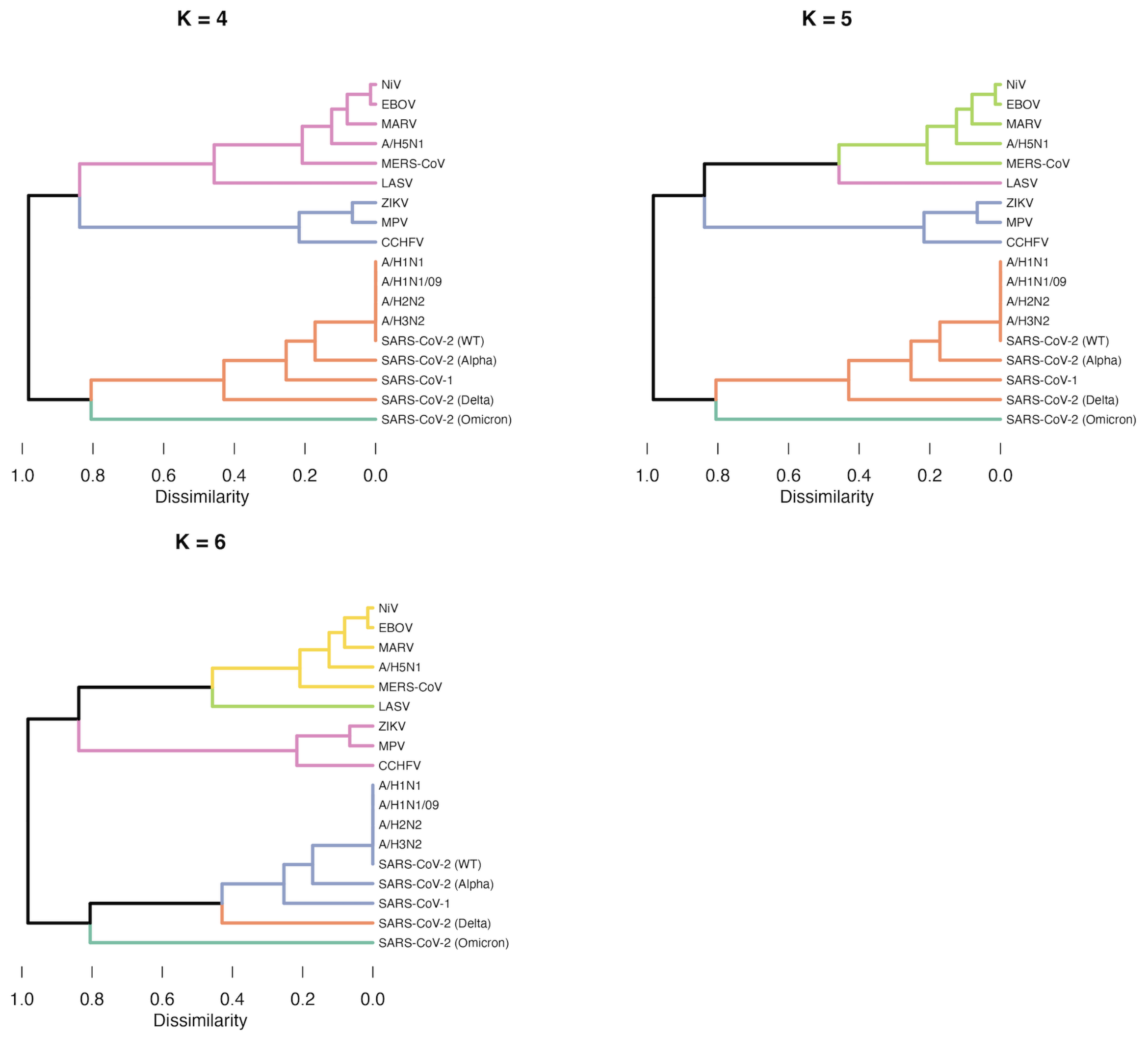

**Supplementary figure S8. Hierarchical clustering with $R_0$, serial interval, case fatality risk and transmission route. K=4, 5, 6**
The dendrogram from **Supplementary figure S7** is cut at K = 4, 5, and 6 clusters to show how pathogen groupings change as the tree is partitioned at different resolutions. Stable groupings appear where pathogens remain together across multiple values of K, whereas splits or reassignments indicate less well-defined relationships within the consensus structure.

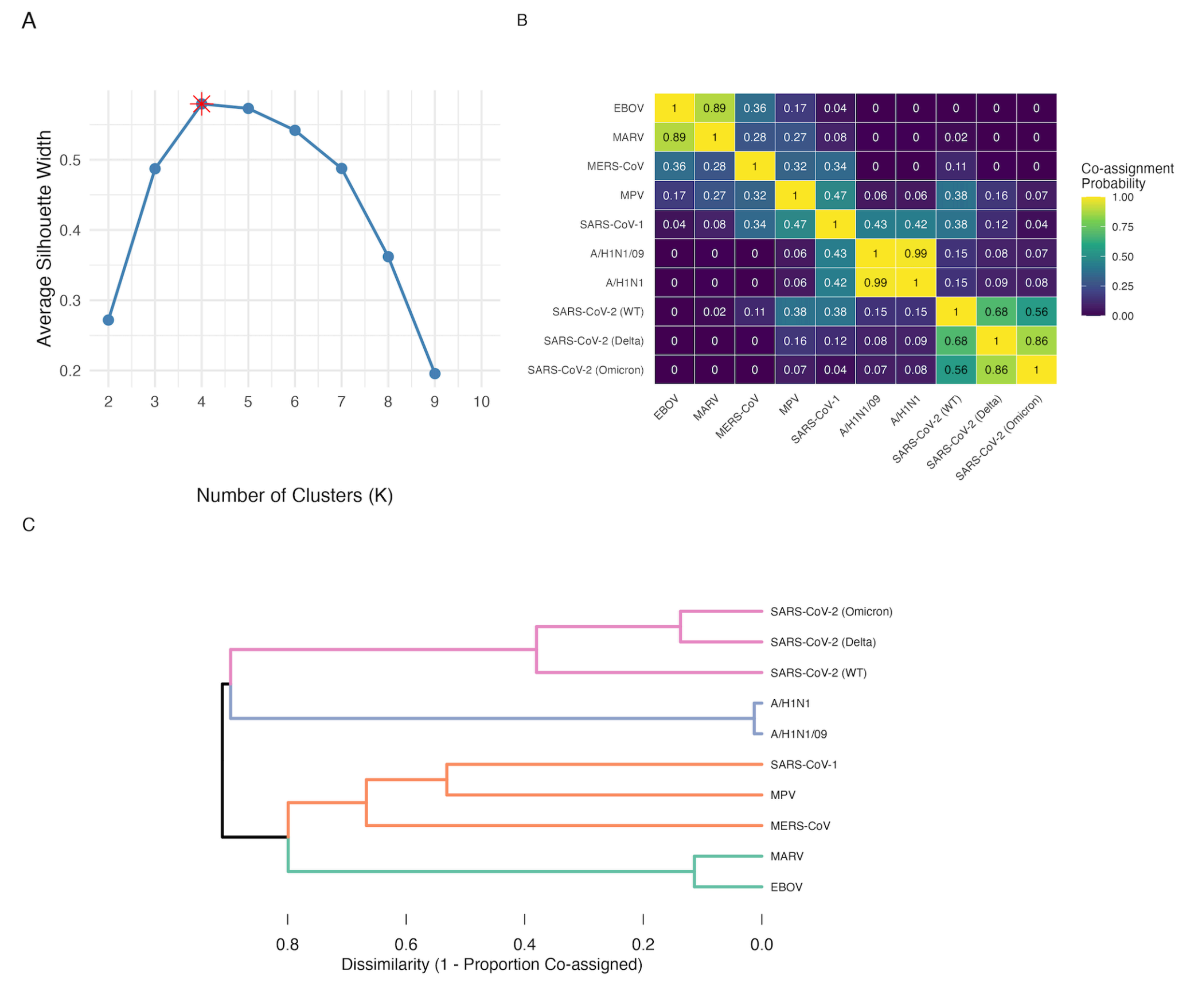

**Supplementary figure S9. Hierarchical clustering with $R_0$, serial interval, proportion of presymptomatic transmission, *k*, incubation, latent and infectious period, case fatality risk and transmission route.**

A) The optimal number of consensus clusters determined by maximizing the average silhouette width. The optimal number of clusters is identified as K=4 indicated by the red asterisk.

B) The co-assignment matrix, visualised as a heatmap, displays the proportion of the 5,000 Monte Carlo iterations in which each pair of pathogens was assigned to the same cluster.

C) The final, stable classification of 18 pathogens based on their epidemiological and transmission characteristics. The horizontal branch lengths represent the dissimilarity between clusters, with shorter branches indicating a higher frequency of co-assignment in the underlying Monte Carlo simulations.

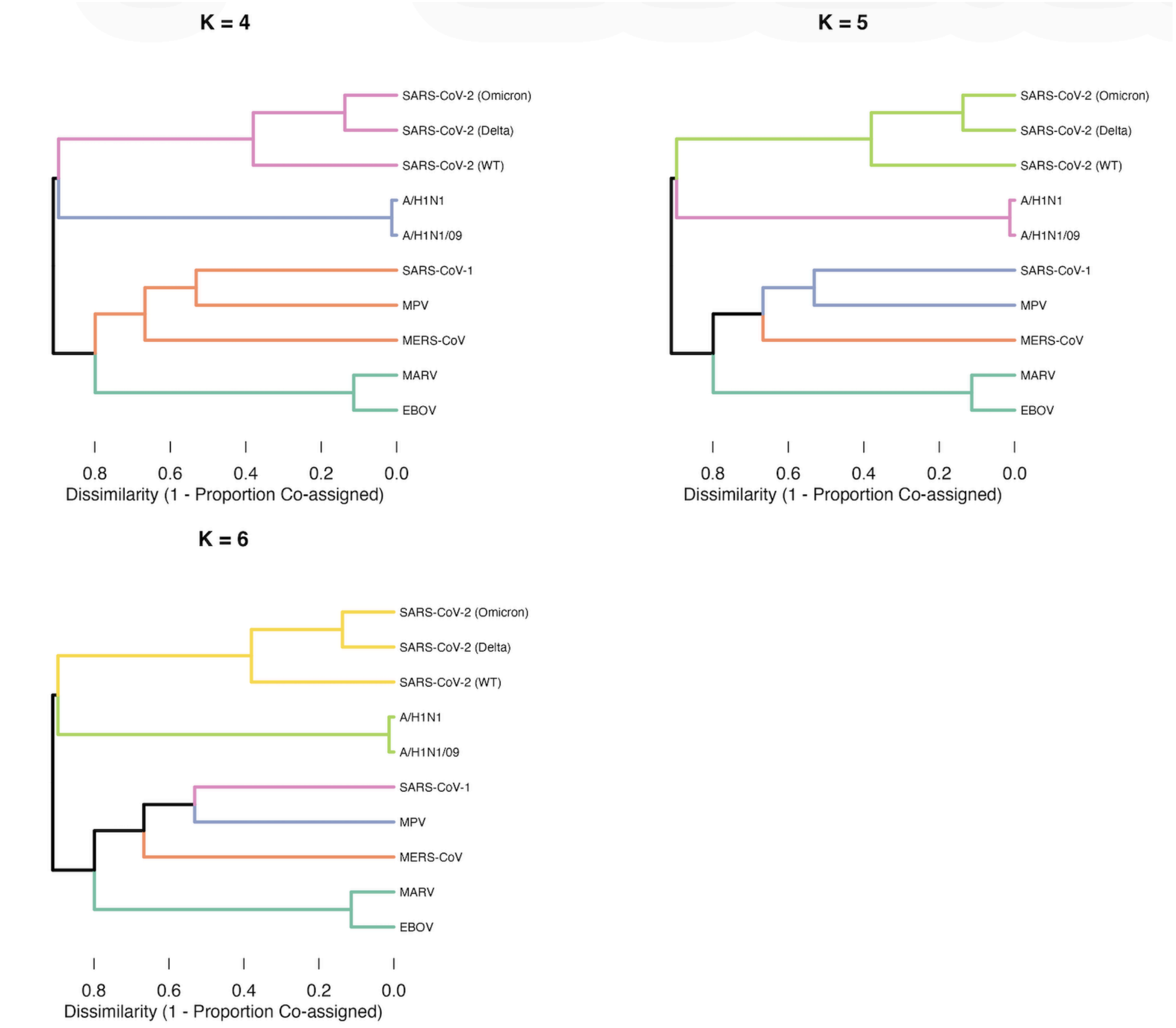

**Supplementary figure S10. Hierarchical clustering with $R_0$, serial interval, proportion of presymptomatic transmission, *k*, incubation, latent and infectious period, case fatality risk and transmission route. K=4, 5, 6**

The dendrogram from **Supplementary figure S9** is cut at K = 4, 5, and 6 clusters to show how pathogen groupings change as the tree is partitioned at different resolutions. Stable groupings appear where pathogens remain together across multiple values of K, whereas splits or reassignments indicate less well-defined relationships within the consensus structure.

| Supplementary Table S2. Archetype characterisation of Supplementary figure S10 when K=6 | | | | | | | | | | |
|---|---|---|---|---|---|---|---|---|---|---|
| Cluster | Pathogens | Archetype parameters (mean (95% CI)) | | | | | | | | Transmissionroute |
| | | R | *k* | Serial interval (d) | Incubation period (d) | Latent period (d) | Infectious period (d) | Presymptomatic transmission (%) | CFR (%) | |
| **1** | COVID-19 (WT, Delta,Omicron) | 5.89 (2.53–11.19) | 0.45 (0.10–0.81) | 3.97 (3.01–5.06) | 4.67 (3.34–6.55) | 4.19 (2.53–5.62) | 6.12 (3.13–10.18) | 80 (32–1.00) | 0.16 (95% CI: 0.0 - 0.4) | Respiratory (n=3) |
| **2** | EBOV, MARV | 1.87 (0.39–4.09) | 0.57 (0.18–1.05) | 11.90 (3.43–17.16) | 7.51 (4.42–9.37) | 8.72 (6.08–12.27) | 5.04 3.08–8.36) | 0.09 (0.0–0.4) | 60 (42–78) | Animal to human (n=2) Direct contact (n=2) |
| **3** | A/H1N1, A/H1N1pdm09 | 1.87 (1.44–2.83) | 4.41 (0.66–1.08) | 2.73 (2.12–3.36) | 1.50 (1.34–1.85) | 2.14 (1.52–3.05) | 2.13 (0.57–4.45) | 7.3 (0.0–63) | 0.12 (0.0–0.3) | Respiratory (n=2) |
| **4** | MERS-CoV | 1.42 (0.42–3.08) | 4.96 (0.00–48.97) | 12.43 (7.82–16.21) | 6.69 (5.70–7.94) | 3.00 (95% CI: 3.00 - 3.00) | 16.20 (9.56–24.21) | 8.1 (0.0–52) | 39 (26–53) | Animal to human (n=1) Respiratory (n=1) |
| **5** | MPV | 1.30 (0.80–1.84) | 0.48 (0.22–1.03) | 9.72 (7.42–14.26) | 8.56 (6.58–11.10) | 3.00 (3.00–3.00) | 6.33 (3.22–10.00) | 39 (0.0–1.00) | 5.4 (2.4–8.3) | Animal to human (n=1) Direct contact (n=1) |
| **6** | SARS-CoV-1 | 1.81 (1.08–2.53) | 0.21 (0.13–0.32) | 7.84 (1.64–19.00) | 4.33 (3.80–5.02) | 6.11 (5.00–6.81) | 8.20 (4.25–12.40) | 0.6 (0.0–11) | 9.6 (8.0–10) | Respiratory (n=1) |

#### 4.2 Generalisability

To explore the flexibility and potential application of our method to a broader range of pathogens, we expanded our analysis to 11 additional pathogens. This analysis focused on a core set of widely applicable and commonly reported parameters: $R_0$, incubation period, CFR and transmission route.

We include pathogens which had been excluded from the original clustering analysis (RVFV). In addition we include a wider array of health hazards outside of those included in the original pathogen set. This set included food and water borne pathogens (*Vibrio cholerae* [Cholera] and Norovirus), viruses more come in paediatrics (Measles virus, Enterovirus A71 [hand, foot, and mouth disease] and Human metapneumovirus), bioterrorism related pathogens (*Yersinia pestis* [Plague], Variola virus [Smallpox] and Bacillus anthracis

[Anthrax]), other vector borne pathogens (SFTS [Severe fever with thrombocytopenia syndrome] virus and Chikungunya), and a retrovirus (Human immunodeficiency virus). The parameters used for this analysis are listed in **supplementary Table S3**.

Given the substantially different composition of the pathogen set, we first determined the optimal number of clusters (k) for this new dataset, by calculating the average silhouette width for a range of k values (from 2 to 15) on the mean parameter values across all MCMC simulations. The k value that maximized the average silhouette width was then used for the subsequent per-iteration and consensus clustering steps (**Supplementary figure S11A**). The resulting dendrogram is shown in **Supplementary figure S11B.**

| Supplementary Table S2. List of parameters used in sensitivity analysis | | | |
|---|---|---|---|
| **Pathogen** | **Reproduction number** | **Incubation period** | **Case fatality risk** |
| Human immunodeficiency virus (HIV) | Range of 1–5 [15] | Mean of 7.8 years (90% CI: 4.2–15.0) [16] | 90% (untreated) [17] |
| *Vibrio cholerae* | Mean of 1.8 (95% CI: 1.3–2.3) [18] | Median of 1.4 days (95% CI, 1.3–1.6) [19] | Range of 4%–5% [18] |
| Measles virus | Median of 13.2 (Range: 4.6–44.4) [20] | Mean of 12.3 days (95% CI: 6.9–20.3) [21] | Median of 1.32 (95% CI: 1.28–1.36) in 0–34 year olds among community-based settings [22] |
| Norovirus | Median of 2.1 (interquartile range [IQR] 1.8-2.5) [23] | Median of 1.2 days (95% CI 1.1-1.2 days) [24] | 0.25 per 1000 cases (95 % CI 0.03–8.99) [25] |
| SFTS virus | Mean of 0.13 (95% CI:0.11–0.16) [26] | Mean of 10.0 days (IQR: 8.0–12.0) [26] | 10.5% (95% CI: 9.8–11.2) [26] |
| Enterovirus A71 | Median of 5.06 (IQR: 2.81–10.20) [27] | Assumed to be 5 days (Range 3–7 days) [28] | 1.7% (95% CI: 1.2–2.4) [29] |
| Human metapneumovirus (hMPV) | 1.2 [30] | Ranged from 4–9 days [31] | 3.9% among hMPV-positive cases (95% CI: 1.9–7.09) [32] |
| *Yersinia pestis* | 6.9 (95% CI: 5.2–9.3) (2017 plague outbreak Madagascar [pneumonic]) [33] | Mean 4.3 (SD: 1.8) (Pneumonic) [34] | 29% (95% CI: 22–37) (Pneumonic) [35] |
| Variola virus | 6.87 (95% CI: 4.52–10.1) [36] | Mean of 16.4 (95% CI: 15.6–17.9) [37] | 52% (95% CI: 48–56) - Variola major unvaccinated [38] |
| Chikungunya virus | Not applicable (NA) [39] | Median of 3.0 days (95% CI: 0.5–3.1) [40] | 0.8 deaths per 1,000 cases (Range 0–2.2 deaths per 1,000) [41] |
| Bacillus anthracis | NA [42] | 7 days (IQR, 4–9) - inhalation [43] | 45% - 2001 anthrax attacks [44] |

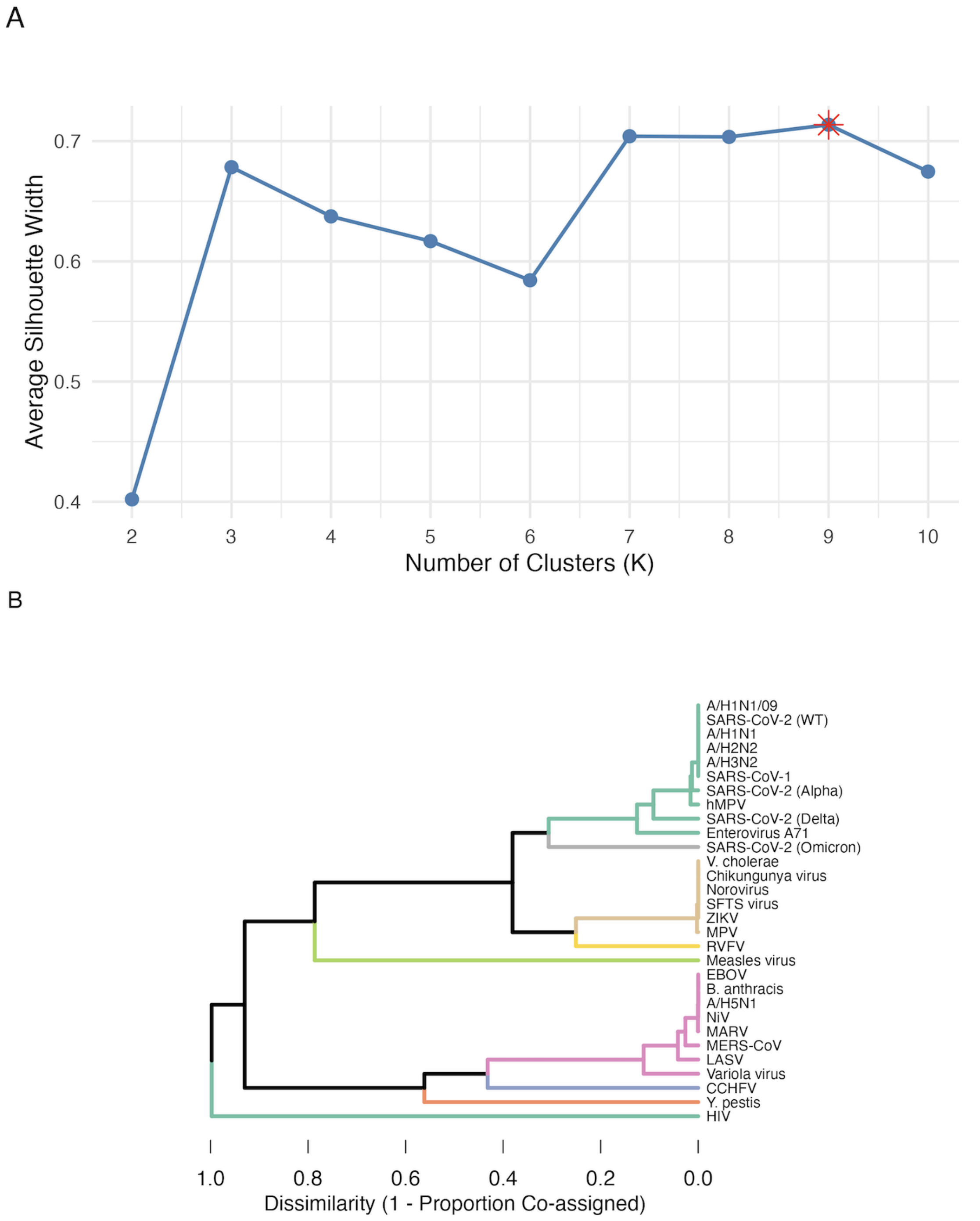

**Supplementary figure S11. Hierarchical clustering with R0, incubation period, CFR and transmission route.**

A) The optimal number of consensus clusters determined by maximizing the average silhouette width. The optimal number of clusters is identified as K=3 indicated by the red asterisk.

B) The final, stable classification of 18 pathogens based on their epidemiological and transmission characteristics. The horizontal branch lengths represent the dissimilarity between clusters, with shorter branches indicating a higher frequency of co-assignment in the underlying Monte Carlo simulations.

#### 5. Computational details

All analyses were implemented in R [45]. Code to reproduce this report is open on GitHub at https://github.com/oswaldogressani/Blueprint for incubation period estimates and https://github.com/Jward2847/archetypes# for serial interval, reproduction number and the clustering analysis.

#### 6. References

[1] Gressani O. EpiLPS: A Fast and Flexible Bayesian Tool for Estimating Epidemiological Parameters. 2021.
[2] Gressani O, Torneri A, Hens N, Faes C. Flexible Bayesian estimation of incubation times. Am J Epidemiol 2025;194:490–501.
[3] Gressani O, Wallinga J, Althaus CL, Hens N, Faes C. EpiLPS: A fast and flexible Bayesian tool for estimation of the time-varying reproduction number. PLoS Comput Biol 2022;18:e1010618.
[4] Ward J, Lambert JW, Russell TW, Azam JM, Kucharski AJ, Funk S, et al. Estimates of epidemiological parameters for H5N1 influenza in humans: a rapid review. medRxiv 2024:2024.12.11.24318702. https://doi.org/10.1101/2024.12.11.24318702.
[5] Charniga K, Park SW, Akhmetzhanov AR, Cori A, Dushoff J, Funk S, et al. Best practices for estimating and reporting epidemiological delay distributions of infectious diseases. PLoS Comput Biol 2024;20:e1012520.
[6] Abbott S, Brand S, Pearson C, Funk S, Charniga K. primarycensored: Primary Event Censored Distributions 2025. https://doi.org/10.5281/zenodo.13632839.
[7] Zhao S, Musa SS, Fu H, He D, Qin J. Large-scale Lassa fever outbreaks in Nigeria: quantifying the association between disease reproduction number and local rainfall. Epidemiol Infect 2020;148:e4.
[8] CDC. About. Lassa Fever 2024. https://www.cdc.gov/lassa-fever/about/index.html (accessed June 26, 2024).
[9] Lo Iacono G, Cunningham AA, Fichet-Calvet E, Garry RF, Grant DS, Khan SH, et al. Using modelling to disentangle the relative contributions of zoonotic and anthroponotic transmission: the case of lassa fever. PLoS Negl Trop Dis 2015;9:e3398.
[10] Azam JM, Funk S, Finger F. epichains: Simulating and Analysing Transmission Chain Statistics Using #> Branching Process Models 2024.
[11] Valle A. howto: How-To Guides For Outbreak Analytics R Packages. 2023.
[12] Martin AD, Quinn KM, Park JH. MCMCpack: Markov Chain Monte Carlo in R. Journal of Statistical Software 2011;42:22. https://doi.org/10.18637/jss.v042.i09.
[13] Cases of Crimean–Congo haemorrhagic fever infected in the EU/EEA, 2013–present. European Centre for Disease Prevention and Control 2021. https://www.ecdc.europa.eu/en/crimean-congo-haemorrhagic-fever/surveillance/cases-eu-since-2013 (accessed February 4, 2025).
[14] Hamerly G, Elkan C. Learning the k in k-means. Advances in Neural Information Processing Systems 2003;16.
[15] Fraser C, Riley S, Anderson RM, Ferguson NM. Factors that make an infectious disease outbreak controllable. Proc Natl Acad Sci U S A 2004;101:6146–51.
[16] Lui KJ, Darrow WW, Rutherford GW 3rd. A model-based estimate of the mean incubation period for AIDS in homosexual men. Science 1988;240:1333–5.
[17] Hawker J, Begg N, Blair I, Reintjes R, Weinberg J. Communicable Disease Control Handbook. Oxford, England: Blackwell Publishing; n.d.
[18] Phelps M, Perner ML, Pitzer VE, Andreasen V, Jensen PKM, Simonsen L. Cholera

epidemics of the past offer new insights into an old enemy. J Infect Dis 2018;217:641–9.
[19] Azman AS, Rudolph KE, Cummings DAT, Lessler J. The incubation period of cholera: a systematic review. J Infect 2013;66:432–8.
[20] Guerra FM, Bolotin S, Lim G, Heffernan J, Deeks SL, Li Y, et al. The basic reproduction number (R0) of measles: a systematic review. Lancet Infect Dis 2017;17:e420–8.
[21] Nishiura H. Early efforts in modeling the incubation period of infectious diseases with an acute course of illness. Emerg Themes Epidemiol 2007;4:2.
[22] Sbarra AN, Mosser JF, Jit M, Ferrari M, Ramshaw RE, O'Connor P, et al. Estimating national-level measles case-fatality ratios in low-income and middle-income countries: an updated systematic review and modelling study. Lancet Glob Health 2023;11:e516–24.
[23] Wang Y, Gao Z, Lu Q, Liu B, Jia L, Shen L, et al. Transmissibility quantification of norovirus outbreaks in 2016-2021 in Beijing, China. J Med Virol 2023;95:e29153.
[24] Lee RM, Lessler J, Lee RA, Rudolph KE, Reich NG, Perl TM, et al. Incubation periods of viral gastroenteritis: a systematic review. BMC Infect Dis 2013;13:446.
[25] Armah G, Lopman BA, Vinjé J, O'Ryan M, Lanata CF, Groome M, et al. Vaccine value profile for norovirus. Vaccine 2023;41 Suppl 2:S134–52.
[26] Fang X, Hu J, Peng Z, Dai Q, Liu W, Liang S, et al. Epidemiological and clinical characteristics of severe fever with thrombocytopenia syndrome bunyavirus human-to-human transmission. PLoS Negl Trop Dis 2021;15:e0009037.
[27] Zhang Z, Liu Y, Liu F, Ren M, Nie T, Cui J, et al. Basic Reproduction Number of Enterovirus 71 and Coxsackievirus A16 and A6: Evidence From Outbreaks of Hand, Foot, and Mouth Disease in China Between 2011 and 2018. Clin Infect Dis 2021;73:e2552–9.
[28] Ma E, Fung C, Yip SHL, Wong C, Chuang SK, Tsang T. Estimation of the basic reproduction number of enterovirus 71 and coxsackievirus A16 in hand, foot, and mouth disease outbreaks. Pediatr Infect Dis J 2011;30:675–9.
[29] Zhao YY, Jin H, Zhang XF, Wang B. Case-fatality of hand, foot and mouth disease associated with EV71: a systematic review and meta-analysis. Epidemiol Infect 2015;143:3094–102.
[30] Chittiprol N, Kandi V, Pinnelli VBK, Suvvari TK, Madamsetti N, Ca J, et al. The re-emergence of human metapneumovirus: Virus classification, characteristics, mechanisms of infection, clinical features, diagnosis, epidemiology, prevention, and treatment. Cureus 2025;17:e85259.
[31] Matsuzaki Y, Itagaki T, Ikeda T, Aoki Y, Abiko C, Mizuta K. Human metapneumovirus infection among family members. Epidemiol Infect 2013;141:827–32.
[32] Miyakawa R, Zhang H, Brooks WA, Prosperi C, Baggett HC, Feikin DR, et al. Epidemiology of human metapneumovirus among children with severe or very severe pneumonia in high pneumonia burden settings: the Pneumonia Etiology Research for Child Health (PERCH) study experience. Clin Microbiol Infect 2025;31:441–50.
[33] Nguyen VK, Parra-Rojas C, Hernandez-Vargas EA. The 2017 plague outbreak in Madagascar: Data descriptions and epidemic modelling. Epidemics 2018;25:20–5.
[34] Gani R, Leach S. Epidemiologic determinants for modeling pneumonic plague outbreaks. Emerg Infect Dis 2004;10:608–14.
[35] Jullien S, Garner P. Antibiotics for treating plague: a systematic review (executive summary). WHO guidelines for plague management: revised recommendations for the use of rapid diagnostic tests, fluoroquinolones for case management and personal protective equipment for prevention of post-mortem transmission [Internet], World Health Organization; 2021.
[36] Eichner M, Dietz K. Transmission potential of smallpox: estimates based on detailed data from an outbreak. Am J Epidemiol 2003;158:110–7.
[37] Nishiura H. Determination of the appropriate quarantine period following smallpox exposure: an objective approach using the incubation period distribution. Int J Hyg Environ Health 2009;212:97–104.
[38] Mack TM. Smallpox in Europe, 1950-1971. J Infect Dis 1972;125:161–9.

[39] CDC. Chikungunya: Causes and How It Spreads. Chikungunya Virus 2025. https://www.cdc.gov/chikungunya/causes-and-spread/index.html (accessed September 15, 2025).
[40] Rudolph KE, Lessler J, Moloney RM, Kmush B, Cummings DAT. Incubation periods of mosquito-borne viral infections: a systematic review. Am J Trop Med Hyg 2014;90:882–91.
[41] de Souza WM, Fumagalli MJ, de Lima STS, Parise PL, Carvalho DCM, Hernandez C, et al. Pathophysiology of chikungunya virus infection associated with fatal outcomes. Cell Host Microbe 2024;32:606–22.e8.
[42] Anthrax in humans. Anthrax in Humans and Animals. 4th edition, World Health Organization; 2008.
[43] Hendricks K, Person MK, Bradley JS, Mongkolrattanothai T, Hupert N, Eichacker P, et al. Clinical features of patients hospitalized for all routes of anthrax, 1880-2018: A systematic review. Clin Infect Dis 2022;75:S341–53.
[44] US Government Printing Office. USAMRIID’s MEDICAL MANAGEMENT OF BIOLOGICAL CASUALTIES HANDBOOK. US Government Printing Office; 2020.
[45] R Core Team. R: A Language and Environment for Statistical Computing 2021.
